# Lower risks of sodium glucose cotransporter 2 (SGLT2) inhibitors compared to dipeptidyl peptidase-4 (DPP4) inhibitors for new-onset non-alcoholic fatty liver disease and hepatocellular carcinoma in type 2 diabetes mellitus: A population-based study

**DOI:** 10.1101/2022.08.16.22278847

**Authors:** Oscar Hou In Chou, Jing Ning, Raymond Ngai Chiu Chan, Cheuk To Chung, Helen Huang, Kenrick Ng, Edward Christopher Dee, Sharen Lee, Apichat Kaewdech, Tong Liu, Fengshi Jing, Bernard Man Yung Cheung, Gary Tse, Jiandong Zhou

**Affiliations:** Division of Clinical Pharmacology, Department of Medicine, School of Clinical Medicine, Li Ka Shing Faculty of Medicine, University of Hong Kong, Hong Kong, China; Diabetes Research Unit, Cardiovascular Analytics Group, Hong Kong, China-UK Collaboration; Wuwei Hospital of Traditional Chinese Medicine, Wuwei 733000, Gansu, China; Department of Medical Oncology, University College London Hospital, London, UK; Department of Medical Oncology, St Bartholomew’s Hospital, London, UK; Department of Radiation Oncology, Memorial Sloan Kettering Cancer, New York, United States; Division of Internal Medicine, Faculty of Medicine, Prince of Songkla University, Hat Yai, Thailand; Tianjin Key Laboratory of Ionic-Molecular Function of Cardiovascular Disease, Department of Cardiology, Tianjin Institute of Cardiology, Second Hospital of Tianjin Medical University, Tianjin 300211, China; Institute for Artificial Intelligence, Guangdong Second Provincial General Hospital, Guangzhou, China; The University of North Carolina at Chapel Hill Project-China, Guangzhou, China; Kent and Medway Medical School, Canterbury, United Kingdom; Nuffield Department of Medicine, University of Oxford, Oxford, United Kingdom

**Author notes:** Correspondence to: Gary Tse, MD, PhD, FRCP, FFPH, Faculty of Health and Medical Sciences, University of Surrey, Guildford, United Kingdom Tianjin Institute of Cardiology, The Second Hospital of Tianjin Medical University Tianjin 300211, China, Kent and Medway Medical School, Canterbury, United Kingdom, Jiandong Zhou PhD, Nuffield Department of Medicine, University of Oxford, Oxford, United Kingdom. Co-first authors (Oscar Hou In Chou, Jing Ning).

**Keywords:** sodium glucose cotransporter 2 inhibitors (SGLT2I), dipeptidyl peptidase-4 inhibitors (DPP4I), non-alcoholic fatty liver disease (NAFLD), hepatocellular carcinoma (HCC), type-2 diabetes, cancer

## Abstract

**Background:** The association between sodium glucose cotransporter 2 inhibitors (SGLT2I) versus dipeptidyl peptidase-4 inhibitors (DPP4I) and the risks of non-alcoholic fatty liver disease (NAFLD) and hepatocellular carcinoma (HCC) are currently unknown.

**Methods:** This was a retrospective population-based cohort study including type-2 diabetes mellitus (T2DM) patients treated with either SGLT2I or DPP4I between 1^st^ January 2015 and 31^st^ December 2019 in Hong Kong. Patients with concurrent DPP4I and SGLT2I usage were excluded. The primary outcomes were NAFLD and HCC. The secondary outcomes included cancer-related mortality and all-cause mortality. Propensity score matching (1:1 ratio) was performed using the nearest neighbour search. Univariable and multivariable Cox regression was applied to identify significant predictors. Competing risks models and multiple approaches using the propensity score were performed.

**Results:** This cohort included 62699 patients with T2DM, amongst which 22154 patients were on SGLT2I and 40545 patients were on DPP4I. After matching (44308 patients), 1090 patients developed new-onset NAFLD (Incidence: 4.6; 95% Confidence interval [CI]: 4.3-4.9) and 187 patients developed HCC (Incidence: 0.8; 95% CI: 0.7-0.9). Overall, SGLT2I was associated with lower risks of NAFLD (Hazard ratio [HR]: 0.39; 95% CI: 0.34-0.46), and HCC (HR: 0.46; 95% CI: 0.29-0.72) compared to DPP4I after adjustments. SGLT2I was also associated with lower risks of cancer-related mortality (HR: 0.29; 95% CI: 0.23-0.37) and all-cause mortality (HR: 0.28; 95% CI: 0.25-0.31). However, amongst patients with hepatitis B virus infection, SGLT2I was associated with higher risks of HCC (HR: 3.28; 95% CI: 1.21-8.90). The results were consistent in competing risk models and different matching approaches.

**Conclusion:** SGLT2I was associated with lower risks of NAFLD, and HCC compared to DPP4I after propensity scores matching and adjustments.

**Lay summary:** The association between two antidiabetic medications, SGLT2I and DPP4I, and the risks of fatty liver disease and liver cancer have not been explored. In our study, SGLT2I was associated with a lower risk of fatty liver disease and liver cancer compared to DPP4I amongst patients with type 2 diabetes. However, DPP4I was associated with lower risks of liver cancer compared to SGLT2I among patients with hepatitis B virus infection

**Central illustration:** 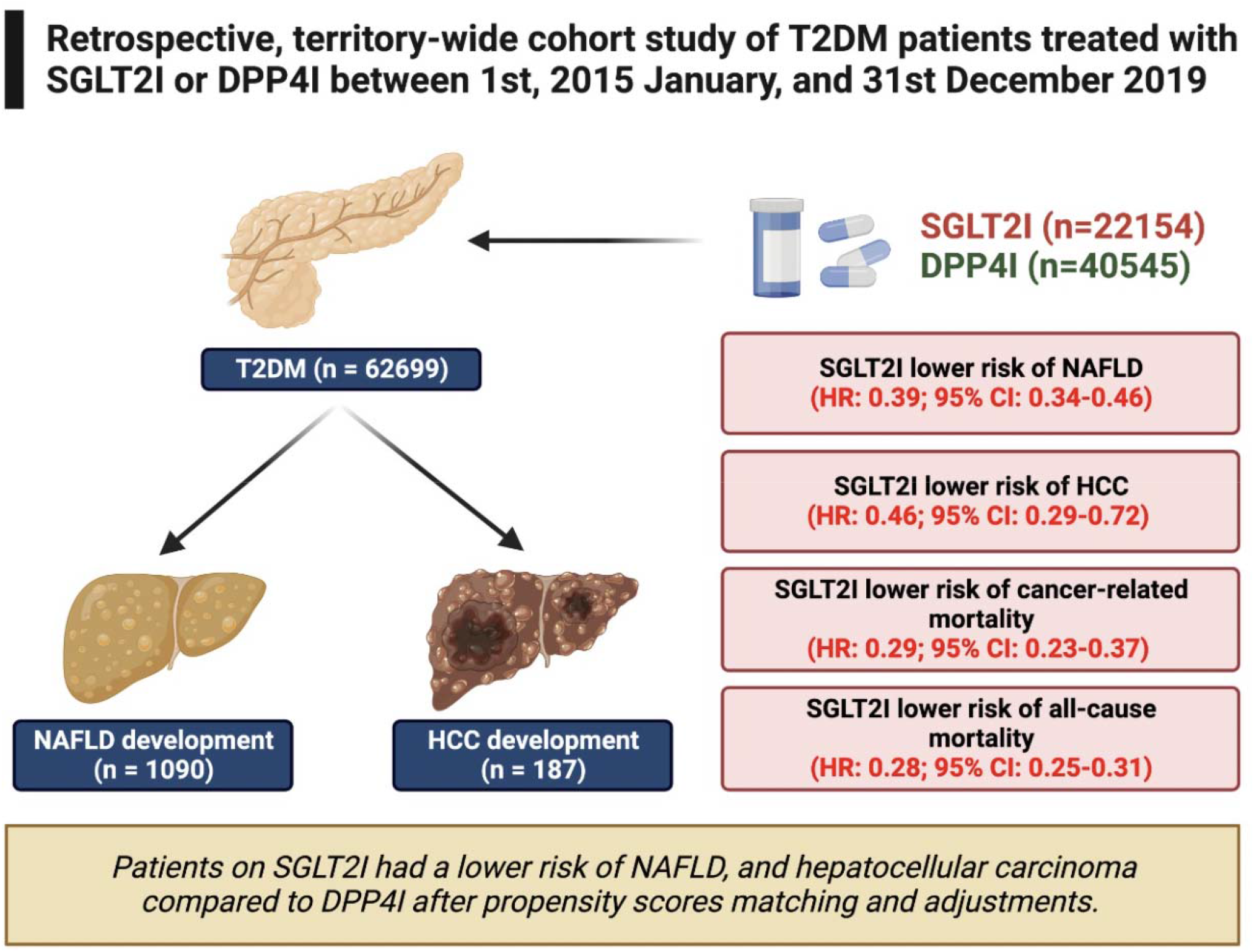

## Introduction

In the past several decades, the incidence of hepatocellular carcinoma (HCC) has continued to rise [1]. In 2020, HCC was the sixth most common type of cancer worldwide, accounting for most cases of liver cancers and ranked the third leading cause of cancer death [2]. The geographical distribution of disease burden varies significantly, with the highest incidences rates observed in Western Pacific and African regions [3]. Hong Kong has a high rate of HCC due to the prevalence of the hepatitis B virus (HBV) within the territory; HBV accounted for 80% of all cases of HCC from 1992 to 2016 [4]. The prognosis of advanced-stage HCC remains poor as symptoms rarely appear during the early stages of the disease and high-risk patients may not be provided timely surveillance [5]. Patients with chronic hepatitis C (HCV) infection, chronic HBV infection, non-alcoholic fatty liver disease (NAFLD) or alcohol-associated liver disease have a higher likelihood of developing cirrhosis and eventually HCC. [6].

Recently, it has been found that patients with type 2 diabetes mellitus (T2DM) are also associated with a higher propensity to develop HCC. A systemic review and meta-analysis revealed that diabetic patients had a 2.31-fold increased risk of HCC and 2.43-fold increased risk of HCC-related death compared to nondiabetic patients [7]. Meanwhile, antidiabetic medications such as metformin have demonstrated protective effects against the disease [8-10]. This led to the growing interest in exploring the prevent effects of novel antidiabetic medications such as sodium-glucose cotransporter 2 inhibitors (SGLT2I) and dipeptidyl peptidase-4 inhibitors (DPP4I) in HCC.

As of now, several anti-diabetic medications show promising anti-tumour effects against HCC. SGLT2I reduces the blood glucose level by blocking the glucose reabsorption at the S1 segment of the proximal convoluted tubules of the kidney. Clinical evidence surrounding the effects of SGLT2I on NAFLD and HCC is relatively scarce. Several studies with a relatively short follow-up suggested that SGLT2I may reduce the risk of NAFLD, which may be indirectly linked to HCC [11-13]. A case report observed spontaneous regression of HCC post-SGLT2I treatment due to the reduction of angiogenesis-related cytokines [14].

DPP4I is an incretin-based antidiabetic drug which inhibits glucagon-like peptide-1 degradation [15]. DPP4I was previously demonstrated to lower the risks of HCC amongst HCV-infected patients in a retrospective cohort study [16]. A case report by Yamamoto *et al*. found a spontaneous regression in HCC after four weeks of DPP4I treatment [17]. Indeed, a meta-analysis demonstrated that DPP4I does not increase the risks of developing overall cancer compared to patients treated with a placebo or other drugs. However, direct comparison between SGLT2I and DPP4I on new-onset NAFLD and HCC remains limited. Therefore, the present study aims to compare the association of SGLT2I versus DPP4I on the risk of new-onset NAFLD and HCC in T2DM patients from Hong Kong.

## Methods

### Study design and population

This was a retrospective, territory-wide cohort study of T2DM patients treated with SGLT2I or DPP4I between 1^st^ January 2015, and 31^st^ December 2019, in Hong Kong. Patients were followed up until 31^st^ December 2020, or until death. This study was approved by The Joint Chinese University of Hong Kong–New Territories East Cluster Clinical Research Ethics Committee and complied with the Declaration of Helsinki. The patients were identified from the Clinical Data Analysis and Reporting System (CDARS), a territory-wide database that centralizes patient information from individual local hospitals to establish comprehensive medical data, including clinical characteristics, disease diagnosis, laboratory results, and drug treatment details. The system has been used by local teams in Hong Kong to conduct comparative studies [18-20] and recently by our team to examine adverse cardiovascular outcomes for SGLT2I/DPP4I users [21, 22]. Patients were excluded if they exhibited any of the following criteria: 1) with both DPP4I and SGLT2I use 2) without complete demographics 3) without mortality data 4) less than 18 years old 5) died within 30 days after drug exposure 6) prior HCC 7) new onset HCC development less than 1 year after drug exposure **(Figure 1)**.

**Figure 1.**
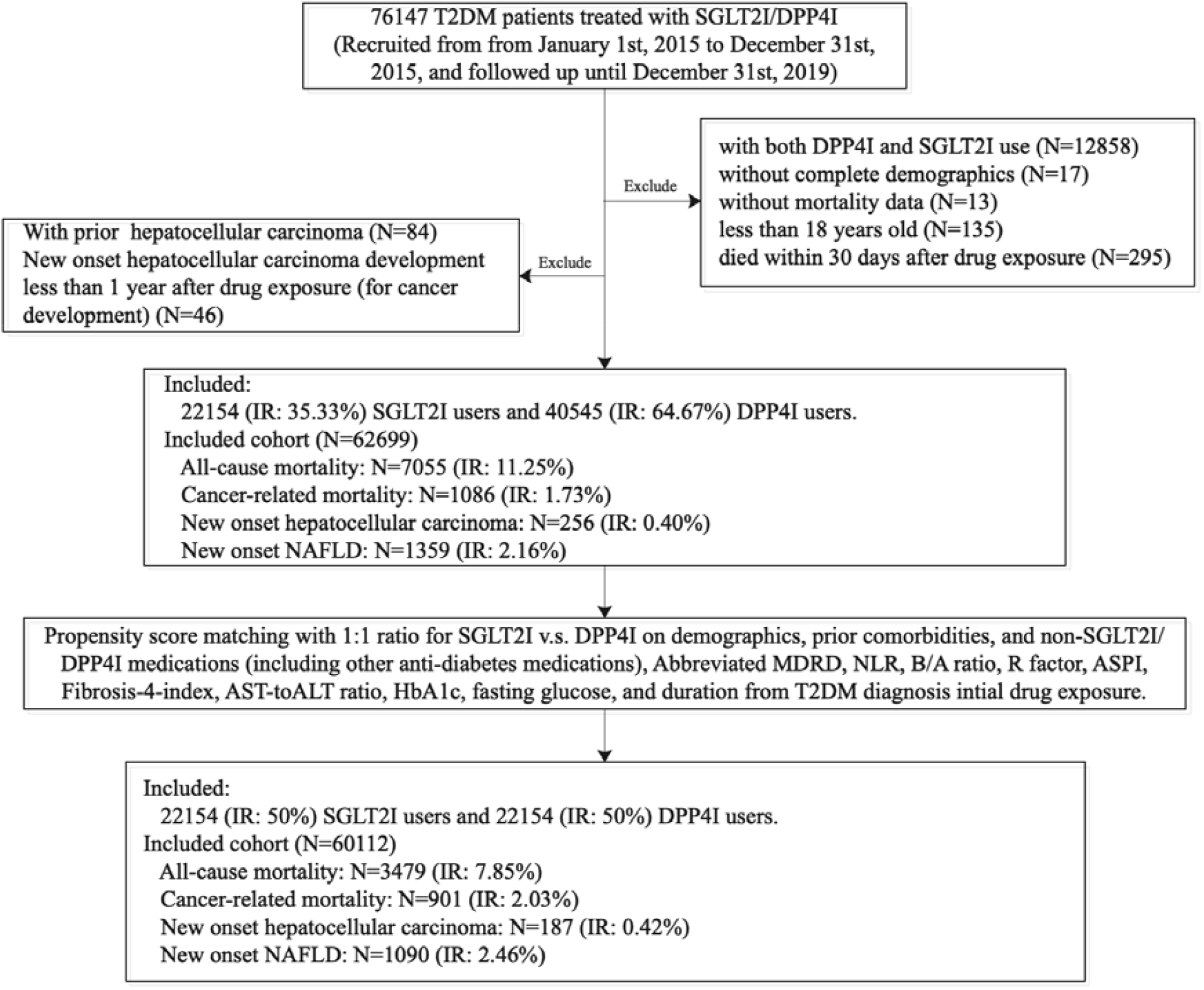
Procedures of data processing. IR: Incidence rate; SGLT2I: Sodium-glucose cotransporter-2 inhibitors; DPP4I: Dipeptidyl peptidase-4 inhibitors.

Patients’ demographics include gender and age of initial drug use (baseline), clinical and biochemical data were extracted for the present study. Prior comorbidities were extracted using the *International Classification of Diseases Ninth Edition* (ICD-9) codes (**Supplementary Table 1**). Charlson’s standard comorbidity index was also calculated. Both cardiovascular medications and anti-diabetic agents were also extracted. The baseline laboratory examinations, including the complete blood count, renal and liver biochemical tests, and the lipid and glucose profiles were extracted. HBV infection was defined by both the ICD-9 codes for HBV infection and positive HBsAg laboratory results. Hepatitis C virus (HCV) infection was defined by both the ICD-9 codes for HCV infection and positive anti-HCV laboratory results. The standard deviation variability measure for the lipid and glucose profiles were also calculated **(Supplementary Table 2)**. The estimated glomerular filtration rate (eGFR) was calculated using the abbreviated modification of diet in renal disease (MDRD) formula [23].

### Adverse outcomes and statistical analysis

The primary outcome included NAFLD (ICD9: 571.5, 571.8, 571.9) and HCC (ICD9:155). Mortality data were obtained from the Hong Kong Death Registry, a population-based official government registry with the registered death records of all Hong Kong citizens linked to CDARS. Mortality was recorded using the *International Classification of Diseases Tenth Edition* (ICD-10) coding. The endpoint date of interest for eligible patients was the event presentation date. The endpoint for those without primary outcome presentation was the mortality date or the endpoint of the study (31^st^ December 2020).

Descriptive statistics are used to summarize baseline clinical and biochemical characteristics of patients with SGLT2I and DPP4I use. For baseline clinical characteristics, the continuous variables were presented as mean (95% confidence interval [CI]/standard deviation [SD])) and the categorical variables were presented as total numbers (percentage). Continuous variables were compared using the two-tailed Mann-Whitney U test, whilst the two-tailed Chi-square test with Yates’ correction was used to test 2×2 contingency data. Propensity score matching with 1:1 ratio for SGLT2I use versus DPP4I use based on demographics, Charlson comorbidity index, prior comorbidities, non-SGLT2I/ DPP4I medications were performed using the nearest neighbour search strategy. We used Stata software (Version 16.0) to conduct the propensity score matching procedures.

Baseline characteristics between patients with SGLT2I and DPP4I use before and after matching were compared with standardized mean difference (SMD), with SMD<0.20 regarded as well-balanced between the two groups. Proportional Cox regression models were used to identify significant risk predictors of adverse study outcomes. Subgroup analysis was conducted to identify the risk predictors stratified by age, gender, and HBV infection status. Cause-specific and subdistribution hazard models were conducted to consider possible competing risks. Multiple propensity adjustment approaches were used, including propensity score stratification [24], propensity score matching with inverse probability of treatment weighting [25] and propensity score matching with stable inverse probability weighting [26]. The hazard ratio (HR), 95% CI and P-value were reported. Statistical significance is defined as P-value < 0.05. All statistical analyses were performed with RStudio software (Version: 1.1.456) and Python (Version: 3.6).

## Results

### Baseline characteristics

This was a retrospective, territory-wide cohort study of 76147 patients with T2DM treated with SGLT2I/DPP4I between 1^st^ January 2015 and 31^st^ December 2019 in Hong Kong. Patients during the aforementioned period were enrolled and followed up until 31^st^ December 2020 or until their deaths. Patients with both DPP4I and SGLT2I use (N=12858), without complete demographics (N=17), without mortality data (N=13), less than 18 years old (N=135), died within 30 days after drug exposure (N=295), prior HCC (N=84), new onset HCC development less than 1 year after drug exposure (N=46) were excluded **(Figure 1)**.

After exclusion, this study included a total of 62699 patients with T2DM (mean age: 63.3 years old [SD: 12.9]; 55.18% males). 22154 patients (35.33%) used SGLT2Is, and 40545 patients (64.67%) used DPP4Is. The DPP4I and SGLT2I cohorts were comparable after matching **(Supplementary Figure 1)**. In the matched cohort, 1090 (2.46%) patients developed NAFLD, and 187 patients (0.42%) developed HCC. The characteristics of patients are shown in **Table 1, Supplementary Table 3 and 4**.

**Table 1.**
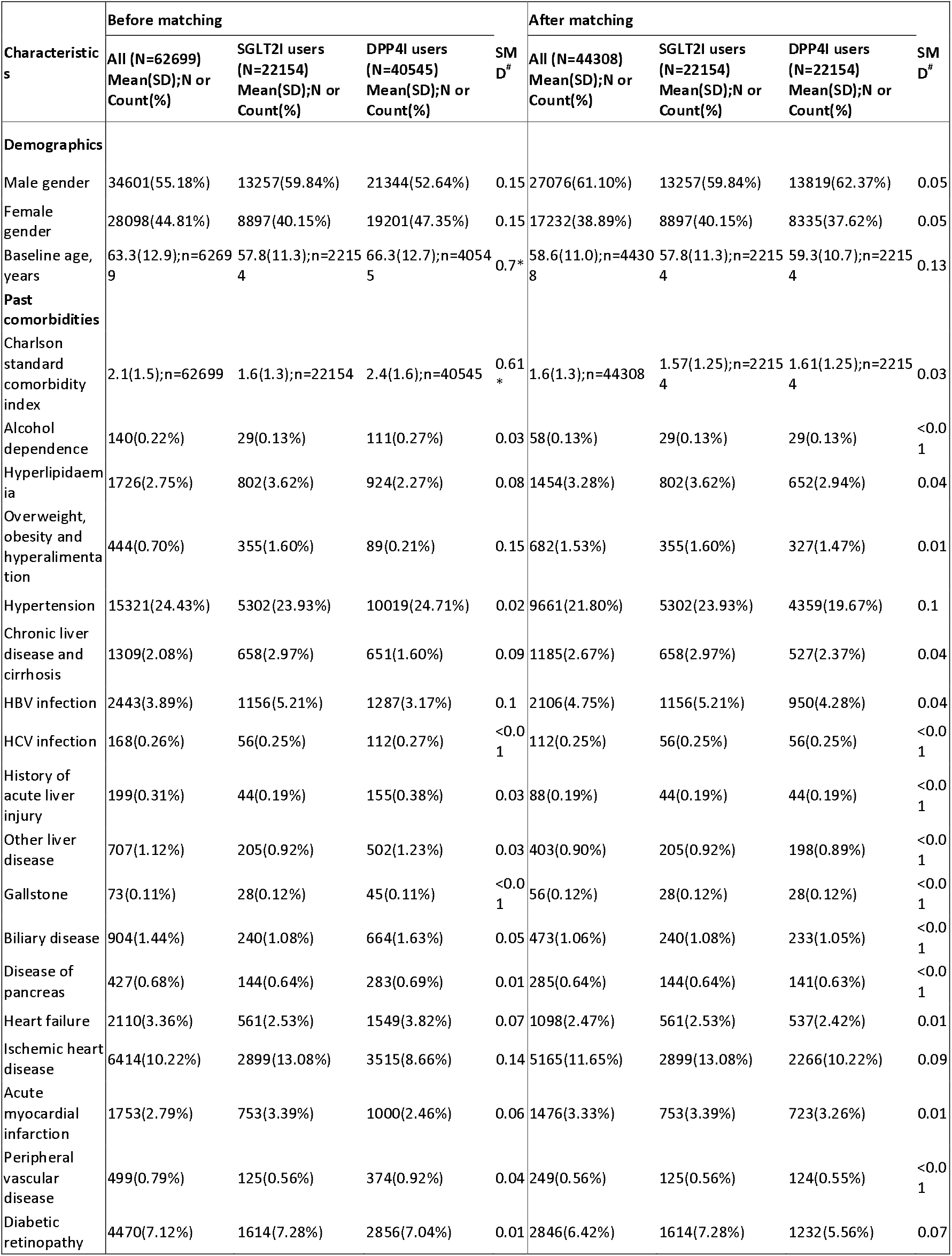
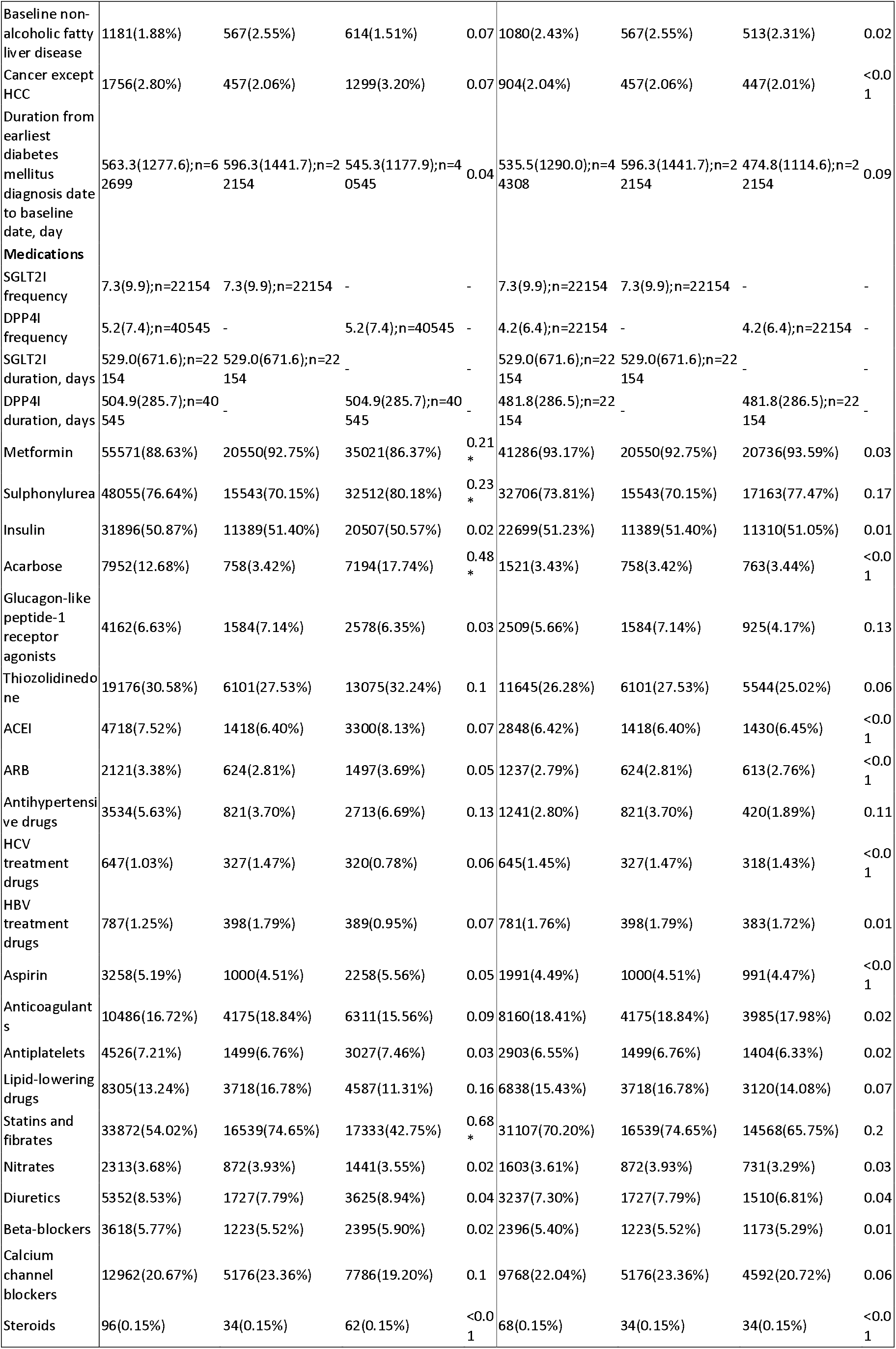
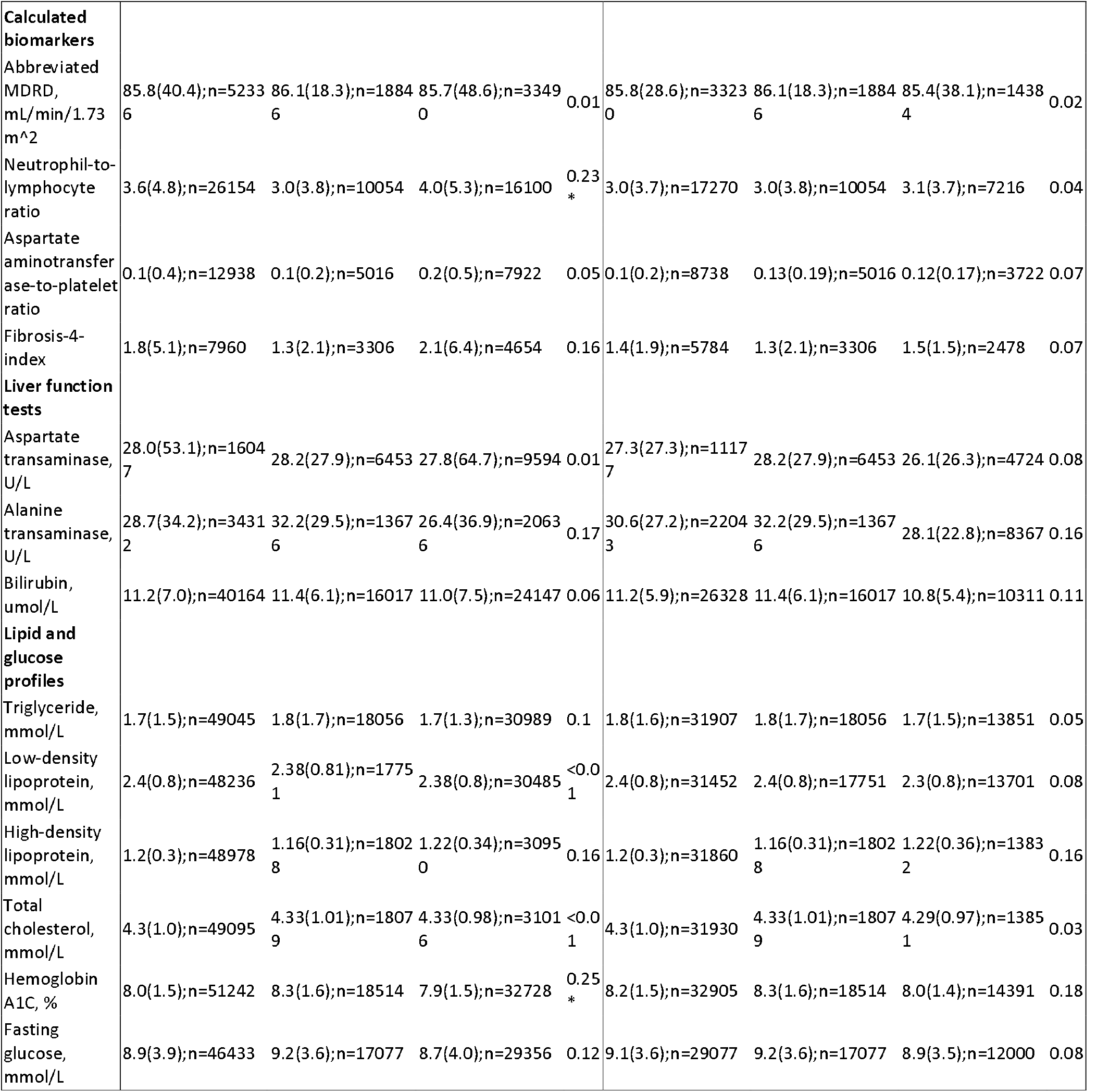
Baseline and clinical characteristics of patients with SGLT2I v.s. DPP4I use before and after propensity score matching (1:1). ^*^for SMD>0.2; SD: standard deviation; SGLT2I: sodium glucose cotransporter-2 inhibitor; DPP4I: dipeptidyl peptidase-4 inhibitor; ACEI: Angiotensin-converting enzyme inhibitors; ARBs: angiotensin receptor blockers; MDRD: modification of diet in renal disease; NAFLD: Non-alcoholic fatty liver disease; # indicated the difference between SGLT2I users and DPP4I users.

Over a total follow-up of 236856.5 person-years, the incidence rate (IR) of NAFLD was lower amongst SGLT2I users (IR: 3.4; 95% CI: 3.0-3.7) compared to DPP4I (IR: 5.9; 95% CI: 5.5-6.4) after propensity score matching **(Table 2)**. Meanwhile, after a follow-up of 239941.9 person-year, the incidence of HCC was lower amongst SGLT2I users (IR: 0.3; 95% CI: 0.2-0.4) compared to DPP4I users (IR: 1.3; 95% CI: 1.1-1.5). SGLT2I users (IR: 1.1; 95% CI: 0.9-1.3) also had a lower incidence of cancer-related mortality than DPP4I users (IR: 6.6; 95% CI: 6.1-7.1); the incidence of all-cause mortality was also lower amongst SGLT2I users (IR: 5.0; 95% CI: 4.6-5.4) than DPP4I users (IR: 24.4; 95% CI: 23.6-25.4).

**Table 2.** Annualized incidence rate of primary and secondary outcomes in patient cohort after 1:1 propensity score matching. NAFLD: Non-alcoholic fatty liver disease; IR: Incidence rate; SGLT2I: sodium glucose cotransporter-2 inhibitor; DPP4I: dipeptidyl peptidase-4 inhibitor

| NAFLD |  |  |  | HCC |  |  |  |
| --- | --- | --- | --- | --- | --- | --- | --- |
| DPP4I users | Person-year | Number of events | IR[95% CI] | DPP4I users | Person-year | Number of events | IR[95% CI] |
| Total | 115096.9 | 680 | 5.9[5.5-6.4] | Total | 116930.9 | 151 | 1.3[1.1-1.5] |
| Year 1 | 22059.8 | 160 | 7.3[6.2-8.5] | Year 1 | 22143.8 | 29 | 1.3[0.9-1.9] |
| Year 2 | 21857.9 | 107 | 4.9[4.1-5.9] | Year 2 | 22062.8 | 46 | 2.1[1.6-2.8] |
| Year 3 | 21269.7 | 245 | 11.5[10.2-13.1] | Year 3 | 21649.6 | 69 | 3.4[2.7-4/3] |
| Year 4 | 20020.2 | 124 | 6.2[5.2-7.4] | Year 4 | 20460.8 | 7 | 0.4[0.2-0.7] |
| Year 5 | 19180.3 | 44 | 2.3[1.7-3.1] | Year 5 | 19623.0 | 0 | 0 |
| Year 6 or above | 10709.0 | 0 | 0 | Year 6 or above | 10990.9 | 0 | 0 |
| SGLT2I users | Person-year | Number of events | IR[95% CI] | SGLT2I users | Person-year | Number of events | IR[95% CI] |
| Total | 121759.6 | 410 | 3.4[3.0-3.7] | Total | 123011 | 36 | 0.3[0.2-0.4] |
| Year 1 | 22084.9 | 111 | 2.8[2.1-3.7] | Year 1 | 22154.0 | 0 | - |
| Year 2 | 22004.9 | 62 | 2.2[1.6-3.1] | Year 2 | 22141.6 | 5 | 0.2[0.1-0.5] |
| Year 3 | 21879.9 | 72 | 2.2[1.6-3.1] | Year 3 | 22081.0 | 7 | 0.3[0.2-0.7] |
| Year 4 | 21698.7 | 91 | 3.7[2.8-4.7] | Year 4 | 21965.2 | 11 | 0.5[0.3-0.9] |
| Year 5 | 21391.1 | 74 | 2.1[1.5-2.7] | Year 5 | 21741.5 | 11 | 0.5[0.3-0.9] |
| Year 6 or above | 12700.1 | 0 | 0 | Year 6 or above | 12927.7 | 2 | 0.2[0-0.6] |
| **Overall** | **236856.5** | **1090** | **4.6[4.3-4.9]** | **Overall** | **239941.9** | **187** | **0.8[0.7-0.9]** |
| Cancer-caused mortality |  |  |  | All-cause mortality |  |  |  |
| DPP4I users | Person-year | Number of events | IR[95% CI] | DPP4I users | Person-year | Number of events | IR[95% CI] |
| Total | 117247.7 | 771 | 6.6[6.1-7.1] | Total | 117247.7 | 2866 | 24.4[23.6-25.4] |
| Year 1 | 22143.8 | 1 | 0[0-0.3] | Year 1 | 22143.8 | 43 | 1.9[0.4-2.6] |
| Year 2 | 22073.7 | 64 | 2.9[2.3-3.7] | Year 2 | 22073.7 | 143 | 6.5[5.5-7.6] |
| Year 3 | 21678.3 | 259 | 11.9[10.6-13.5] | Year 3 | 21678.3 | 694 | 32.0[29.7-34.5] |
| Year 4 | 20553.7 | 336 | 16.3[14.7-18.2] | Year 4 | 20553.7 | 1230 | 59.8[56.6-63.3] |
| Year 5 | 19751.6 | 105 | 5.3[4.4-6.4] | Year 5 | 19751.6 | 636 | 32.2[29.8-34.8] |
| Year 6 or above | 11046.6 | 6 | 0.5[0.2-1.2] | Year 6 or above | 11046.6 | 120 | 10.9[9.1-13.0] |
| SGLT2I users | Person-year | Number of events | IR[95% CI] | SGLT2I users | Person-year | Number of events | IR[95% CI] |
| Total | 123072.9 | 130 | 1.1[0.9-1.3] | Total | 123072.9 | 613 | 5.0[4.6-5.4] |
| Year 1 | 22154.0 | 0 | 0 | Year 1 | 22154.0 | 0 | - |
| Year 2 | 22143.8 | 5 | 0.2[0.1-0.5] | Year 2 | 22143.8 | 33 | 1.5[1.1-2.1] |
| Year 3 | 22089.4 | 16 | 0.7[0.4-1.2] | Year 3 | 22089.4 | 78 | 3.5[2.8-4.4] |
| Year 4 | 21978.7 | 36 | 1.6[1.2-2.3] | Year 4 | 21978.7 | 151 | 6.9[5.9-8.1] |
| Year 5 | 21764.2 | 58 | 2.7[2.1-3.4] | Year 5 | 21764.2 | 262 | 12.2[10.7-13.6] |
| **Year 6 or above** | 12942.8 | 15 | 1.2[0.7-1.9] | Year 6 or above | 12942.8 | 89 | 6.9[5.6-8.5] |
| **Overall** | 240320.6 | 901 | 3.7[3.5-4] | **Overall** | 240320.6 | 3479 | 14.5[14.0-15.0] |

### Significant predictors of the study outcomes

Univariable Cox regression identified the significant risk factors for NAFLD and HCC before and after propensity score matching (1:1) **(Supplementary Table 5)**. In the multivariable Cox models, SGLT2I was associated with lower risks of NAFLD (Hazard ratio [HR]: 0.39; 95% CI: 0.34-0.46) and HCC (HR: 0.46; 95% CI: 0.29-0.72) after adjustments for significant demographics, past comorbidities, non-SGLT2I/DPP4I medications, abbreviated MDRD, fasting glucose, HbA1c, and duration from earliest diabetes mellitus date to initial drug exposure date. SGLT2I was also associated with lower risks of cancer-related mortality (HR: 0.29; 95% CI: 0.23-0.37) and all-cause mortality (HR: 0.28; 95% CI: 0.25-0.31) upon adjustments. The cumulative incidence curves stratified by SGLT2I versus DPP4I demonstrated that SGLT2I was associated with a lower cumulative hazard for NAFLD, HCC, cancer-related mortality and all-cause mortality after matching **(Figure 2)**.

**Figure 2.**
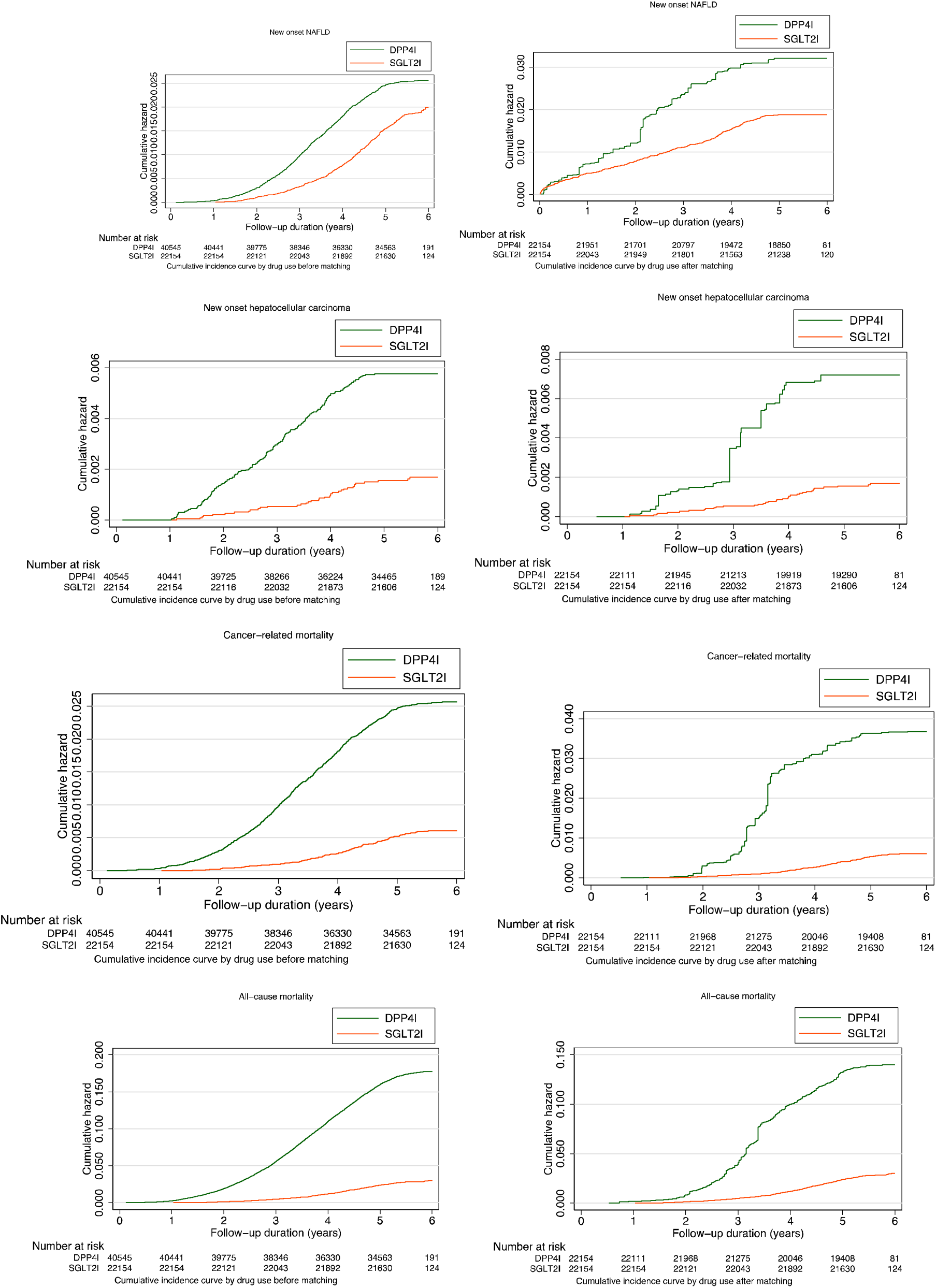
Cumulative incidence curves for all-cause mortality, cancer-related mortality, new onset HCC, new onset NAFLD stratified by drug exposure of SGLT2I and DPP4I before and after propensity score matching (1:1)

### Subgroup analysis and sensitivity analyses

In the subgroup analysis, SGLT2I was associated lower cumulative incidence of NAFLD and HCC regardless of gender **(Figure 3 and Supplementary Figure 2)**. SGLT2I was associated with reduced risks of NAFLD compared to DPP4I particularly amongst patients younger than 65 years old regardless of history of hypertension and ischaemic heart diseases. SGLT2I was also associated with lower risks of HCC amongst patients older than 65 years old, without history of hypertension or ischaemic diseases. Amongst HBV negative patients, SGLT2I was associated with lower risks of NAFLD (HR: 0.35; 95% CI: 0.30-0.42), HCC (HR: 0.30; 95% CI: 0.18-0.50), cancer-related mortality (HR: 0.32; 95% CI: 0.25-0.41), and all-cause mortality (HR: 0.30; 95% CI: 0.27-0.34). Meanwhile, amongst HBV-positive patients, SGLT2I was associated with an insignificant risk of NAFLD (HR: 1.21; 95% CI: 0.69-2.13) and higher risks of HCC (HR: 3.28; 95% CI: 1.21-8.90) compared to DPP4I **(Supplementary Table 6)**. However, SGLT2I was associated with lower risk of cancer-related mortality (HR: 0.08; 95% CI: 0.05-0.19) and all-cause mortality (HR: 0.09; 95% CI: 0.04-0.12) among those patients.

**Figure 3A.**
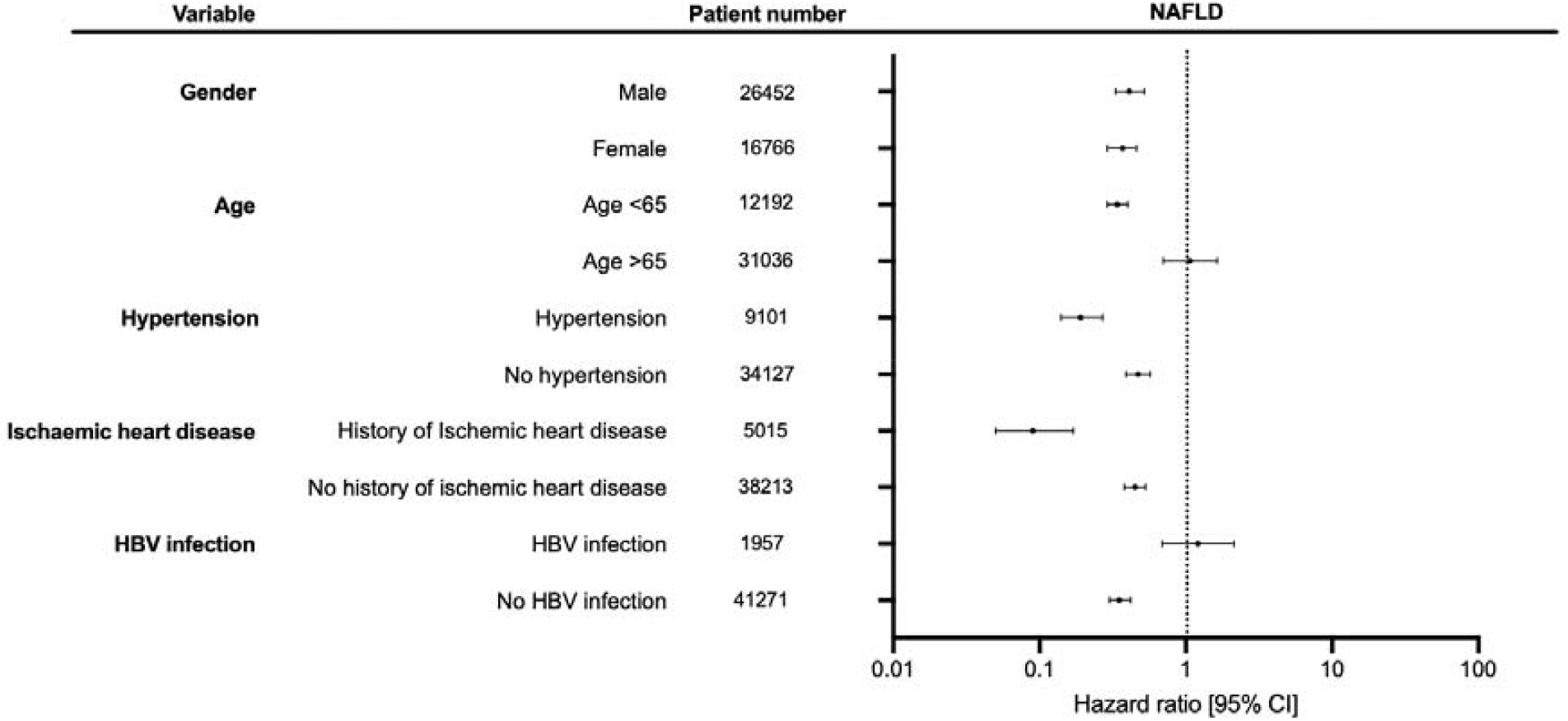
Subgroup analyses for SGLT2I v.s. DPP4I exposure predict new onset NAFLD in the matched cohort. SGLT2I: Sodium-glucose cotransporter-2 inhibitors; DPP4I: Dipeptidyl peptidase-4 inhibitors.

**Figure 3B.**
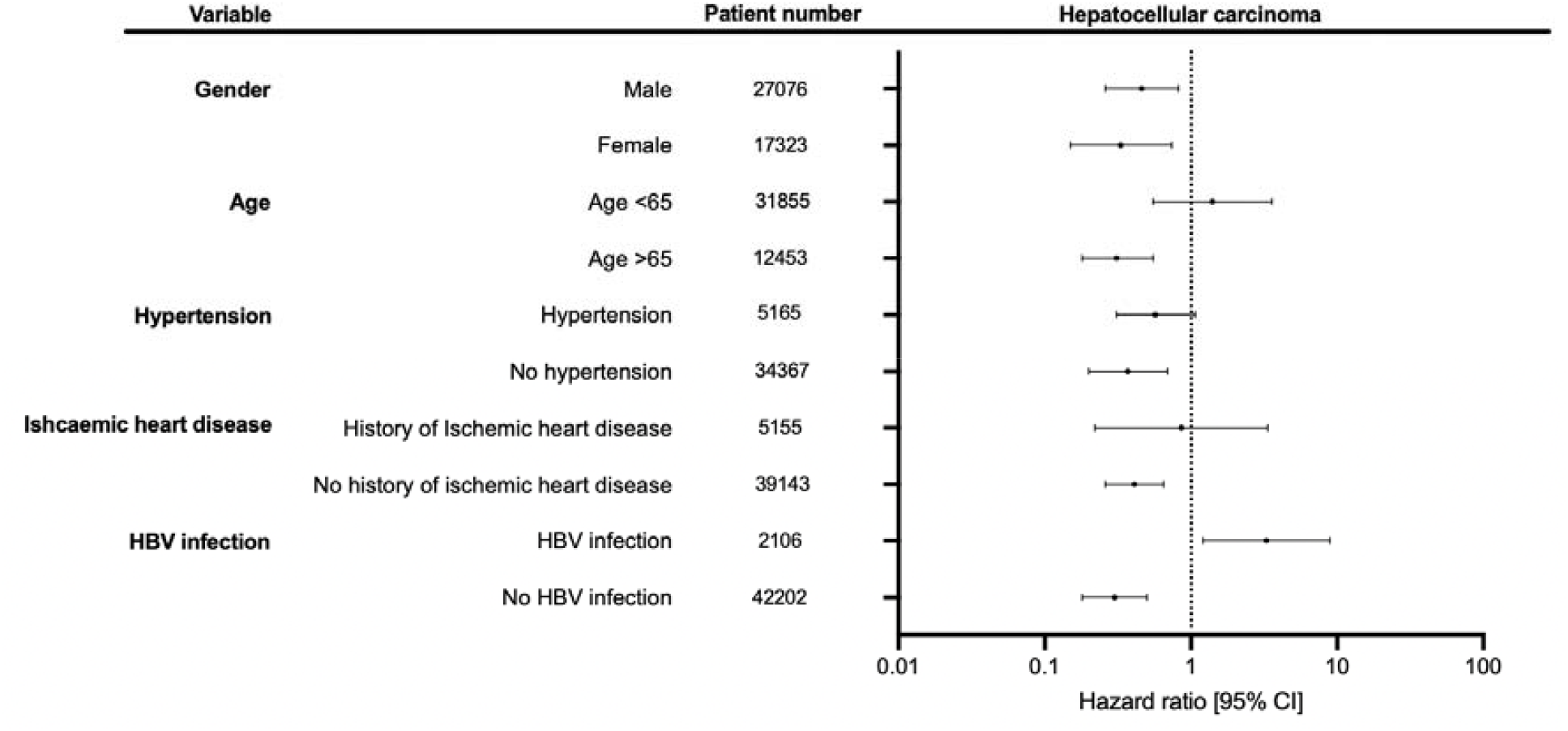
Subgroup analyses for SGLT2I v.s. DPP4I exposure predict new onset HCC in the matched cohort. SGLT2I: Sodium-glucose cotransporter-2 inhibitors; DPP4I: Dipeptidyl peptidase-4 inhibitors.

The interaction effects with abbreviated MDRD demonstrated that SGLT2I was associated with lower risks of NAFLD, HCC, cancer-related mortality, and all-cause mortality across different abbreviated MDRD **(Supplementary Figure 3A)**. Meanwhile, as the neutrophil-to-lymphocyte ratio was lowered, the risks for NAFLD increased for DPP4I users while the risks for HCC, cancer-related mortality and all-cause mortality decreased (**Supplementary Figure 3B)**. The SGLT2I was also associated with lower risks of NAFLD, HCC, cancer-related mortality, and all-cause mortality across all aspartate aminotransferase-to-platelet ratio and fibrosis-4 index **(Supplementary Figure 3C and 3D)**.

Sensitivity analyses were performed to confirm the predictive ability of the models. SGLT2I was associated with lower risks of NAFLD (HR: 0.45; 95% CI: 0.35-0.64) and HCC (HR: 0.42; 95% CI: 0.31-0.54) compared to DPP4I in the cause-specific hazard. SGLT2I was also associated with lower risks of NAFLD (HR: 0.56; 95% CI: 0.39-0.82) and HCC (HR: 0.53; 95% CI: 0.41-0.75) compared to DPP4I in the subdistribution model **(Table 4)**. SGLT2I also was associated with lower risks of new-onset NAFLD, HCC, cancer-related mortality, and all-cause mortality after different propensity score approaches (all P-values <0.0001).

**Table 3.** Multivariate Cox regression models with adjustments to predict primary and secondary outcomes in the matched cohort. *for p≤ 0.05, ** for p ≤ 0.01, *** for p ≤ 0.001; HR: hazard ratio; CI: confidence interval; SGLT2I: sodium glucose cotransporter-2 inhibitor; DPP4I: dipeptidyl peptidase-4 inhibitor; Model 1 adjusted for significant demographics. Model 2 adjusted for significant demographics, and past comorbidities. Model 3 adjusted for significant demographics, past comorbidities, and non-SGLT2I/DPP4I medications. Model 4 adjusted for significant demographics, past comorbidities, non-SGLT2I/DPP4I medications, abbreviated MDRD, fasting glucose, HbA1c, and duration from earliest diabetes mellitus date to initial drug exposure date. ^ Patients with prior NAFLD were excluded in the multivariate Cox regression models to predict new onset NAFLD.

|  |  | **Model 1** | **Model 2** | **Model 3** | **Model 4** |
| --- | --- | --- | --- | --- | --- |
| **SGLT2I**<br>**v.s.**<br>**DPP4I** | **New onset NAFLD**<br>**HR [95% CI];P**<br>**value<sup>^</sup>** | 0.48[0.42-0.55];<br><0.0001*** | 0.46[0.40-0.53];<br><0.0001*** | 0.45[0.39-0.52];<br><0.0001*** | 0.39[0.34-0.46];<br><0.0001*** |
|  | **New onset HCC**<br>**HR [95% CI];P**<br>**value** | 0.24[0.17-0.35];<br><0.0001*** | 0.23[0.16-0.34];<br><0.0001*** | 0.26[0.18-0.39];<br><0.0001*** | 0.46[0.29-0.72];<br>0.0007*** |
|  | **New onset cancer**<br>**related mortality**<br>**HR [95% CI];P**<br>**value** | 0.17[0.14-0.20];<br><0.0001*** | 0.17[0.14-0.21];<br><0.0001*** | 0.19[0.15-0.22];<br><0.0001*** | 0.29[0.23-0.37];<br><0.0001*** |
|  | **New onset all-cause**<br>**mortality**<br>**HR [95% CI];P**<br>**value** | 0.21[0.19-0.23];<br><0.0001*** | 0.21[0.19-0.23];<br><0.0001*** | 0.23[0.21-0.25];<br><0.0001*** | 0.28[0.25-0.31];<br><0.0001*** |

**Table 4.** Sensitivity analyses for SGLT2I v.s. DPP4I exposure predict new onset NAFLD and new onset HCC in the matched cohort. *for p≤ 0.05, ** for p ≤ 0.01, *** for p ≤ 0.001; SGLT2I: Sodium-glucose cotransporter-2 inhibitors; DPP4I: Dipeptidyl peptidase-4 inhibitors; HR: hazard ratio; CI: confidence interval; PS: propensity score; IPTW: inverse probability of treatment weighting, SIPTW: stable inverse probability of treatment weighting.

| Model | New onset NAFLD<br>HR [95% CI] | New onset HCC<br>HR [95% CI] | New onset cancer-related mortality<br>HR [95% CI];P<br>value | All-cause mortality<br>HR [95% CI];P<br>value |
| --- | --- | --- | --- | --- |
| Cause-specific<br>hazard models | 0.45[0.37-0.62];<0.0001*** | 0.42[0.31-0.57];<0.0001*** | 0.52[0.33-0.7];<0.0001*** | 0.33[0.13-0.35];<0.0001*** |
| Sub-<br>distribution<br>hazard models | 0.59[0.38-0.83];<0.0001*** | 0.54[0.44-0.76];<0.0001*** | 0.26[0.21-0.52];<0.0001*** | 0.22[0.2-0.54];<0.0001*** |
| PS<br>stratification | 0.49[0.2-0.83];<0.0001*** | 0.38[0.32-0.69];<0.0001*** | 0.28[0.21-0.72];<0.0001*** | 0.41[0.29-0.64];<0.0001*** |
| PS with IPTW | 0.5[0.37-0.77];<0.0001*** | 0.39[0.35-0.53];<0.0001*** | 0.35[0.29-0.6];<0.0001*** | 0.26[0.19-0.44];<0.0001*** |
| PS with<br>SIPTW | 0.42[0.35-0.61];<0.0001*** | 0.33[0.18-0.52];<0.0001*** | 0.36[0.23-0.57];<0.0001*** | 0.35[0.23-0.71];<0.0001*** |

## Discussion

In this territory-wide retrospective cohort study, we used real-world data from routine clinical practice to compare the association between SGLT2I versus DPP4I and NAFLD and HCC. Our findings demonstrated that SGLT2I was associated with 58% lower risk of NAFLD and 55% lower risk of HCC than DPP4I users. However, amongst patients infected with HBV, DPP4I was associated with lower risks of HCC compared to SGLT2I.

### Comparison with previous studies

T2DM is a metabolic syndrome characterized by hyperinsulinemia and contributes as a comorbidity in NAFLD, subsequently increasing the risk of HCC development [27, 28]. The prevalence of NAFLD in T2DM patients ranges from 29.6% to 87.1% [29]. T2DM patients have a 2.5-fold increased risk of developing HCC, while NAFLD further increases the risks [30, 31]. As the prevalence of NAFLD continues to be on the rise, managing diabetes becomes more pressing considering the risks of developing HCC. Antidiabetic agents such as metformin, thiazolidinediones, and GLP-1 analogues have been shown to improve the pathological manifestations of NAFLD and HCC in T2DM [32, 33].

The protective effects of SGLT2I on the cardiovascular system are well-established [34-37]. For hepatic diseases, the literature generally supports the notion that SGLT2I and DPP4I are beneficial in hepatic diseases but lack direct comparisons. The results from our study suggest that SGLT2I may have a lower risk of NAFLD and HCC development compared to DPP4I. Furthermore, the reduction in NAFLD and hepatocellular carcinoma might be one of the contributing factors to the reduction in all-cause mortality by reducing cancer-related mortality.

It was suggested that a significant proportion of HCC developed amongst patients without cirrhosis [38]. In our results, SGLT2I reduced the risks of NAFLD and HCC risks across all baseline severity of liver fibrosis and cirrhosis, as indirectly reflected by the aspartate aminotransferase-to-platelet ratio and fibrosis-4 index interaction. Multiple studies have found that SGLT2I lowered the risk of NAFLD with pathological reductions of steatosis on histological examinations [13, 39-41]. In one study, SGLT2I reduced the risk for death and improved the survival of T2DM veterans with cirrhosis when compared to DPP4I [42]. Furthermore, SGLT2I also reduced the body mass index significantly and reduced the hepatocarcinogenesis for NAFLD [43]. On the other hand, while DPP4I studies have also been shown to reduce the risk of HCC in T2DM patients with chronic HCV infection [16, 44], data assessing the association with the risk of NAFLD are mixed [45-47]. A systematic review demonstrated no hepatic benefit associated with DPP4I in patients with hepatic steatosis but significant risk reductions in patients receiving SGLT2I [48].

Chronic HBV infection is still prevalent in Asia despite universal vaccination for individuals over 20 years [38]. The co-presence of steatohepatitis and HBV infection significantly increased the risks of HCC and death [49]. While SGLT2I lowered the risks of HCC amongst patients without HBV infection, SGLT2I was associated with higher risks of HCC (HR: 3.28; 95% CI: 1.21-8.90) compared to DPP4I amongst patients infected with HBV **(Supplementary Table 6)**. While HBV infection may contribute to more severe fibrosis, SGLT2I was associated with lower risks of HCC across all baseline severity of liver fibrosis. However, the effect can be due to dipeptidyl peptidase-4 (DPP4). DPP4 is an important molecule involved in the development of HCC, such that inhibition of DPP4 may help prevent HCC amongst HCV-infected patients. Indeed, the serum DPP4 level was previously suggested to be elevated amongst viral hepatitis patients [50]. However, the underlying mechanism mediating this finding in patients with T2DM and coinfection with HBV remains unclear. Future research is needed.

### Potential underlying mechanisms

Several mechanisms have been proposed to explain the relationship between SGLT2I and NAFLD and HCC. It was hypothesized that SGLT2I inhibited de novo lipogenesis by inhibiting the expression of the FAS gene involved in fatty acid biosynthesis, which decreases fatty acid production and reduces steatosis [51]. Besides, SGLT2I has demonstrated anti-inflammatory and anti-steatosis properties preventing the progression of NAFLD to HCC [52]. Furthermore, SGLT2 receptors are highly expressed in liver tumours due to their increased demand for glucose for ATP synthesis and overall growth [53]. Meanwhile, DPP4I was also suggested to reduce the risks of HCC via several mechanisms. It was previously suggested that the GLP-1 hormone might help ameliorate liver fat accumulation and prevent the progression of NAFLD [54]. It also reduces HCC through the activation of lymphocyte chemotaxis and downregulation of the pentose phosphate pathway [55, 56]. However, there are also some conflicting results which suggest DPP4I may play a role in the progression of non-alcoholic steatohepatitis-related HCC by suppressing p62 and Keap1 [22]. Future research is needed to confirm the effects of SGLT2I and DPP4I in HCC.

### Clinical implications

Given the importance of NAFLD and HCC in T2DM, [57], there is a need to investigate how SGLT2I and DPP4I may modify the risks for these diseases. The present study used data from routine clinical practice, which may influence the choice of second-line antidiabetic therapy in T2DM patients in terms of the hepatic disease risks. The findings of our study show that SGLTI and DPP4I may help prevent NAFLD and HCC compared to DPP4I. It particularly reduced the risks of NAFLD regardless of gender, prior history of hypertension and ischaemic heart diseases. However, amongst HBV-positive patients, DPP4I may be a better option to prevent the new-onset HCC. Generally, SGLT2I were found to reduce the risk of malignancies. SGLT2I patient groups had a lower risk of haematological and urinary tract malignancies in a nationwide study conducted by *Rokszin et al*. [48]. By exploring the association of SGLT2I and DPP4I in HCC, we add to the growing body of evidence supporting the use of antidiabetic agents in preventing NAFLD and potentially HCC. Further investigations are needed to confirm the causation relationship between SGLT2I and DPP4I with HCC, especially amongst HBV-positive patients.

### Limitations

This study had several limitations. Firstly, given the observational nature of this study, there is inherent under-coding, coding errors, and missing data leading to information bias. The retrospective design necessitates the presentation of associations but not causal links between SGLT2I/DPP4I use and the risk of new-onset NAFLD and HCC. Secondly, medication adherence can only be assessed indirectly through prescription refills but not through direct measurement of drug exposure. Thirdly, residual and post-baseline confounding may be present despite robust propensity-matching, particularly with the unavailability of information on NAFLD and HCC risk factors such as smoking, obesity, and the potential overlooked alcohol consumption. As such, lipid profile was included in an attempt to account for obesity in an indirect way. Last but not least, the duration of drug exposure has not been controlled, which may affect their risk against the study outcomes. Time-varying analysis was also not included in this study.

## Conclusion

SGLT2I was associated with lower risks of NAFLD and HCC compared to DPP4I after propensity scores matching and adjustments. SGLT2I was also associated with lower risks of cancer-related mortality and all-cause mortality compared to DPP4I. However, amongst patients infected with HBV, DPP4I was associated with lower risks of HCC compared to SGLT2I. The results supported the need for further evaluation in the prospective setting.

## Data Availability

An anonymised version without identifiable or personal information is available from the corresponding authors upon reasonable request for research purposes.

## Acknowledgements

None.

