## Supplementary Table 1 for "Lower risks of sodium glucose cotransporter 2 (SGLT2) inhibitors compared to dipeptidyl peptidase-4 (DPP4) inhibitors for new-onset non-alcoholic fatty liver disease and hepatocellular carcinoma in type 2 diabetes mellitus: A population-based study"

**Supplementary Appendix**

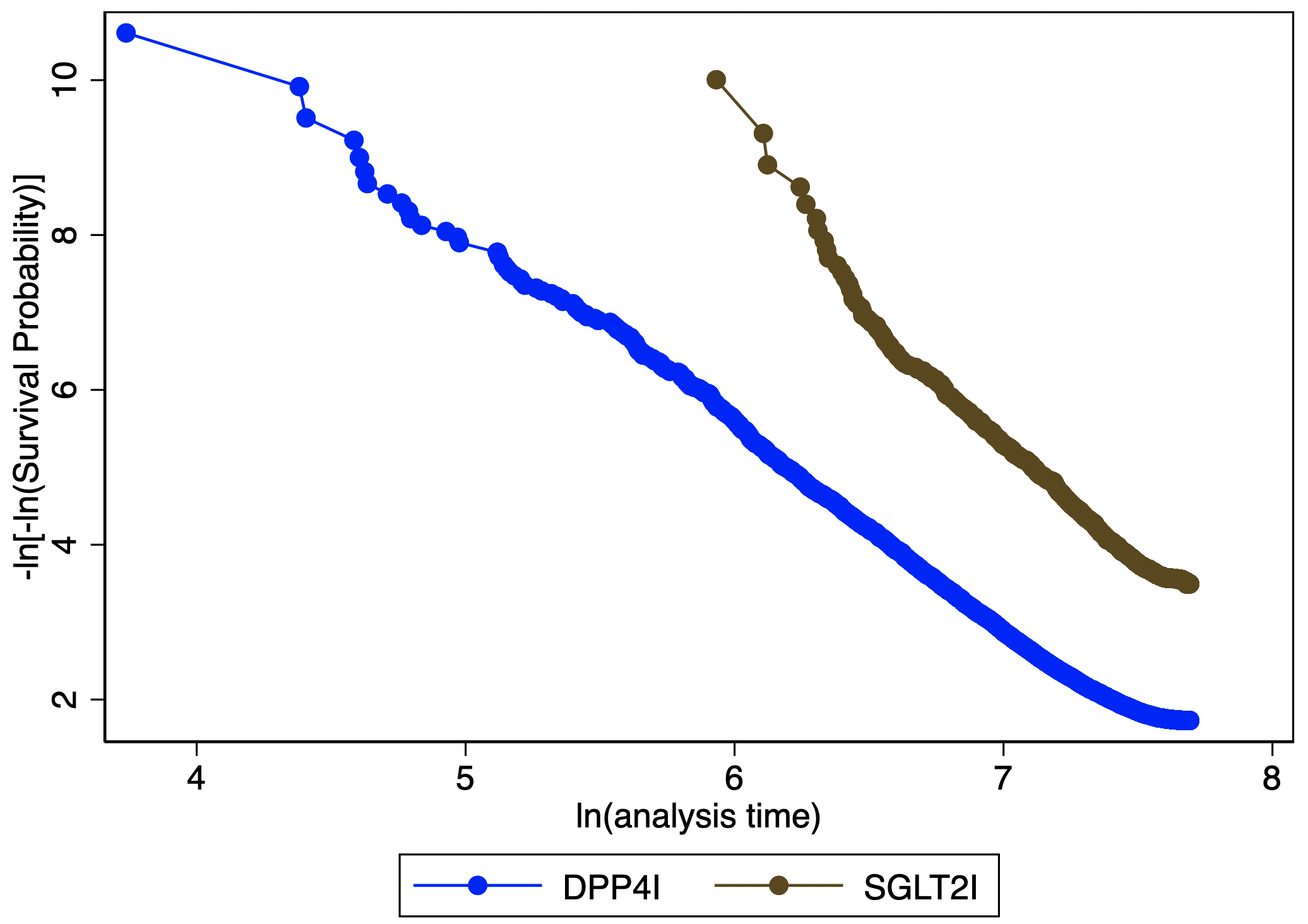

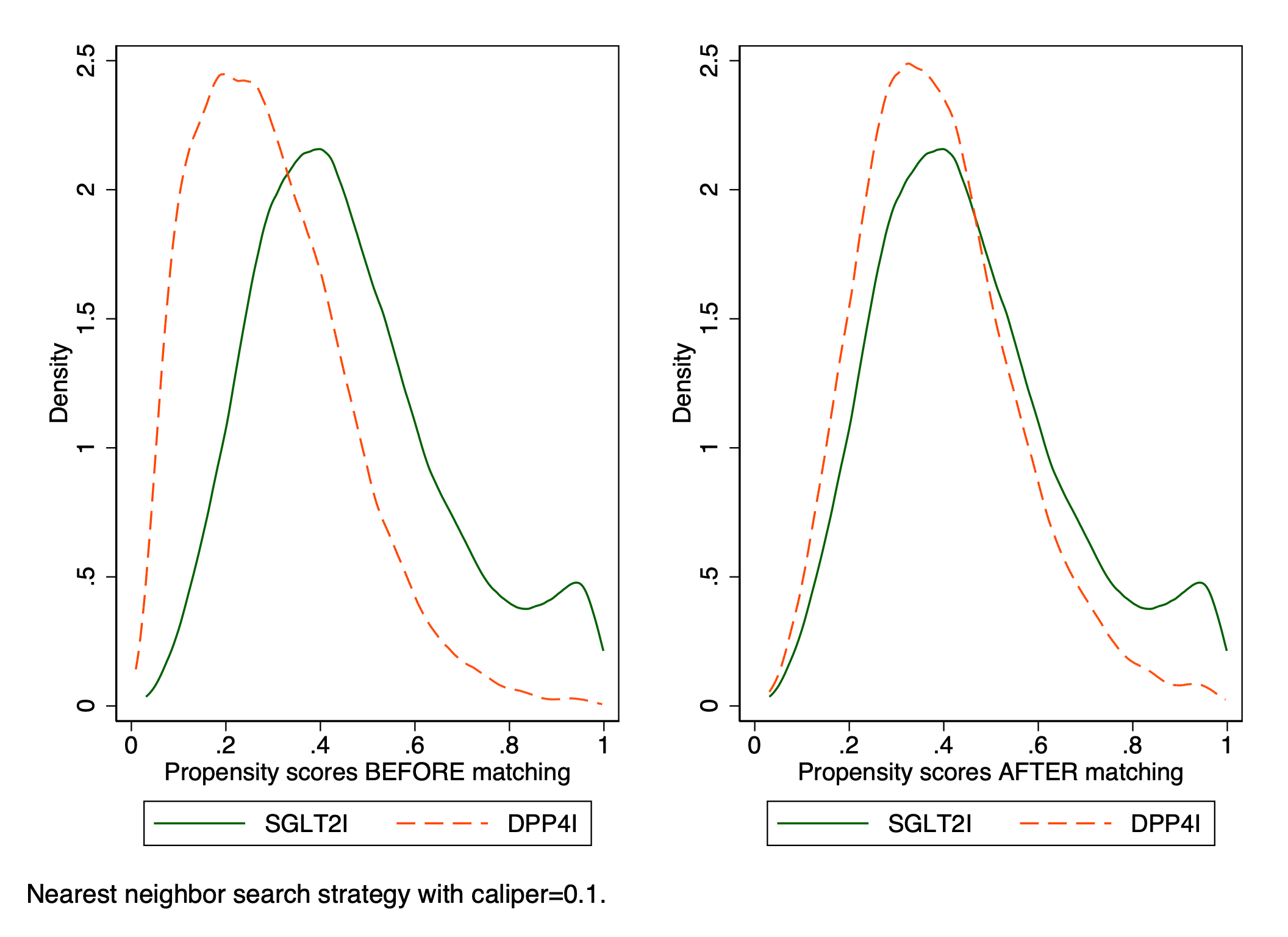

**Supplementary Figure 1. Propensity score matching comparisons and proportional hazard assumption checking with parallel lines for SGLT2I v.s. DPP4I before and after 1:1 matching with nearest neighbor search strategy with caliper of 0.1**

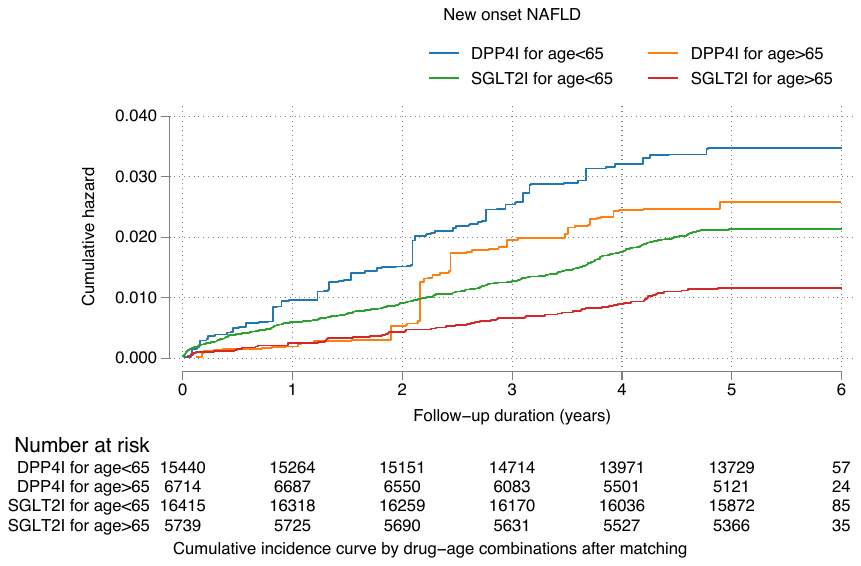

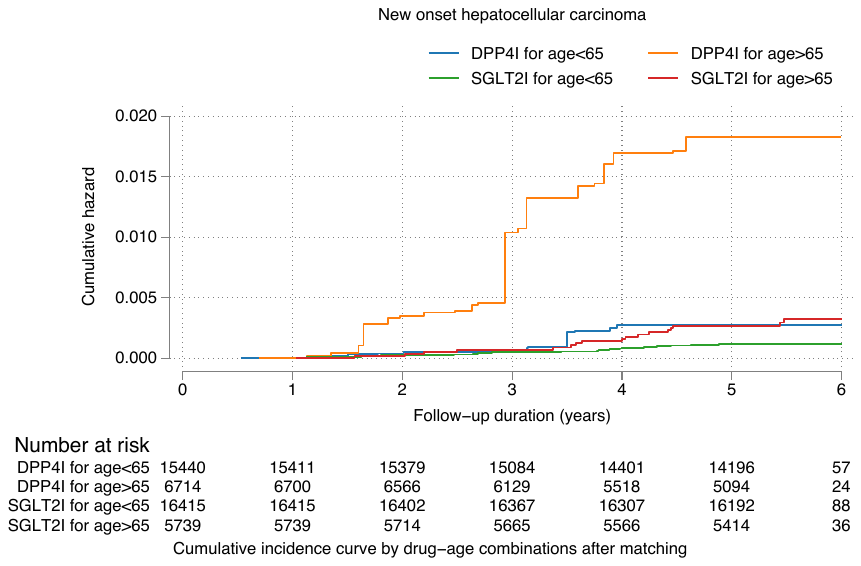

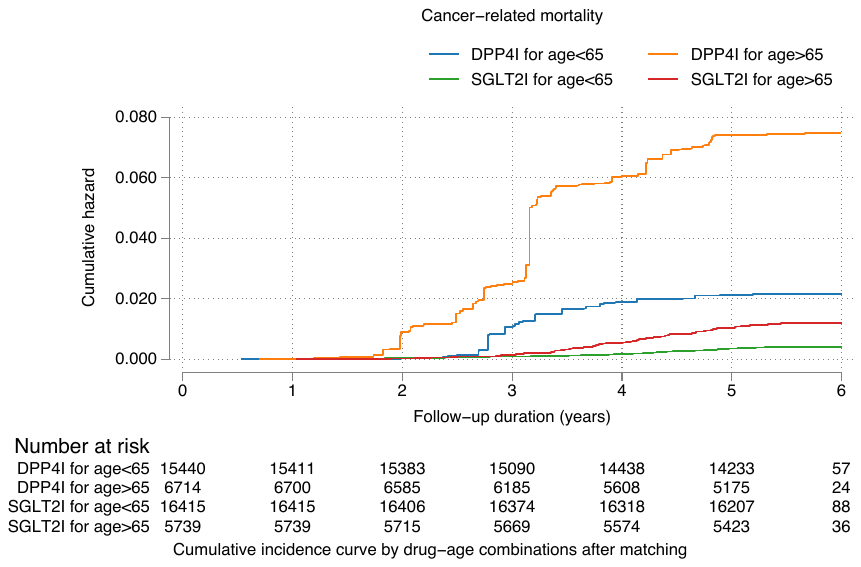

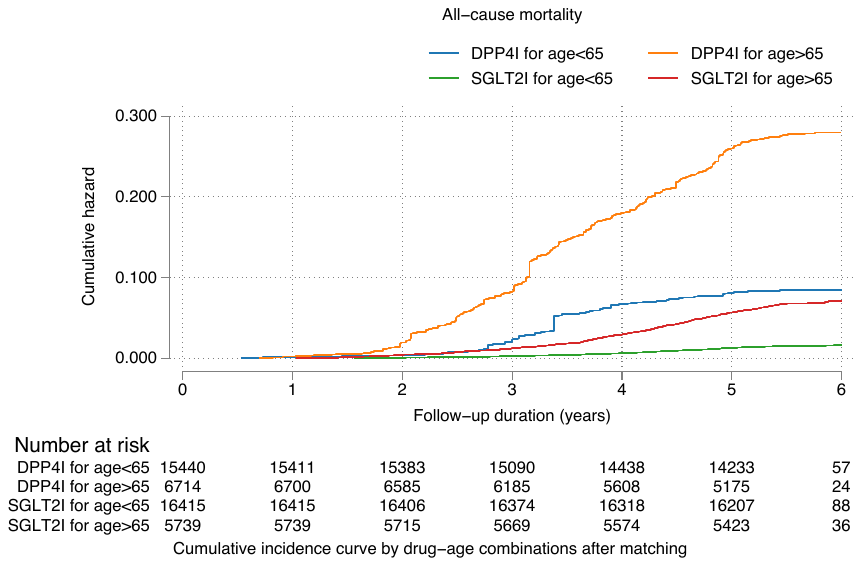

**Supplementary Figure 2A. Subgroup analysis: Cumulative incidence curves for all-cause mortality, cancer-related mortality, new onset hepatocellular carcinoma, new onset NAFLD stratified by drug-age combinations and the effects of SGLT2I and DPP4I in the matched cohort (1:1)**

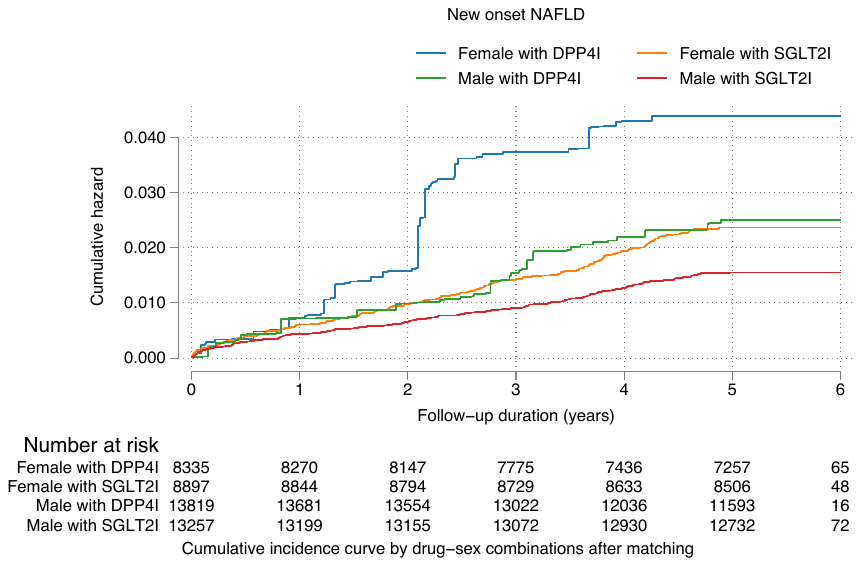

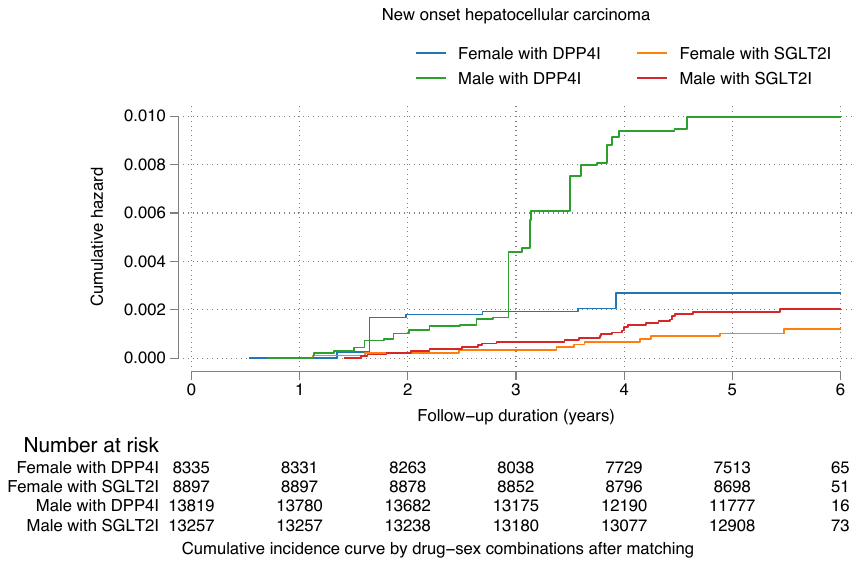

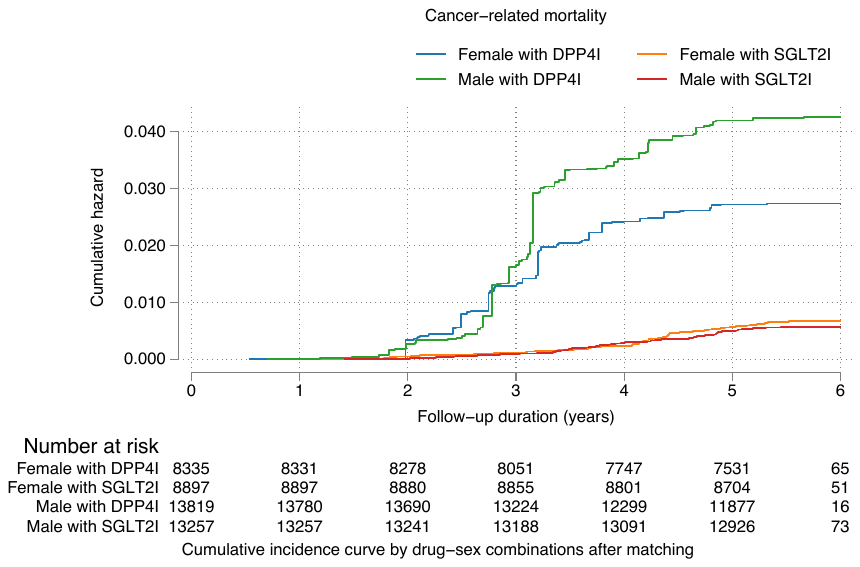

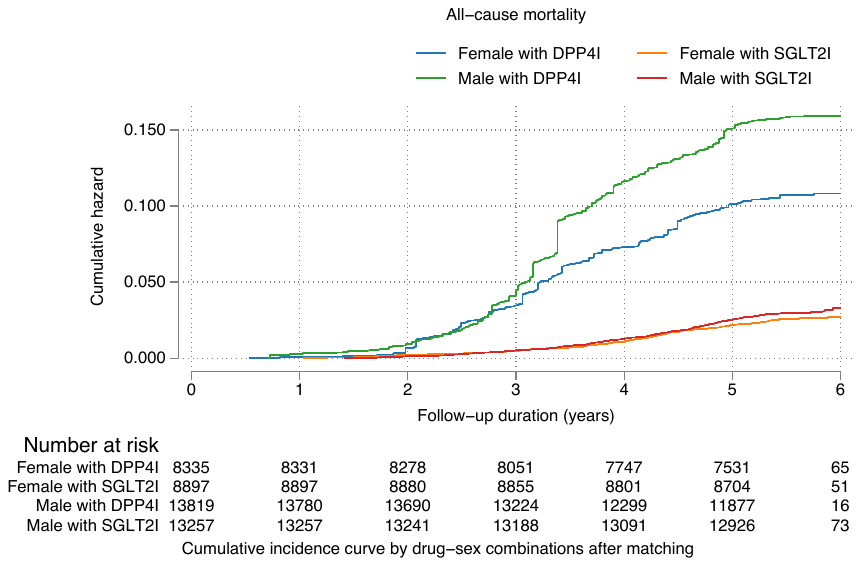

**Supplementary Figure 2B. Subgroup analysis: Cumulative incidence curves for all-cause mortality, cancer-related mortality, new onset hepatocellular carcinoma, new onset NAFLD stratified by drug-sex combinations and the effects of SGLT2I and DPP4I in the matched cohort (1:1)**

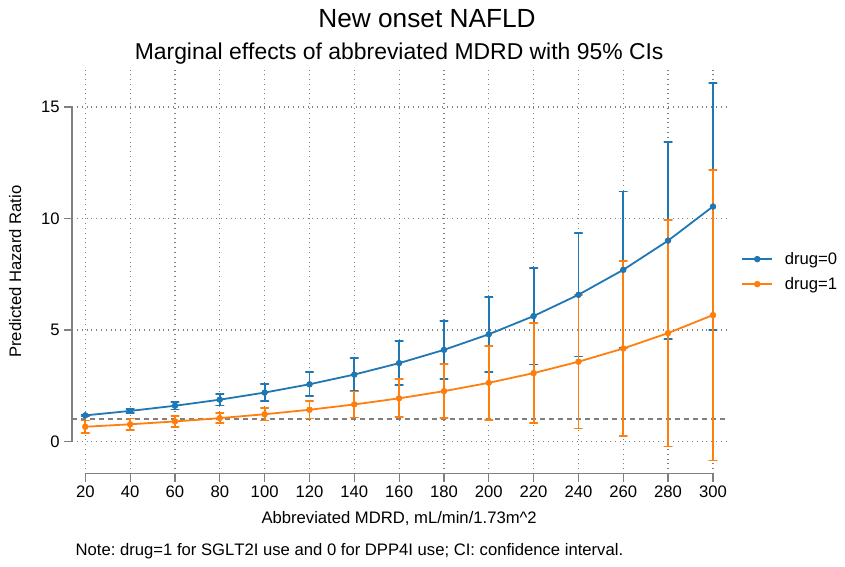

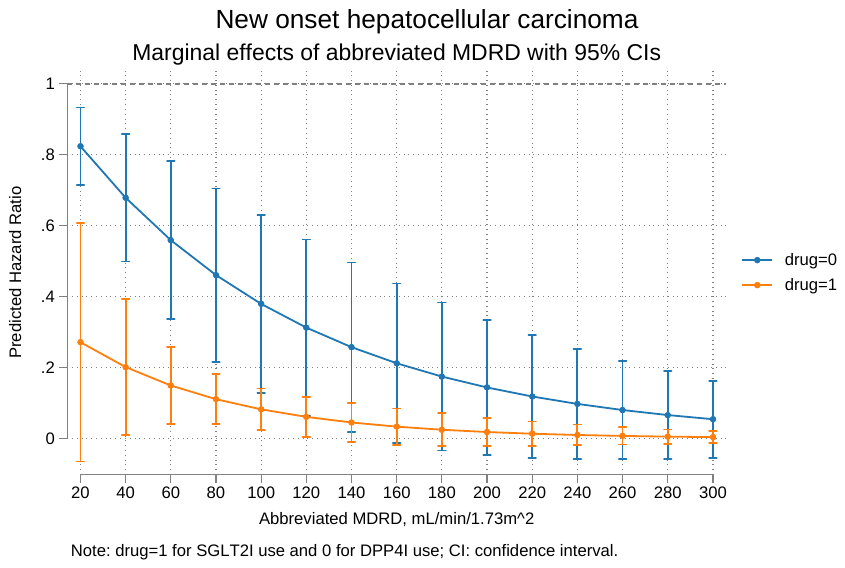

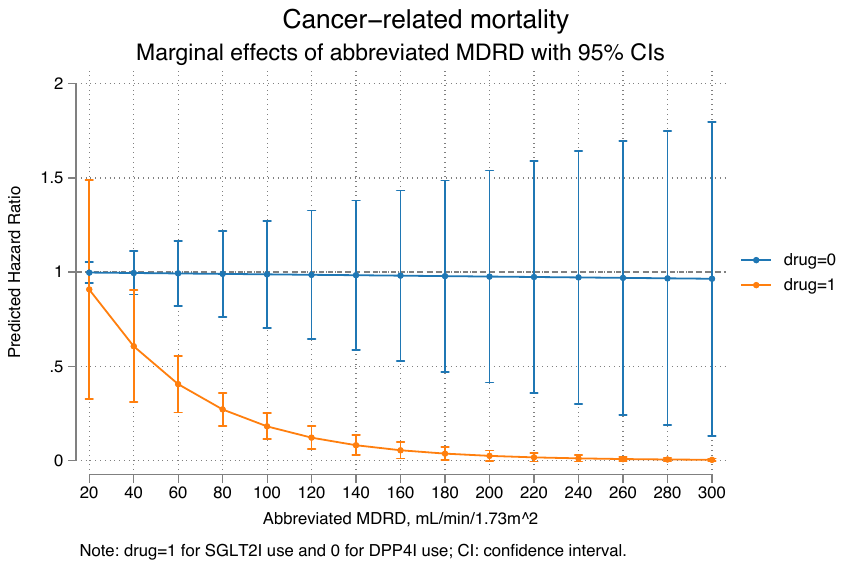

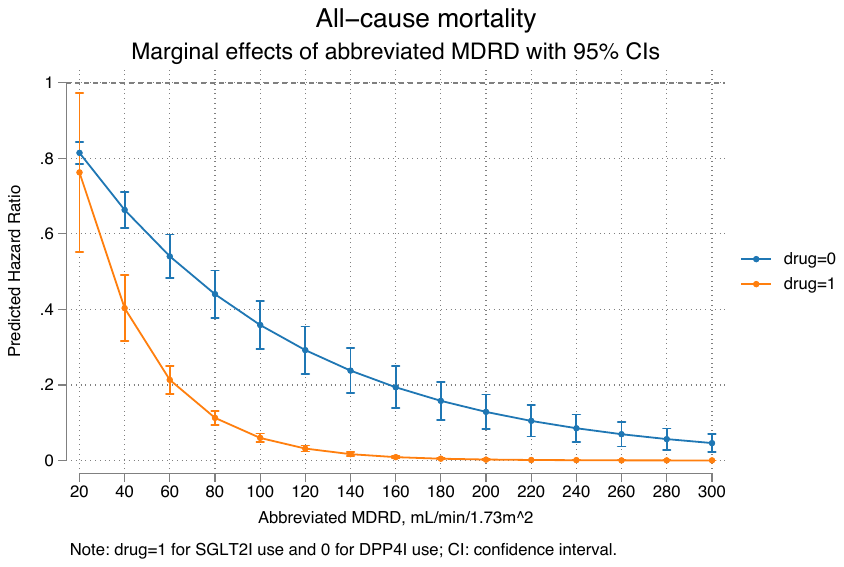

**Supplementary Figure 3A. Interaction effects of MDRD and drug exposure of SGLT2I and DPP4I for all-cause mortality, cancer-related mortality, new onset hepatocellular carcinoma, new onset NAFLD in the matched cohort (1:1)**

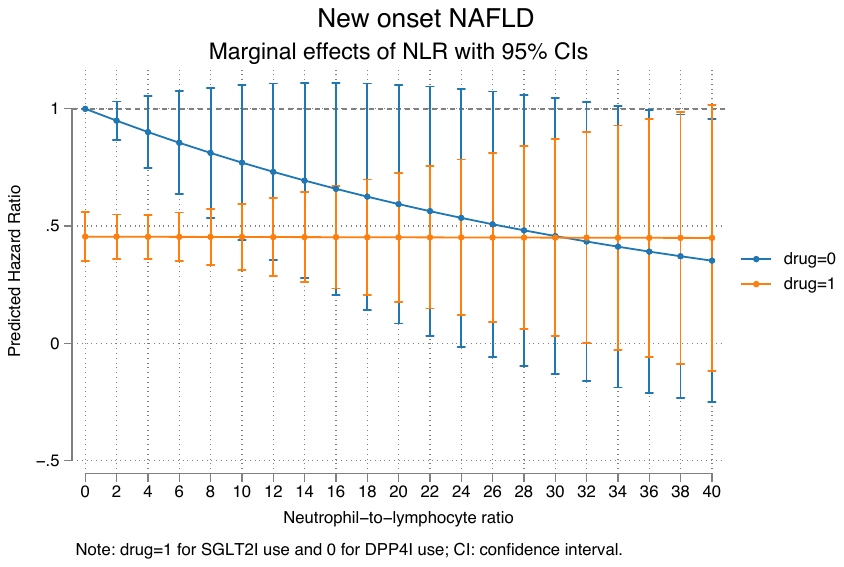

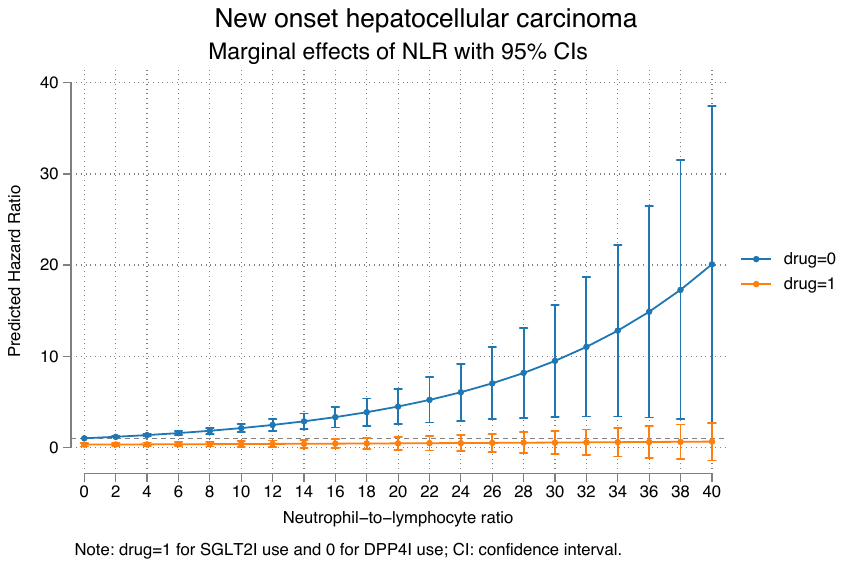

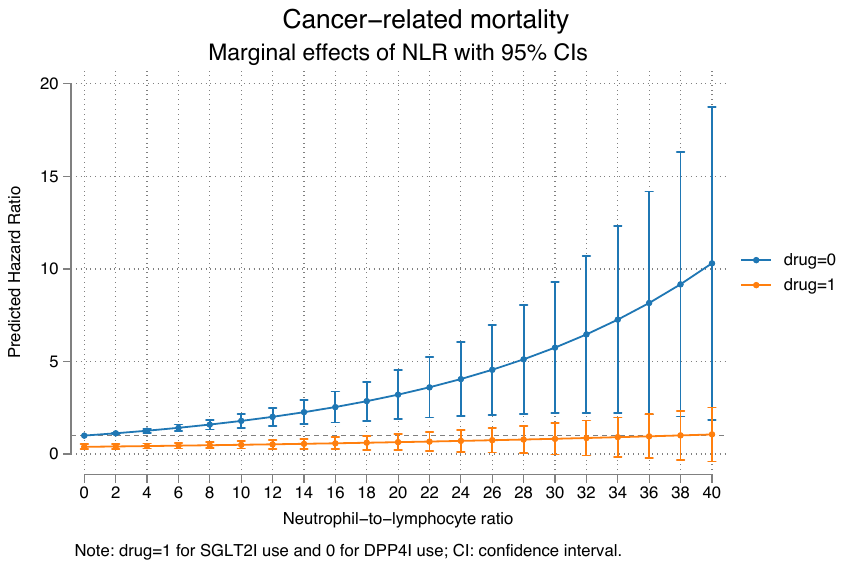

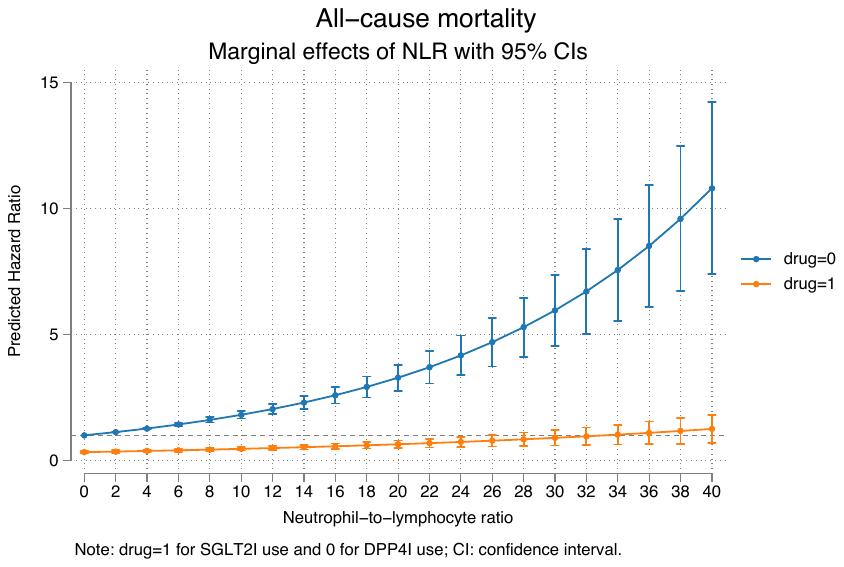

**Supplementary Figure 3B. Interaction effects of NLR and drug exposure of SGLT2I and DPP4I for all-cause mortality, cancer-related mortality, new onset hepatocellular carcinoma, new onset NAFLD in the matched cohort (1:1)**

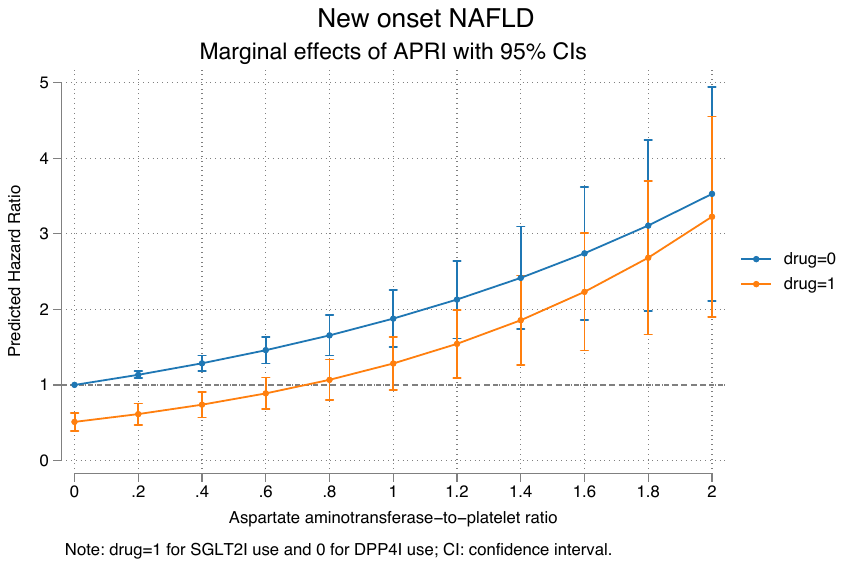

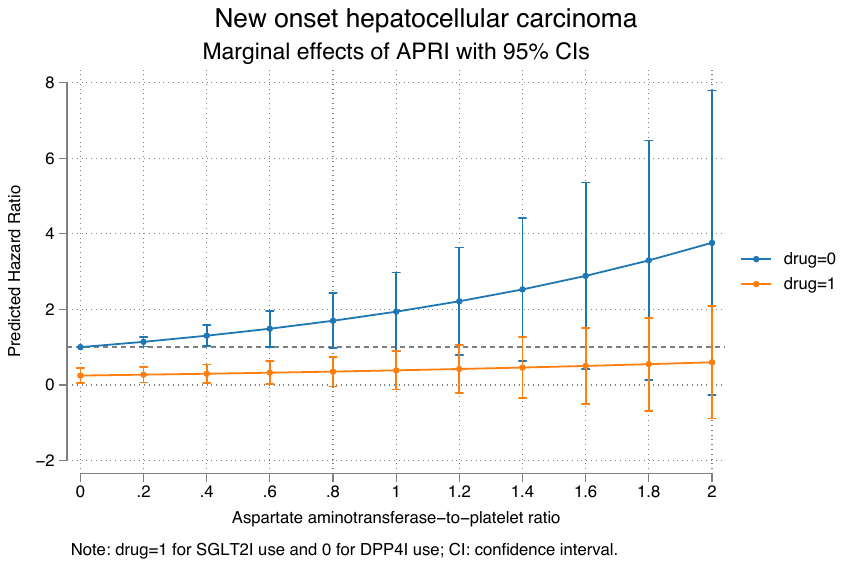

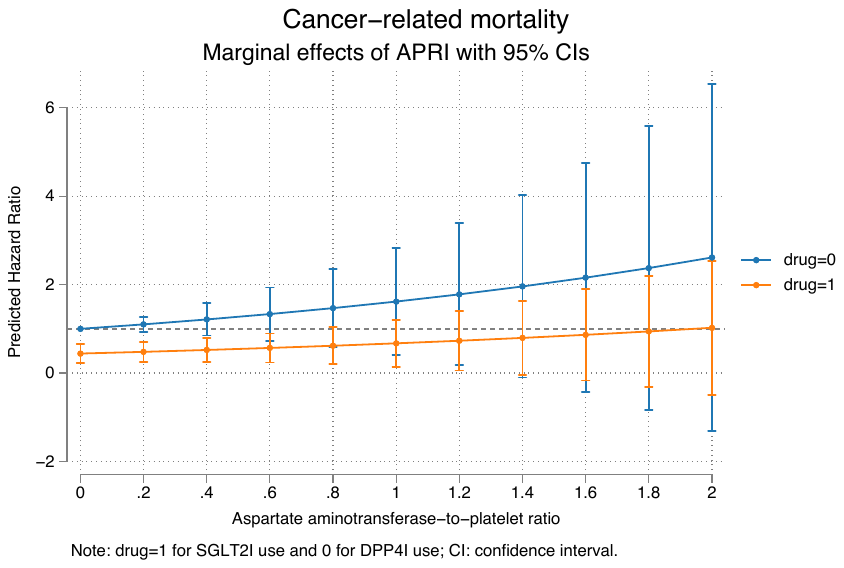

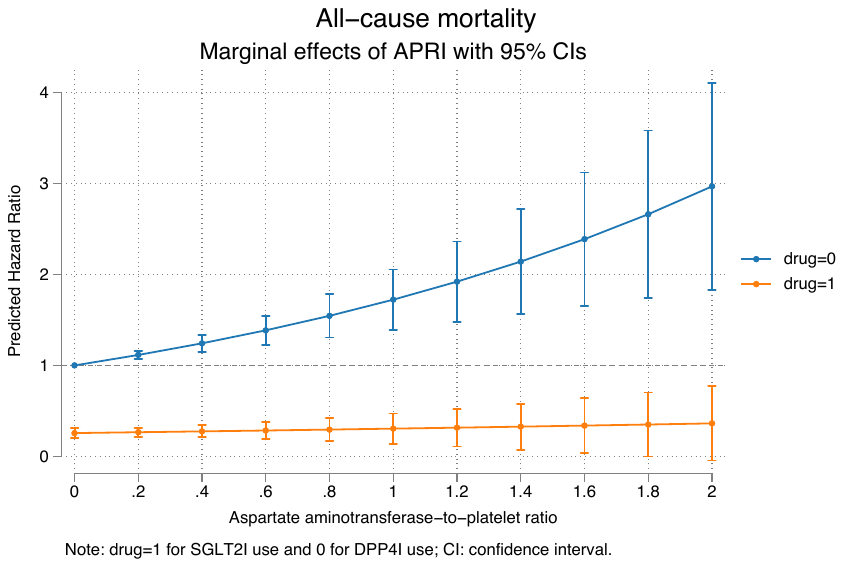

**Supplementary Figure 3C. Interaction effects of APRI and drug exposure of SGLT2I and DPP4I for all-cause mortality, cancer-related mortality, new onset hepatocellular carcinoma, new onset NAFLD in the matched cohort (1:1)**

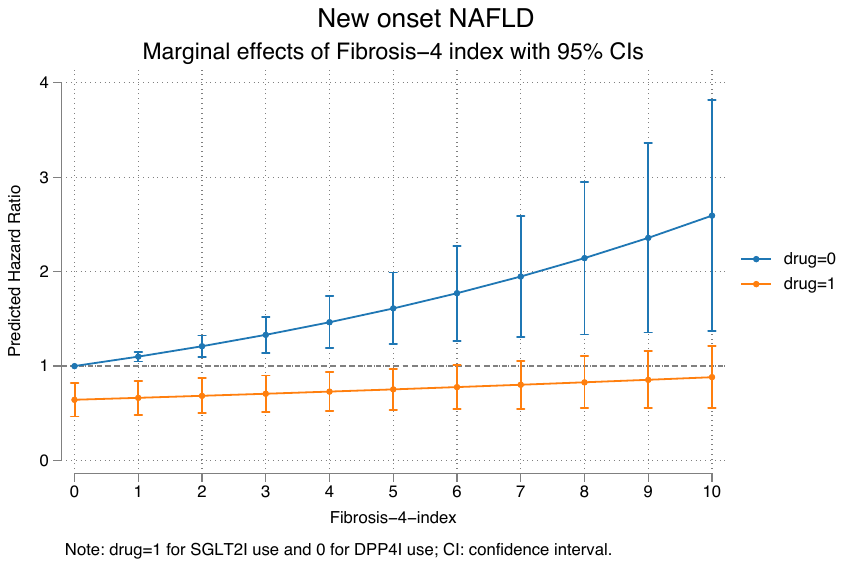

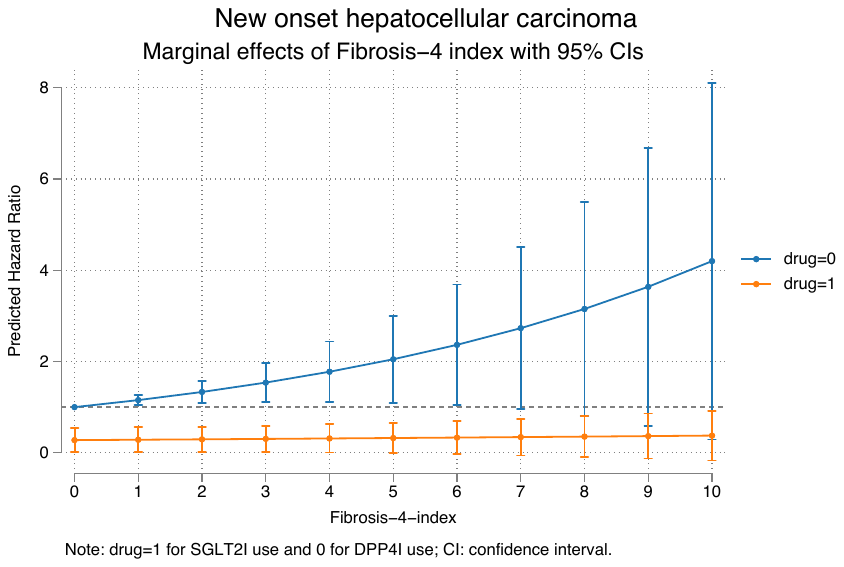

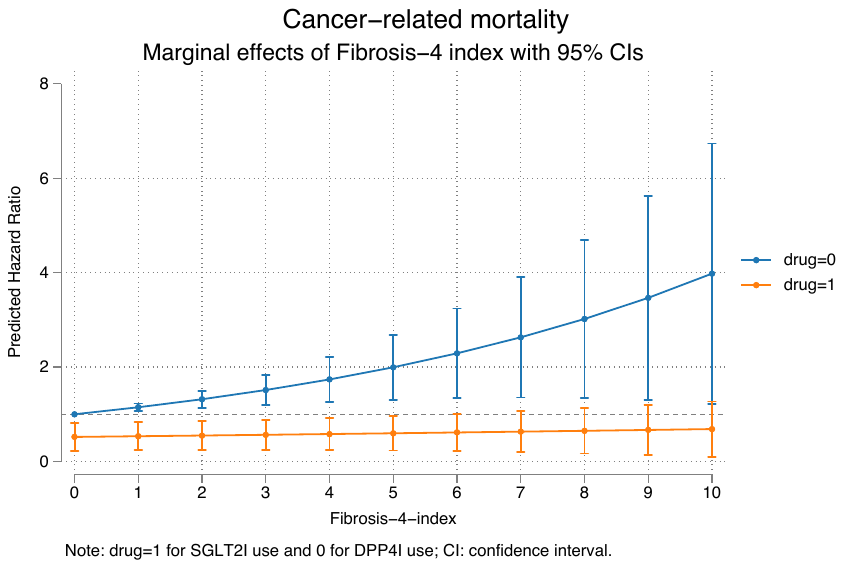

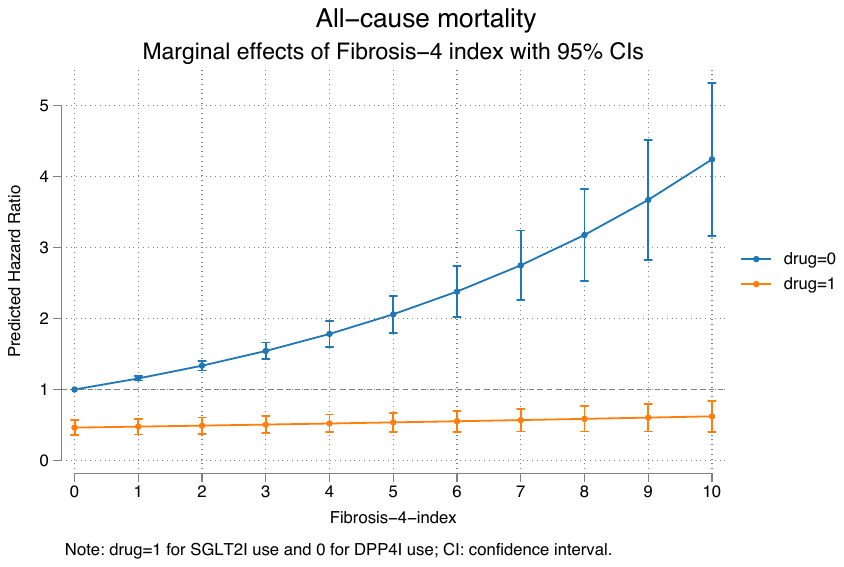

**Supplementary Figure 3D. Interaction effects of Fibrosis-4 index and drug exposure of SGLT2I and DPP4I for all-cause mortality, cancer-related mortality, new onset hepatocellular carcinoma, new onset NAFLD in the matched cohort (1:1)**

**Supplementary Table 1. ICD9 codes for comorbidities and ICD10 codes for outcomes**

Diabetes mellitus 250	250.01	250.02	250.03	250.1	250.11	250.12	250.13	250.2	250.21	250.22	250.23	250.3	250.31	250.32	250.33	250.4	250.41	250.42	250.43	250.5	250.51	250.52	250.53	250.6	250.61	250.62	250.63	250.7	250.71	250.72	250.73	250.8	250.81	250.82	250.83	250.9	250.91	250.92	250.93

Renal diseases	582	582	582.1	582.2	582.4	582.8	582.81	582.89	582.9	583	583	583.1	583.2	583.4	583.6	583.7	585	585.1	585.2	585.3	585.4	585.5	585.6	585.9	586	588	588	588.1	588.8	588.81	588.89	588.9

Acute myocardial infarction 410	410.01	410.02	410.1	410.11	410.12	410.2	410.21	410.22	410.3	410.31	410.32	410.4	410.41	410.42	410.5	410.51	410.52	410.6	410.61	410.62	410.7	410.71	410.72	410.8	410.81	410.82	410.9	410.91	410.92

Heart failure	428	428	428.1	428.2	428.2	428.21	428.22	428.23	428.3	428.3	428.31	428.32	428.33	428.4	428.4	428.41	428.42	428.43	428.9	398.91	402.01	402.11	402.91	404.01	404.03	404.11	404.13	404.91	404.93

Gallstone	574.00	574.01	574.10	574.11	574.20	574.21

Biliary disease	574.30	574.31	574.40	574.41	574.50	574.51	574.60	574.61	574.70	574.71	574.80	574.81	594.90	574.91	575.0	575.10	575.11	575.12	575.2	575.3	575.4	575.5	575.6	575.8	575.9	576.0	576.1	576.2	576.3	576.4	576.5	576.8	576.9

Chronic liver disease and cirrhosis 571.0	571.1	571.2	571.3	571.40	571.41	571.42	471.49	571.5	571.6	571.8,	571.9

Viral hepatitis	070.0	070.1	070.20	070.21	070.22	070.23	070.30	070.31	070.32	070.33	070.41	070.42	070.43	070.44	070.49	070.51	070.52	070.53	070.54	070.59	070.6	070.70	070.71	070.9	573.1	573.2

Disease of pancreas	577.0	577.1	577.2	577.8	577.9

History of acute liver injury 570	572.2

Other Liver disease	275.1	275.0	572.0	572.4	572.1	572.3	572.8	573.0	573.4	573.8	573.9

Peripheral vascular disease	250.7	443.9	443	443.1	443.2	443.21	443.22	443.23	443.24	443.29	443.8	443.81	443.82	443.89	441	443.9	785.4	V43.4

Ischemic heart disease	410.01	410.02	410.1	410.11	410.12	410.2	410.21	410.22	410.3	410.31	410.32	410.4	410.41	410.42	410.5	410.51	410.52	410.6	410.61	410.62	410.7	410.71	410.72	410.8	410.81	410.82	410.9	410.91	410.92	411	411.1	411.8	411.81	411.89	413	413.1	413.9	414	414.01	414.02	414.03	414.04	414.05	414.06	414.07	414.1	414.11	414.12	414.19	414.2	414.3	414.4	414.8	414.9	410	412

Cancer 140	140.1	140.3	140.4	140.5	140.6	140.8	140.9	141	141.1	141.2	141.3	141.4	141.5	141.6	141.8	141.9	142	142.1	142.2	142.8	142.9	143	143.1	143.8	143.9	144	144.1	144.8	144.9	145	145.1	145.2	145.3	145.4	145.5	145.6	145.8	145.9	146	146.1	146.2	146.3	146.4	146.5	146.6	146.7	146.8	146.9	147	147.1	147.2	147.3	147.8	147.9	148	148.1	148.2	148.3	148.8	148.9	149	149.1	149.8	149.9	150	150.1	150.2	150.3	150.4	150.5	150.8	150.9	151	151.1	151.2	151.3	151.4	151.5	151.6	151.8	151.9	152	152.1	152.2	152.3	152.8	152.9	153	153.1	153.2	153.3	153.4	153.5	153.6	153.7	153.8	153.9	154	154.1	154.2	154.3	154.8	155	155.1	155.2	156	156.1	156.2	156.8	156.9	157	157.1	157.2	157.3	157.4	157.8	157.9	158	158.8	158.9	159	159.1	159.8	159.9	160	160.1	160.2	160.3	160.4	160.5	160.8	160.9	161	161.1	161.2	161.3	161.8	161.9	162	162.2	162.3	162.4	162.5	162.8	162.9	163	163.1	163.8	163.9	164	164.1	164.2	164.3	164.8	164.9	165	165.8	165.9	170	170.1	170.2	170.3	170.4	170.5	170.6	170.7	170.8	170.9	171	171.2	171.3	171.4	171.5	171.6	171.7	171.8	171.9	172	172.1	172.2	172.3	172.4	172.5	172.6	172.7	172.8	172.9 173	173.01	173.02	173.09	173.1	173.11	173.12	173.19	173.2	173.21	173.22	173.29	173.3	173.31	173.32	173.39	173.4	173.41	173.42	173.49	173.5	173.51	173.52	173.59	173.6	173.61	173.62	173.69	173.7	173.71	173.72	173.79	173.8	173.81	173.82	173.89	173.9	173.91	173.92	173.99	174	174.1	174.2	174.3	174.4	174.5	174.6	174.8	174.9	175	175.9	176	176.1	176.2	176.3	176.4	176.5	176.8	176.9	179	180	180.1	180.8	180.9	181	182	182.1	182.8	183	183.2	183.3	183.4	183.5	183.8	183.9	184	184.1	184.2	184.3	184.4	184.8	184.9	185	186	186.9	187	187.1	187.2	187.3	187.4	187.5	187.6	187.7	187.8	187.9	188	188.1	188.2	188.3	188.4	188.5	188.6	188.7	188.8	188.9	189	189.1	189.2	189.3	189.4	189.8	189.9	190	190.1	190.2	190.3	190.4	190.5	190.6	190.7	190.8	190.9	191	191.1	191.2	191.3	191.4	191.5	191.6	191.7	191.8	191.9	192	192.1	192.2	192.3	192.8	192.9	193	194	194.1	194.3	194.4	194.5	194.6	194.8	194.9	195	195.1	195.2	195.3	195.4	195.5	195.8	200	200.01	200.02	200.03	200.04	200.05	200.06	200.07	200.08	200.1	200.11	200.12	200.13	200.14	200.15	200.16	200.17	200.18	200.2	200.21	200.22	200.23	200.24	200.25	200.26	200.27	200.28	200.3	200.31	200.32	200.33	200.34	200.35	200.36	200.37	200.38	200.4	200.41	200.42	200.43	200.44	200.45	200.46	200.47	200.48	200.5	200.51	200.52	200.53	200.54	200.55	200.56	200.57	200.58	200.6	200.61	200.62	200.63	200.64	200.65	200.66	200.67	200.68	200.7	200.71	200.72	200.73	200.74	200.75	200.76	200.77	200.78	200.8	200.81	200.82	200.83	200.84	200.85	200.86	200.87	200.88	201	201.01	201.02	201.03	201.04	201.05	201.06	201.07	201.08	201.1	201.11	201.12	201.13	201.14	201.15	201.16	201.17	201.18	201.2	201.21	201.22	201.23	201.24	201.25	201.26	201.27	201.28	201.4	201.41	201.42	201.43	201.44	201.45	201.46	201.47	201.48	201.5	201.51	201.52	201.53	201.54	201.55	201.56	201.57	201.58	201.6	201.61	201.62	201.63	201.64	201.65	201.66	201.67	201.68	201.7	201.71	201.72	201.73	201.74	201.75	201.76	201.77	201.78	201.9	201.91	201.92	201.93	201.94	201.95	201.96	201.97	201.98	202	202.01	202.02	202.03	202.04	202.05	202.06	202.07	202.08	202.1	202.11	202.12	202.13	202.14	202.15	202.16	202.17	202.18	202.2	202.21	202.22	202.23	202.24	202.25	202.26	202.27	202.28	202.3	202.31	202.32	202.33	202.34	202.35	202.36	202.37	202.38	202.4	202.41	202.42	202.43	202.44	202.45	202.46	202.47	202.48	202.5	202.51	202.52	202.53	202.54	202.55	202.56	202.57	202.58	202.6	202.61	202.62	202.63	202.64	202.65	202.66	202.67	202.68	202.7	202.71	202.72	202.73	202.74	202.75	202.76	202.77	202.78	202.8	202.81	202.82	202.83	202.84	202.85	202.86	202.87	202.88	202.9	202.91	202.92	202.93	202.94	202.95	202.96	202.97	202.98	203	203.01	203.02	203.1	203.11	203.12	203.8	203.81	203.82	204	204.01	204.02	204.1	204.11	204.12	204.2	204.21	204.22	204.8	204.81	204.82	204.9	204.91	204.92	205	205.01	205.02	205.1	205.11	205.12	205.2	205.21	205.22	205.3	205.31	205.32	205.8	205.81	205.82	205.9	205.91	205.92	206	206.01	206.02	206.1	206.11	206.12	206.2	206.21	206.22	206.8	206.81	206.82	206.9	206.91	206.92	207	207.01	207.02	207.1	207.11	207.12	207.2	207.21	207.22	207.8	207.81	207.82	208	208.01	208.02	208.1	208.11	208.12	208.2	208.21	208.22	208.8	208.81	208.82	208.9	208.91	208.92 196	196.1	196.2	196.3	196.5	196.6	196.8	196.9	197	197.1	197.2	197.3	197.4	197.5	197.6	197.7	197.8	198	198.1	198.2	198.3	198.4	198.5	198.6	198.7	198.8	198.81	198.82	198.89	199	199.1

Hypertension	401	401.1	401.9	402	402.01	402.1	402.11	402.9	402.91	403	403.01	403.1	403.11	403.9	403.91	404	404.01	404.02	404.03	404.1	404.11	404.12	404.13	404.9	404.91	404.92	404.93	405	405.01	405.09	405.1	405.11	405.19	405.9	405.91	405.99	437.2

Overweight, obesity	278	278	278	278.01	278.02	278.03	278.1	278.2	278.3	278.4	278.8

Alcohol dependence 291	303	535.3	571.0	571.2	571.3	980.0

Diabetic retinopathy 249.5	250.5	362.01	362.02	362.03	362.04	362.05	362.06 362.07 362.1 362.53	362.81	362.82	362.83	362.07	379.23

**Supplementary Table 2. Calculations for SD variability measure**

| **Variability measure** | **Definition** |
| --- | --- |
| Standard deviation | $\sqrt{\frac{1}{Number of measurements}\sum_{i=1}^{Number of measurements} {({test}_{i}-individual mean)}^{2}}$ |

.

**Supplementary Table 3A. Baseline and clinical characteristics of patients with/without all-cause mortality risk before and after propensity score matching (1:1).**

* for SMD≥0.2; SD: standard deviation; SGLT2I: sodium glucose cotransporter-2 inhibitor; DPP4I: dipeptidyl peptidase-4 inhibitor; ACEis: Angiotensin-converting enzyme inhibitors; ARBs: angiotensin receptor blockers; MDRD: modification of diet in renal disease; NAFLD: Non-alcoholic fatty liver disease; # indicated characteristics difference significance.

| **Characteristics** | **Before matching** |  | **SMD^#^** | **After matching** |  | **SMD^#^** |
| --- | --- | --- | --- | --- | --- | --- |
|  | **All-cause mortality (N=7055) Mean(SD);N or Count(%)** | **Alive (N=55644) Mean(SD);N or Count(%)** |  | **All-cause mortality (N=3479) Mean(SD);N or Count(%)** | **Alive(N=40829) Mean(SD);N or Count(%)** |  |
| **Demographics** |  |  |  |  |  |  |
| Male gender | 3800(53.86%) | 30801(55.35%) | 0.03 | 2412(69.33%) | 24664(60.40%) | 0.19 |
| Female gender | 3255(46.13%) | 24843(44.64%) | 0.03 | 1067(30.66%) | 16165(39.59%) | 0.19 |
| Baseline age, years | 75.2(11.5);n=7055 | 61.8(12.2);n=55644 | 1.13* | 66.0(10.9);n=3479 | 57.9(10.8);n=40829 | 0.74* |
| **Past comorbidities** |  |  |  |  |  |  |
| Charlson standard comorbidity index | 3.7(1.7);n=7055 | 1.9(1.4);n=55644 | 1.1* | 2.4(1.4);n=3479 | 1.5(1.2);n=40829 | 0.7* |
| Alcohol dependence | 53(0.75%) | 87(0.15%) | 0.09 | 23(0.66%) | 35(0.08%) | 0.09 |
| Hyperlipidaemia | 189(2.67%) | 1537(2.76%) | 0.01 | 68(1.95%) | 1386(3.39%) | 0.09 |
| Overweight, obesity and hyperalimentation | 23(0.32%) | 421(0.75%) | 0.06 | 25(0.71%) | 657(1.60%) | 0.08 |
| Hypertension | 2788(39.51%) | 12533(22.52%) | 0.37* | 896(25.75%) | 8765(21.46%) | 0.1 |
| Chronic liver disease and cirrhosis | 141(1.99%) | 1168(2.09%) | 0.01 | 41(1.17%) | 1144(2.80%) | 0.12 |
| HBV infection diagnosis | 228(3.23%) | 2215(3.98%) | 0.04 | 252(7.24%) | 1854(4.54%) | 0.11 |
| HCV infection diagnosis | 37(0.52%) | 131(0.23%) | 0.05 | 30(0.86%) | 82(0.20%) | 0.09 |
| History of acute liver injury | 66(0.93%) | 133(0.23%) | 0.09 | 9(0.25%) | 79(0.19%) | 0.01 |
| Other liver disease | 145(2.05%) | 562(1.00%) | 0.09 | 43(1.23%) | 360(0.88%) | 0.03 |
| Gallstone | 13(0.18%) | 60(0.10%) | 0.02 | 6(0.17%) | 50(0.12%) | 0.01 |
| Biliary disease | 193(2.73%) | 711(1.27%) | 0.1 | 45(1.29%) | 428(1.04%) | 0.02 |
| Disease of pancreas | 78(1.10%) | 349(0.62%) | 0.05 | 21(0.60%) | 264(0.64%) | 0.01 |
| Heart failure | 764(10.82%) | 1346(2.41%) | 0.34* | 231(6.63%) | 867(2.12%) | 0.22* |
| Ischemic heart disease | 1117(15.83%) | 5297(9.51%) | 0.19 | 488(14.02%) | 4677(11.45%) | 0.08 |
| Acute myocardial infarction | 395(5.59%) | 1358(2.44%) | 0.16 | 206(5.92%) | 1270(3.11%) | 0.14 |
| Peripheral vascular disease | 178(2.52%) | 321(0.57%) | 0.16 | 61(1.75%) | 188(0.46%) | 0.12 |
| Diabetic retinopathy | 772(10.94%) | 3698(6.64%) | 0.15 | 258(7.41%) | 2588(6.33%) | 0.04 |
| Baseline non-alcoholic fatty liver disease | 125(1.77%) | 1056(1.89%) | 0.01 | 31(0.89%) | 1049(2.56%) | 0.13 |
| Cancer except for hepatocellular carcinoma | 417(5.91%) | 1339(2.40%) | 0.18 | 115(3.30%) | 789(1.93%) | 0.09 |
| Duration from earliest diabetes mellitus diagnosis date to baseline date, day | 624.1(1249.9);n=7055 | 555.6(1280.8);n=55644 | 0.05 | 428.6(1094.2);n=3479 | 544.7(1304.9);n=40829 | 0.1 |
| **Medications** |  |  |  |  |  |  |
| SGLT2I v.s. DPP4I | 613(8.68%) | 21541(38.71%) | 0.75* | 613(17.62%) | 21541(52.75%) | 0.79* |
| SGLT2I frequency | 11.0(15.8);n=613 | 7.1(9.7);n=21541 | 0.29* | 11.0(15.8);n=613 | 7.1(9.7);n=21541 | 0.29* |
| DPP4I frequency | 5.0(12.1);n=6442 | 5.3(6.2);n=34103 | 0.03 | 4.7(8.7);n=2866 | 4.2(6.0);n=19288 | 0.06 |
| SGLT2I duration, days | 449.3(575.9);n=613 | 531.3(674.0);n=21541 | 0.13 | 449.3(575.9);n=613 | 531.3(674.0);n=21541 | 0.13 |
| DPP4I duration, days | 430.8(295.6);n=6442 | 518.9(281.7);n=34103 | 0.31* | 464.5(293.6);n=2866 | 484.4(285.4);n=19288 | 0.07 |
| Metformin | 4988(70.70%) | 50583(90.90%) | 0.53* | 1962(86.12%) | 39324(93.56%) | 0.25* |
| Sulphonylurea | 5441(77.12%) | 42614(76.58%) | 0.01 | 1812(79.54%) | 30894(73.50%) | 0.14 |
| Insulin | 5873(83.24%) | 26023(46.76%) | 0.83* | 1867(81.95%) | 20832(49.56%) | 0.73* |
| Acarbose | 1536(21.77%) | 6416(11.53%) | 0.28* | 117(5.13%) | 1404(3.34%) | 0.09 |
| Glucagon-like peptide-1 receptor agonists | 412(5.83%) | 3750(6.73%) | 0.04 | 33(1.44%) | 2476(5.89%) | 0.24* |
| Thiozolidinedone | 2131(30.20%) | 17045(30.63%) | 0.01 | 450(19.75%) | 11195(26.63%) | 0.16 |
| ACEI | 1032(14.62%) | 3686(6.62%) | 0.26* | 239(10.49%) | 2609(6.20%) | 0.16 |
| ARB | 397(5.62%) | 1724(3.09%) | 0.12 | 67(2.94%) | 1170(2.78%) | 0.01 |
| Antihypertensive drugs | 851(12.06%) | 2683(4.82%) | 0.26* | 128(5.61%) | 1113(2.64%) | 0.15 |
| HCV treatment drugs | 68(0.96%) | 579(1.04%) | 0.01 | 39(1.71%) | 606(1.44%) | 0.02 |
| HBV treatment drugs | 72(1.02%) | 715(1.28%) | 0.02 | 40(1.75%) | 741(1.76%) | <0.01 |
| Aspirin | 838(11.87%) | 2420(4.34%) | 0.28* | 200(8.77%) | 1791(4.26%) | 0.18 |
| Anticoagulants | 2014(28.54%) | 8472(15.22%) | 0.33* | 693(30.42%) | 7467(17.76%) | 0.30* |
| Antiplatelets | 1093(15.49%) | 3433(6.16%) | 0.30* | 298(13.08%) | 2605(6.19%) | 0.23* |
| Lipid-lowering drugs | 1341(19.00%) | 6964(12.51%) | 0.18 | 467(20.50%) | 6371(15.15%) | 0.14 |
| Statins and fibrates | 2712(38.44%) | 31160(55.99%) | 0.36* | 1416(62.15%) | 29691(70.64%) | 0.18 |
| Nitrates | 590(8.36%) | 1723(3.09%) | 0.23* | 194(8.51%) | 1409(3.35%) | 0.22* |
| Diuretics | 1446(20.49%) | 3906(7.01%) | 0.40* | 401(17.60%) | 2836(6.74%) | 0.34* |
| Beta-blockers | 887(12.57%) | 2731(4.90%) | 0.27* | 277(12.15%) | 2119(5.04%) | 0.26* |
| Calcium channel blockers | 1753(24.84%) | 11209(20.14%) | 0.11 | 571(25.06%) | 9197(21.88%) | 0.08 |
| Steroids | 17(0.24%) | 79(0.14%) | 0.02 | 0(0.00%) | 68(0.16%) | 0.06 |
| **Calculated biomarkers** |  |  |  |  |  |  |
| Abbreviated MDRD, mL/min/1.73m^2 | 61.5(38.9);n=6101 | 89.0(39.5);n=46235 | 0.7* | 74.9(33.0);n=2084 | 86.5(28.1);n=31146 | 0.38* |
| Neutrophil-to-lymphocyte ratio | 5.1(6.8);n=4193 | 3.3(4.3);n=21961 | 0.31* | 4.5(6.3);n=1217 | 2.9(3.4);n=16053 | 0.31* |
| Aspartate aminotransferase-to-platelet ratio | 0.2(0.5);n=1990 | 0.1(0.4);n=10948 | 0.08 | 0.2(0.4);n=613 | 0.1(0.2);n=8125 | 0.1 |
| Fibrosis-4-index | 2.7(7.7);n=1223 | 1.6(4.4);n=6737 | 0.18 | 2.3(3.5);n=350 | 1.3(1.7);n=5434 | 0.34* |
| **Liver function tests** |  |  |  |  |  |  |
| Alkaline phosphatase, U/L | 88.9(48.7);n=5136 | 75.5(29.1);n=35226 | 0.33* | 83.5(40.8);n=1611 | 75.0(26.1);n=24771 | 0.25* |
| Aspartate transaminase, U/L | 28.1(49.0);n=2158 | 28.0(53.7);n=13889 | <0.01 | 29.2(58.9);n=720 | 27.2(23.6);n=10457 | 0.05 |
| Alanine transaminase, U/L | 23.5(36.4);n=4270 | 29.4(33.9);n=30042 | 0.17 | 26.4(44.0);n=1295 | 30.9(25.8);n=20748 | 0.13 |
| Bilirubin, umol/L | 10.8(8.6);n=5120 | 11.2(6.7);n=35044 | 0.06 | 11.18(7.27);n=1611 | 11.15(5.78);n=24717 | <0.01 |
| **Lipid and glucose profiles** |  |  |  |  |  |  |
| Triglyceride, mmol/L | 1.6(1.2);n=5154 | 1.7(1.5);n=43891 | 0.08 | 1.6(1.4);n=1895 | 1.8(1.6);n=30012 | 0.13 |
| SD of triglyceride | 0.4(0.6);n=2556 | 0.5(1.0);n=22844 | 0.07 | 0.4(0.5);n=1003 | 0.5(1.0);n=17324 | 0.16 |
| Low-density lipoprotein, mmol/L | 2.3(0.9);n=5080 | 2.4(0.8);n=43156 | 0.06 | 2.3(0.8);n=1884 | 2.4(0.8);n=29568 | 0.03 |
| SD of low-density lipoprotein | 0.38(0.38);n=2492 | 0.36(0.34);n=22216 | 0.07 | 0.38(0.35);n=963 | 0.36(0.34);n=16893 | 0.05 |
| High-density lipoprotein, mmol/L | 1.21(0.37);n=5147 | 1.2(0.33);n=43831 | 0.03 | 1.19(0.38);n=1895 | 1.19(0.33);n=29965 | 0.02 |
| SD of high-density lipoprotein | 0.13(0.11);n=2482 | 0.1(0.08);n=21963 | 0.31* | 0.12(0.1);n=942 | 0.1(0.08);n=16693 | 0.22* |
| Total cholesterol, mmol/L | 4.26(1.09);n=5164 | 4.33(0.98);n=43931 | 0.08 | 4.2(1.0);n=1895 | 4.3(1.0);n=30035 | 0.1 |
| SD of total cholesterol | 0.5(0.4);n=2548 | 0.4(0.4);n=22873 | 0.1 | 0.44(0.39);n=981 | 0.44(0.42);n=17280 | 0.02 |
| Hemoglobin A1C, % | 7.9(1.8);n=5694 | 8.0(1.5);n=45548 | 0.06 | 8.1(1.7);n=1993 | 8.2(1.5);n=30912 | 0.02 |
| SD of hemoglobin A1C | 0.7(0.9);n=3758 | 0.5(0.7);n=32375 | 0.18 | 0.7(0.6);n=1384 | 0.6(0.8);n=23350 | 0.12 |
| Fasting glucose, mmol/L | 9.2(5.2);n=5307 | 8.9(3.7);n=41126 | 0.07 | 9.3(4.8);n=1748 | 9.0(3.5);n=27329 | 0.07 |
| SD of fasting glucose | 2.8(2.8);n=3663 | 1.9(2.1);n=25509 | 0.38* | 2.6(2.5);n=1206 | 1.8(1.8);n=18963 | 0.36* |

**Supplementary Table 3B. Baseline and clinical characteristics of patients with/without new onset hepatocellular carcinoma before and after propensity score matching (1:1).**

* for SMD≥0.2; SD: standard deviation; SGLT2I: sodium glucose cotransporter-2 inhibitor; DPP4I: dipeptidyl peptidase-4 inhibitor; ACEis: Angiotensin-converting enzyme inhibitors; ARBs: angiotensin receptor blockers; MDRD: modification of diet in renal disease; NAFLD: Non-alcoholic fatty liver disease; # indicated the characteristics difference significance.

| **Characteristics** | **Before matching** |  | **SMD^#^** | **After matching** |  | **SMD^#^** |
| --- | --- | --- | --- | --- | --- | --- |
|  | **New onset hepatocellular carcinoma (N=256) Mean(SD);N or Count(%)** | **No new onset hepatocellular carcinoma (N=62443) Mean(SD);N or Count(%)** |  | **New onset hepatocellular carcinoma (N=187) Mean(SD);N or Count(%)** | **No new onset hepatocellular carcinoma (N=44121) Mean(SD);N or Count(%)** |  |
| **Demographics** |  |  |  |  |  |  |
| Male gender | 193(75.39%) | 34408(55.10%) | 0.44* | 155(82.88%) | 26921(61.01%) | 0.50* |
| Female gender | 63(24.60%) | 28035(44.89%) | 0.44* | 32(17.11%) | 17200(38.98%) | 0.50* |
| Baseline age, years | 67.0(10.6);n=256 | 63.3(12.9);n=62443 | 0.32* | 65.0(9.4);n=187 | 58.5(11.0);n=44121 | 0.64* |
| **Past comorbidities** |  |  |  |  |  |  |
| Charlson standard comorbidity index | 2.7(1.6);n=256 | 2.1(1.5);n=62443 | 0.37* | 2.3(1.3);n=187 | 1.6(1.2);n=44121 | 0.57* |
| Alcohol dependence | 12(4.68%) | 128(0.20%) | 0.29* | 4(2.13%) | 54(0.12%) | 0.19 |
| Hyperlipidaemia | 6(2.34%) | 1720(2.75%) | 0.03 | 1(0.53%) | 1453(3.29%) | 0.20* |
| Overweight, obesity and hyperalimentation | 3(1.17%) | 441(0.70%) | 0.05 | 36(19.25%) | 646(1.46%) | 0.61* |
| Hypertension | 82(32.03%) | 15239(24.40%) | 0.17 | 88(47.05%) | 9573(21.69%) | 0.55* |
| Chronic liver disease and cirrhosis | 23(8.98%) | 1286(2.05%) | 0.31* | 10(5.34%) | 1175(2.66%) | 0.14 |
| HBV infection | 73(28.51%) | 2370(3.79%) | 0.71* | 49(26.20%) | 2057(4.66%) | 0.62* |
| HCV infection | 10(3.90%) | 158(0.25%) | 0.26* | 0(0.00%) | 112(0.25%) | 0.07 |
| History of acute liver injury | 22(8.59%) | 177(0.28%) | 0.41* | 9(4.81%) | 79(0.17%) | 0.30* |
| Other liver disease | 19(7.42%) | 688(1.10%) | 0.32* | 7(3.74%) | 396(0.89%) | 0.19 |
| Gallstone | 12(4.68%) | 61(0.09%) | 0.30* | 7(3.74%) | 49(0.11%) | 0.27* |
| Bilirary disease | 7(2.73%) | 897(1.43%) | 0.09 | 2(1.06%) | 471(1.06%) | <0.01 |
| Disease of pancreas | 3(1.17%) | 424(0.67%) | 0.05 | 0(0.00%) | 285(0.64%) | 0.11 |
| Heart failure | 8(3.12%) | 2102(3.36%) | 0.01 | 1(0.53%) | 1097(2.48%) | 0.16 |
| Ischemic heart disease | 30(11.71%) | 6384(10.22%) | 0.05 | 14(7.48%) | 5151(11.67%) | 0.14 |
| Acute myocardial infarction | 12(4.68%) | 1741(2.78%) | 0.1 | 4(2.13%) | 1472(3.33%) | 0.07 |
| Peripheral vascular disease | 1(0.39%) | 498(0.79%) | 0.05 | 0(0.00%) | 249(0.56%) | 0.11 |
| Diabetic retinopathy | 18(7.03%) | 4452(7.12%) | <0.01 | 5(2.67%) | 2841(6.43%) | 0.18 |
| Baseline non-alcoholic fatty liver disease | 21(8.20%) | 1160(1.85%) | 0.29* | 9(4.81%) | 1071(2.42%) | 0.13 |
| Cancer except hepatocellular carcinoma | 8(3.12%) | 1748(2.79%) | 0.02 | 17(9.09%) | 887(2.01%) | 0.31* |
| Duration from earliest diabetes mellitus diagnosis date to baseline date, day | 419.7(963.7);n=256 | 563.9(1278.7);n=62443 | 0.13 | 201.4(527.5);n=187 | 537.0(1292.1);n=44121 | 0.34* |
| **Medications** |  |  |  |  |  |  |
| SGLT2I v.s. DPP4I | 36(14.06%) | 22118(35.42%) | 0.51* | 36(19.25%) | 22118(50.13%) | 0.69* |
| SGLT2I frequency | 13.5(17.1);n=36 | 7.2(9.9);n=22118 | 0.45* | 13.5(17.1);n=36 | 7.2(9.9);n=22118 | 0.45* |
| DPP4I frequency | 7.6(14.6);n=220 | 5.2(7.4);n=40325 | 0.21* | 3.8(4.7);n=151 | 4.2(6.4);n=22003 | 0.08 |
| SGLT2I duration, days | 750.2(708.9);n=36 | 528.6(671.5);n=22118 | 0.32* | 750.2(708.9);n=36 | 528.6(671.5);n=22118 | 0.32* |
| DPP4I duration, days | 476.8(294.8);n=220 | 505.1(285.7);n=40325 | 0.1 | 548.6(294.6);n=151 | 481.4(286.4);n=22003 | 0.23* |
| Metformin | 215(83.98%) | 55356(88.65%) | 0.14 | 143(95.97%) | 41143(93.17%) | 0.12 |
| Sulphonylurea | 192(74.99%) | 47863(76.65%) | 0.04 | 127(85.23%) | 32579(73.77%) | 0.29* |
| Insulin | 209(81.64%) | 31687(50.74%) | 0.69* | 123(82.55%) | 22576(51.12%) | 0.71* |
| Acarbose | 40(15.62%) | 7912(12.67%) | 0.08 | 6(4.02%) | 1515(3.43%) | 0.03 |
| Glucagon-like peptide-1 receptor agonists | 8(3.12%) | 4154(6.65%) | 0.16 | 2(1.34%) | 2507(5.67%) | 0.24* |
| Thiozolidinedone | 61(23.82%) | 19115(30.61%) | 0.15 | 17(11.40%) | 11628(26.33%) | 0.39* |
| ACEI | 33(12.89%) | 4685(7.50%) | 0.18 | 14(9.39%) | 2834(6.41%) | 0.11 |
| ARB | 13(5.07%) | 2108(3.37%) | 0.08 | 3(2.01%) | 1234(2.79%) | 0.05 |
| Antihypertensive drugs | 24(9.37%) | 3510(5.62%) | 0.14 | 9(6.04%) | 1232(2.78%) | 0.16 |
| HCV treatment drugs | 47(18.35%) | 600(0.96%) | 0.62* | 23(15.43%) | 622(1.40%) | 0.52* |
| HBV treatment drugs | 55(21.48%) | 732(1.17%) | 0.68* | 24(16.10%) | 757(1.71%) | 0.52* |
| Aspirin | 15(5.85%) | 3243(5.19%) | 0.03 | 6(4.02%) | 1985(4.49%) | 0.02 |
| Anticoagulants | 69(26.95%) | 10417(16.68%) | 0.25* | 33(22.14%) | 8127(18.40%) | 0.09 |
| Antiplatelets | 31(12.10%) | 4495(7.19%) | 0.17 | 14(9.39%) | 2889(6.54%) | 0.11 |
| Lipid-lowering drugs | 40(15.62%) | 8265(13.23%) | 0.07 | 16(10.73%) | 6822(15.44%) | 0.14 |
| Statins and fibrates | 122(47.65%) | 33750(54.04%) | 0.13 | 98(65.77%) | 31009(70.22%) | 0.1 |
| Nitrates | 13(5.07%) | 2300(3.68%) | 0.07 | 3(2.01%) | 1600(3.62%) | 0.1 |
| Diuretics | 49(19.14%) | 5303(8.49%) | 0.31* | 17(11.40%) | 3220(7.29%) | 0.14 |
| Beta-blockers | 20(7.81%) | 3598(5.76%) | 0.08 | 3(2.01%) | 2393(5.41%) | 0.18 |
| Calcium channel blockers | 74(28.90%) | 12888(20.63%) | 0.19 | 38(25.50%) | 9730(22.03%) | 0.08 |
| Steroids | 0(0.00%) | 96(0.15%) | 0.06 | 0(0.00%) | 68(0.15%) | 0.06 |
| **Calculated biomarkers** |  |  |  |  |  |  |
| Abbreviated MDRD, mL/min/1.73m^2 | 87.9(45.7);n=223 | 85.8(40.4);n=52113 | 0.05 | 76.7(24.3);n=143 | 85.8(28.6);n=33087 | 0.34* |
| Neutrophil-to-lymphocyte ratio | 4.4(7.2);n=132 | 3.6(4.8);n=26022 | 0.13 | 5.9(13.3);n=62 | 3.0(3.7);n=17208 | 0.3* |
| Aspartate aminotransferase-to-platelet ratio | 0.5(1.1);n=85 | 0.1(0.4);n=12853 | 0.47* | 0.2(0.1);n=35 | 0.1(0.2);n=8703 | 0.47* |
| Fibrosis-4-index | 4.7(5.9);n=55 | 1.8(5.1);n=7905 | 0.54* | 2.3(1.3);n=25 | 1.4(1.9);n=5759 | 0.54* |
| **Liver function tests** |  |  |  |  |  |  |
| Aspartate transaminase, U/L | 48.4(86.1);n=104 | 27.9(52.8);n=15943 | 0.29* | 33.7(16.6);n=42 | 27.3(27.3);n=11135 | 0.28* |
| Alanine transaminase, U/L | 46.4(53.4);n=146 | 28.6(34.1);n=34166 | 0.4* | 50.0(62.6);n=71 | 30.6(27.0);n=21972 | 0.4* |
| Bilirubin, umol/L | 15.3(10.7);n=188 | 11.1(7.0);n=39976 | 0.46* | 13.6(6.6);n=83 | 11.1(5.9);n=26245 | 0.39* |
| **Lipid and glucose profiles** |  |  |  |  |  |  |
| Triglyceride, mmol/L | 1.3(0.7);n=192 | 1.7(1.5);n=48853 | 0.31* | 1.3(0.7);n=131 | 1.8(1.6);n=31776 | 0.34* |
| SD of triglyceride | 0.3(0.5);n=84 | 0.5(1.0);n=25316 | 0.2* | 0.4(0.3);n=52 | 0.5(1.0);n=18275 | 0.1 |
| Low-density lipoprotein, mmol/L | 2.2(0.7);n=192 | 2.4(0.8);n=48044 | 0.18 | 2.3(0.7);n=131 | 2.4(0.8);n=31321 | 0.02 |
| SD of low-density lipoprotein | 0.3(0.2);n=85 | 0.4(0.3);n=24623 | 0.29* | 0.2(0.2);n=54 | 0.4(0.3);n=17802 | 0.55* |
| High-density lipoprotein, mmol/L | 1.23(0.42);n=192 | 1.2(0.33);n=48786 | 0.08 | 1.19(0.26);n=131 | 1.19(0.33);n=31729 | 0.02 |
| SD of high-density lipoprotein | 0.13(0.13);n=79 | 0.1(0.08);n=24366 | 0.26* | 0.11(0.08);n=49 | 0.1(0.08);n=17586 | 0.17 |
| Total cholesterol, mmol/L | 4.1(0.9);n=192 | 4.3(1.0);n=48903 | 0.23* | 4.1(0.6);n=131 | 4.3(1.0);n=31799 | 0.21* |
| SD of total cholesterol | 0.3(0.3);n=84 | 0.4(0.4);n=25337 | 0.33* | 0.3(0.1);n=53 | 0.4(0.4);n=18208 | 0.49* |
| Hemoglobin A1C, % | 7.97(1.57);n=204 | 8.03(1.54);n=51038 | 0.04 | 7.7(1.4);n=138 | 8.2(1.5);n=32767 | 0.3* |
| SD of hemoglobin A1C | 0.58(0.59);n=133 | 0.57(0.72);n=36000 | 0.02 | 0.4(0.3);n=108 | 0.6(0.8);n=24626 | 0.23* |
| Fasting glucose, mmol/L | 8.6(3.4);n=189 | 8.9(3.9);n=46244 | 0.07 | 7.9(2.4);n=118 | 9.1(3.6);n=28959 | 0.37* |
| SD of fasting glucose | 2.1(2.1);n=116 | 2.0(2.2);n=29056 | 0.06 | 2.1(1.3);n=57 | 1.9(1.9);n=20112 | 0.15 |

**Supplementary Table 3C. Baseline and clinical characteristics of patients with/without new onset NAFLD before and after propensity score matching (1:1).**

* for SMD≥0.2; SD: standard deviation; SGLT2I: sodium glucose cotransporter-2 inhibitor; DPP4I: dipeptidyl peptidase-4 inhibitor; ACEis: Angiotensin-converting enzyme inhibitors; ARBs: angiotensin receptor blockers; MDRD: modification of diet in renal disease; NAFLD: Non-alcoholic fatty liver disease; # indicated the characteristics difference significance.

| **Characteristics** | **Before matching** |  | **SMD^#^** | **After matching** |  | **SMD^#^** |
| --- | --- | --- | --- | --- | --- | --- |
|  | **New onset NAFLD (N=1359) Mean(SD);N or Count(%)** | **No new onset NAFLD (N=61340) Mean(SD);N or Count(%)** |  | **New onset NAFLD (N=1090) Mean(SD);N or Count(%)** | **No new onset NAFLD (N=43218) Mean(SD);N or Count(%)** |  |
| **Demographics** |  |  |  |  |  |  |
| Male gender | 678(49.88%) | 33923(55.30%) | 0.11 | 531(48.71%) | 26545(61.42%) | 0.26* |
| Female gender | 681(50.11%) | 27417(44.69%) | 0.11 | 559(51.28%) | 16673(38.57%) | 0.26* |
| Baseline age, years | 60.4(12.6);n=1359 | 63.4(12.9);n=61340 | 0.23* | 56.0(12.1);n=1090 | 58.6(11.0);n=43218 | 0.23* |
| **Past comorbidities** |  |  |  |  |  |  |
| Charlson standard comorbidity index | 2.0(1.8);n=1359 | 2.1(1.5);n=61340 | 0.06 | 1.4(1.4);n=1090 | 1.6(1.2);n=43218 | 0.11 |
| Alcohol dependence | 24(1.76%) | 116(0.18%) | 0.16 | 5(0.45%) | 53(0.12%) | 0.06 |
| Hyperlipidaemia | 44(3.23%) | 1682(2.74%) | 0.03 | 35(3.21%) | 1419(3.28%) | <0.01 |
| Overweight, obesity and hyperalimentation | 27(1.98%) | 417(0.67%) | 0.11 | 80(7.33%) | 602(1.39%) | 0.29* |
| Hypertension | 375(27.59%) | 14946(24.36%) | 0.07 | 360(33.02%) | 9301(21.52%) | 0.26* |
| Chronic liver disease and cirrhosis | 265(19.49%) | 1044(1.70%) | 0.60* | 189(17.33%) | 996(2.30%) | 0.52* |
| HBV infection | 166(12.21%) | 2277(3.71%) | 0.32* | 114(10.45%) | 1992(4.60%) | 0.22* |
| HCV infection | 24(1.76%) | 144(0.23%) | 0.15 | 5(0.45%) | 107(0.24%) | 0.04 |
| History of acute liver injury | 78(5.73%) | 121(0.19%) | 0.33* | 29(2.66%) | 59(0.13%) | 0.22* |
| Other liver disease | 71(5.22%) | 636(1.03%) | 0.24* | 43(3.94%) | 360(0.83%) | 0.20* |
| Gallstone | 17(1.25%) | 56(0.09%) | 0.14 | 11(1.00%) | 45(0.10%) | 0.12 |
| Bilirary disease | 35(2.57%) | 869(1.41%) | 0.08 | 34(3.11%) | 439(1.01%) | 0.15 |
| Disease of pancreas | 18(1.32%) | 409(0.66%) | 0.07 | 13(1.19%) | 272(0.62%) | 0.06 |
| Heart failure | 55(4.04%) | 2055(3.35%) | 0.04 | 44(4.03%) | 1054(2.43%) | 0.09 |
| Ischemic heart disease | 116(8.53%) | 6298(10.26%) | 0.06 | 123(11.28%) | 5042(11.66%) | 0.01 |
| Acute myocardial infarction | 25(1.83%) | 1728(2.81%) | 0.06 | 14(1.28%) | 1462(3.38%) | 0.14 |
| Peripheral vascular disease | 10(0.73%) | 489(0.79%) | 0.01 | 4(0.36%) | 245(0.56%) | 0.03 |
| Diabetic retinopathy | 77(5.66%) | 4393(7.16%) | 0.06 | 45(4.12%) | 2801(6.48%) | 0.11 |
| Baseline non-alcoholic fatty liver disease | 266(19.57%) | 915(1.49%) | 0.62* | 193(17.70%) | 887(2.05%) | 0.54* |
| Cancer except hepatocellular carcinoma | 34(2.50%) | 1722(2.80%) | 0.02 | 18(1.65%) | 886(2.05%) | 0.03 |
| Duration from earliest diabetes mellitus diagnosis date to baseline date, day | 574.6(1237.1);n=1359 | 563.1(1278.4);n=61340 | 0.01 | 556.6(1271.8);n=1090 | 535.0(1290.4);n=43218 | 0.02 |
| **Medications** |  |  |  |  |  |  |
| SGLT2I v.s. DPP4I | 410(30.16%) | 21744(35.44%) | 0.11 | 410(37.61%) | 21744(50.31%) | 0.26* |
| SGLT2I frequency | 9.7(14.2);n=410 | 7.2(9.8);n=21744 | 0.21* | 9.7(14.2);n=410 | 7.2(9.8);n=21744 | 0.21* |
| DPP4I frequency | 5.9(9.7);n=949 | 5.2(7.4);n=39596 | 0.09 | 10.1(16.7);n=680 | 4.1(5.7);n=21474 | 0.48* |
| SGLT2I duration, days | 602.7(676.7);n=410 | 527.6(671.4);n=21744 | 0.11 | 602.7(676.7);n=410 | 527.6(671.4);n=21744 | 0.11 |
| DPP4I duration, days | 501.5(287.0);n=949 | 505.0(285.7);n=39596 | 0.01 | 456.8(269.2);n=680 | 482.6(287.0);n=21474 | 0.09 |
| Metformin | 1210(89.03%) | 54361(88.62%) | 0.01 | 910(92.47%) | 40376(93.19%) | 0.03 |
| Sulphonylurea | 1066(78.44%) | 46989(76.60%) | 0.04 | 789(80.18%) | 31917(73.67%) | 0.16 |
| Insulin | 792(58.27%) | 31104(50.70%) | 0.15 | 551(55.99%) | 22148(51.12%) | 0.1 |
| Acarbose | 198(14.56%) | 7754(12.64%) | 0.06 | 31(3.15%) | 1490(3.43%) | 0.02 |
| Glucagon-like peptide-1 receptor agonists | 122(8.97%) | 4040(6.58%) | 0.09 | 72(7.31%) | 2437(5.62%) | 0.07 |
| Thiozolidinedone | 477(35.09%) | 18699(30.48%) | 0.1 | 265(26.93%) | 11380(26.26%) | 0.02 |
| ACEI | 138(10.15%) | 4580(7.46%) | 0.09 | 67(6.80%) | 2781(6.41%) | 0.02 |
| ARB | 58(4.26%) | 2063(3.36%) | 0.05 | 36(3.65%) | 1201(2.77%) | 0.05 |
| Antihypertensive drugs | 88(6.47%) | 3446(5.61%) | 0.04 | 18(1.82%) | 1223(2.82%) | 0.07 |
| HCV treatment drugs | 74(5.44%) | 573(0.93%) | 0.26* | 53(5.38%) | 592(1.36%) | 0.22* |
| HBV treatment drugs | 80(5.88%) | 707(1.15%) | 0.26* | 54(5.48%) | 727(1.67%) | 0.21* |
| Aspirin | 95(6.99%) | 3163(5.15%) | 0.08 | 48(4.87%) | 1943(4.48%) | 0.02 |
| Anticoagulants | 284(20.89%) | 10202(16.63%) | 0.11 | 157(15.95%) | 8003(18.47%) | 0.07 |
| Antiplatelets | 121(8.90%) | 4405(7.18%) | 0.06 | 62(6.30%) | 2841(6.55%) | 0.01 |
| Lipid-lowering drugs | 200(14.71%) | 8105(13.21%) | 0.04 | 131(13.31%) | 6707(15.48%) | 0.06 |
| Statins and fibrates | 799(58.79%) | 33073(53.91%) | 0.1 | 732(74.39%) | 30375(70.11%) | 0.1 |
| Nitrates | 64(4.70%) | 2249(3.66%) | 0.05 | 20(2.03%) | 1583(3.65%) | 0.1 |
| Diuretics | 192(14.12%) | 5160(8.41%) | 0.18 | 79(8.02%) | 3158(7.28%) | 0.03 |
| Beta-blockers | 94(6.91%) | 3524(5.74%) | 0.05 | 50(5.08%) | 2346(5.41%) | 0.01 |
| Calcium channel blockers | 308(22.66%) | 12654(20.62%) | 0.05 | 208(21.13%) | 9560(22.06%) | 0.02 |
| Steroids | 3(0.22%) | 93(0.15%) | 0.02 | 2(0.20%) | 66(0.15%) | 0.01 |
| **Calculated biomarkers** |  |  |  |  |  |  |
| Abbreviated MDRD, mL/min/1.73m^2 | 91.7(42.9);n=1235 | 85.7(40.4);n=51101 | 0.14 | 95.1(39.9);n=933 | 85.5(28.2);n=32297 | 0.28* |
| Neutrophil-to-lymphocyte ratio | 3.7(5.5);n=754 | 3.6(4.8);n=25400 | 0.01 | 2.9(2.9);n=581 | 3.0(3.8);n=16689 | 0.04 |
| Aspartate aminotransferase-to-platelet ratio | 0.3(0.7);n=450 | 0.1(0.4);n=12488 | 0.37* | 0.2(0.5);n=309 | 0.1(0.2);n=8429 | 0.34* |
| Fibrosis-4-index | 3.0(4.2);n=332 | 1.7(5.1);n=7628 | 0.28* | 1.9(2.7);n=239 | 1.4(1.8);n=5545 | 0.22* |
| **Liver function tests** |  |  |  |  |  |  |
| Alkaline phosphatase, U/L | 91.4(50.3);n=1091 | 76.8(31.8);n=39271 | 0.35* | 85.3(32.2);n=874 | 75.1(27.0);n=25508 | 0.34* |
| Aspartate transaminase, U/L | 44.8(55.7);n=527 | 27.4(52.9);n=15520 | 0.32* | 43.4(42.4);n=416 | 26.7(26.3);n=10761 | 0.47* |
| Alanine transaminase, U/L | 47.6(43.0);n=933 | 28.2(33.8);n=33379 | 0.5* | 50.1(42.4);n=744 | 29.9(26.3);n=21299 | 0.57* |
| **Lipid and glucose profiles** |  |  |  |  |  |  |
| Triglyceride, mmol/L | 1.9(1.6);n=1173 | 1.7(1.5);n=47872 | 0.11 | 2.1(1.8);n=899 | 1.8(1.6);n=31008 | 0.21* |
| SD of triglyceride | 0.53(1.05);n=672 | 0.47(0.95);n=24728 | 0.06 | 0.7(1.1);n=587 | 0.5(1.0);n=17740 | 0.17 |
| Low-density lipoprotein, mmol/L | 2.39(0.83);n=1154 | 2.38(0.8);n=47082 | 0.01 | 2.5(0.8);n=879 | 2.3(0.8);n=30573 | 0.14 |
| SD of low-density lipoprotein | 0.3(0.3);n=642 | 0.4(0.3);n=24066 | 0.09 | 0.41(0.33);n=561 | 0.36(0.34);n=17295 | 0.17 |
| High-density lipoprotein, mmol/L | 1.17(0.33);n=1173 | 1.2(0.33);n=47805 | 0.07 | 1.1(0.3);n=899 | 1.2(0.3);n=30961 | 0.17 |
| SD of high-density lipoprotein | 0.11(0.1);n=651 | 0.1(0.08);n=23794 | 0.12 | 0.11(0.1);n=573 | 0.1(0.08);n=17062 | 0.15 |
| Total cholesterol, mmol/L | 4.4(1.0);n=1173 | 4.3(1.0);n=47922 | 0.06 | 4.5(1.0);n=899 | 4.3(1.0);n=31031 | 0.2 |
| SD of total cholesterol | 0.41(0.38);n=677 | 0.44(0.42);n=24744 | 0.06 | 0.5(0.4);n=588 | 0.4(0.4);n=17673 | 0.07 |
| Hemoglobin A1C, % | 7.99(1.57);n=1199 | 8.04(1.54);n=50043 | 0.03 | 8.3(1.4);n=893 | 8.2(1.5);n=32012 | 0.07 |
| SD of hemoglobin A1C | 0.62(0.73);n=885 | 0.56(0.72);n=35248 | 0.08 | 0.57(0.51);n=657 | 0.57(0.78);n=24077 | <0.01 |
| Fasting glucose, mmol/L | 9.0(3.8);n=1126 | 8.9(3.9);n=45307 | 0.03 | 9.2(3.5);n=814 | 9.1(3.6);n=28263 | 0.03 |
| SD of fasting glucose | 2.1(2.1);n=792 | 2.0(2.2);n=28380 | 0.05 | 2.1(1.8);n=652 | 1.9(1.9);n=19517 | 0.14 |

**Supplementary Table 4A. Baseline and clinical characteristics of patient sex groups before and after propensity score matching (1:1).**

* for SMD≥0.2; SD: standard deviation; SGLT2I: sodium glucose cotransporter-2 inhibitor; DPP4I: dipeptidyl peptidase-4 inhibitor; ACEis: Angiotensin-converting enzyme inhibitors; ARBs: angiotensin receptor blockers; MDRD: modification of diet in renal disease; NAFLD: Non-alcoholic fatty liver disease; # indicated the characteristics difference significance.

| **Characteristics** | **Before matching** |  | **SMD^#^** | **After matching** |  | **SMD^#^** |
| --- | --- | --- | --- | --- | --- | --- |
|  | **Male gender (N=34601) Mean(SD);N or Count(%)** | **Female gender (N=28098) Mean(SD);N or Count(%)** |  | **Male gender (N=27076) Mean(SD);N or Count(%)** | **Female gender (N=17232) Mean(SD);N or Count(%)** |  |
| **Demographics** |  |  |  |  |  |  |
| Baseline age, years | 61.8(12.4);n=34601 | 65.1(13.2);n=28098 | 0.26* | 57.8(10.6);n=27076 | 59.7(11.6);n=17232 | 0.17 |
| **Past comorbidities** |  |  |  |  |  |  |
| Charlson standard comorbidity index | 2.0(1.5);n=34601 | 2.3(1.6);n=28098 | 0.22* | 1.5(1.2);n=27076 | 1.7(1.3);n=17232 | 0.14 |
| Alcohol dependence | 124(0.35%) | 16(0.05%) | 0.07 | 51(0.18%) | 7(0.04%) | 0.04 |
| Hyperlipidaemia | 908(2.62%) | 818(2.91%) | 0.02 | 801(2.95%) | 653(3.78%) | 0.05 |
| Overweight, obesity and hyperalimentation | 240(0.69%) | 204(0.72%) | <0.01 | 349(1.28%) | 333(1.93%) | 0.05 |
| Hypertension | 7679(22.19%) | 7642(27.19%) | 0.12 | 5392(19.91%) | 4269(24.77%) | 0.12 |
| Chronic liver disease and cirrhosis | 735(2.12%) | 574(2.04%) | 0.01 | 694(2.56%) | 491(2.84%) | 0.02 |
| HBV infection | 1530(4.42%) | 913(3.24%) | 0.06 | 1412(5.21%) | 694(4.02%) | 0.06 |
| HCV infection | 97(0.28%) | 71(0.25%) | 0.01 | 64(0.23%) | 48(0.27%) | 0.01 |
| History of acute liver injury | 127(0.36%) | 72(0.25%) | 0.02 | 52(0.19%) | 36(0.20%) | <0.01 |
| Other liver disease | 390(1.12%) | 317(1.12%) | <0.01 | 245(0.90%) | 158(0.91%) | <0.01 |
| Gallstone | 39(0.11%) | 34(0.12%) | <0.01 | 38(0.14%) | 18(0.10%) | 0.01 |
| Bilirary disease | 509(1.47%) | 395(1.40%) | 0.01 | 280(1.03%) | 193(1.12%) | 0.01 |
| Disease of pancreas | 216(0.62%) | 211(0.75%) | 0.02 | 160(0.59%) | 125(0.72%) | 0.02 |
| Heart failure | 1085(3.13%) | 1025(3.64%) | 0.03 | 705(2.60%) | 393(2.28%) | 0.02 |
| Ischemic heart disease | 4189(12.10%) | 2225(7.91%) | 0.14 | 3842(14.18%) | 1323(7.67%) | 0.21* |
| Acute myocardial infarction | 1235(3.56%) | 518(1.84%) | 0.11 | 1197(4.42%) | 279(1.61%) | 0.16 |
| Peripheral vascular disease | 310(0.89%) | 189(0.67%) | 0.03 | 174(0.64%) | 75(0.43%) | 0.03 |
| Diabetic retinopathy | 2287(6.60%) | 2183(7.76%) | 0.04 | 1589(5.86%) | 1257(7.29%) | 0.06 |
| Baseline non-alcoholic fatty liver disease | 634(1.83%) | 547(1.94%) | 0.01 | 614(2.26%) | 466(2.70%) | 0.03 |
| Cancer except hepatocellular carcinoma | 805(2.32%) | 951(3.38%) | 0.06 | 410(1.51%) | 494(2.86%) | 0.09 |
| Duration from earliest diabetes mellitus diagnosis date to baseline date, day | 515.3(1214.5);n=34601 | 622.5(1348.9);n=28098 | 0.08 | 471.8(1190.8);n=27076 | 635.7(1426.4);n=17232 | 0.12 |
| **Medications** |  |  |  |  |  |  |
| SGLT2I v.s. DPP4I | 13257(38.31%) | 8897(31.66%) | 0.14 | 13257(48.96%) | 8897(51.63%) | 0.05 |
| SGLT2I frequency | 6.8(9.2);n=13257 | 7.9(10.8);n=8897 | 0.11 | 6.8(9.2);n=13257 | 7.9(10.8);n=8897 | 0.11 |
| DPP4I frequency | 5.1(7.7);n=21344 | 5.4(7.1);n=19201 | 0.04 | 4.0(5.6);n=13819 | 4.6(7.5);n=8335 | 0.09 |
| SGLT2I duration, days | 499.5(662.7);n=13257 | 573.0(682.4);n=8897 | 0.11 | 499.5(662.7);n=13257 | 573.0(682.4);n=8897 | 0.11 |
| DPP4I duration, days | 504.0(285.2);n=21344 | 505.9(286.3);n=19201 | 0.01 | 496.0(285.7);n=13819 | 458.3(286.3);n=8335 | 0.13 |
| Metformin | 30518(88.19%) | 25053(89.16%) | 0.03 | 24683(92.95%) | 16603(93.52%) | 0.02 |
| Sulphonylurea | 26525(76.65%) | 21530(76.62%) | <0.01 | 19689(74.14%) | 13017(73.32%) | 0.02 |
| Insulin | 17300(49.99%) | 14596(51.94%) | 0.04 | 13184(49.64%) | 9515(53.59%) | 0.08 |
| Acarbose | 3755(10.85%) | 4197(14.93%) | 0.12 | 844(3.17%) | 677(3.81%) | 0.03 |
| Glucagon-like peptide-1 receptor agonists | 2128(6.15%) | 2034(7.23%) | 0.04 | 1464(5.51%) | 1045(5.88%) | 0.02 |
| Thiozolidinedone | 9833(28.41%) | 9343(33.25%) | 0.1 | 6662(25.08%) | 4983(28.06%) | 0.07 |
| ACEI | 2902(8.38%) | 1816(6.46%) | 0.07 | 2095(7.88%) | 753(4.24%) | 0.15 |
| ARB | 1073(3.10%) | 1048(3.72%) | 0.03 | 802(3.02%) | 435(2.45%) | 0.03 |
| Antihypertensive drugs | 1846(5.33%) | 1688(6.00%) | 0.03 | 945(3.55%) | 296(1.66%) | 0.12 |
| HCV treatment drugs | 454(1.31%) | 193(0.68%) | 0.06 | 494(1.86%) | 151(0.85%) | 0.09 |
| HBV treatment drugs | 557(1.60%) | 230(0.81%) | 0.07 | 596(2.24%) | 185(1.04%) | 0.09 |
| Aspirin | 2020(5.83%) | 1238(4.40%) | 0.06 | 1581(5.95%) | 410(2.30%) | 0.18 |
| Anticoagulants | 6148(17.76%) | 4338(15.43%) | 0.06 | 5626(21.18%) | 2534(14.27%) | 0.18 |
| Antiplatelets | 2881(8.32%) | 1645(5.85%) | 0.1 | 2268(8.54%) | 635(3.57%) | 0.21* |
| Lipid-lowering drugs | 4943(14.28%) | 3362(11.96%) | 0.07 | 4660(17.54%) | 2178(12.26%) | 0.15 |
| Statins and fibrates | 18988(54.87%) | 14884(52.97%) | 0.04 | 18397(69.27%) | 12710(71.59%) | 0.05 |
| Nitrates | 1498(4.32%) | 815(2.90%) | 0.08 | 1273(4.79%) | 330(1.85%) | 0.16 |
| Diuretics | 3102(8.96%) | 2250(8.00%) | 0.03 | 2288(8.61%) | 949(5.34%) | 0.13 |
| Beta-blockers | 2217(6.40%) | 1401(4.98%) | 0.06 | 1818(6.84%) | 578(3.25%) | 0.16 |
| Calcium channel blockers | 7151(20.66%) | 5811(20.68%) | <0.01 | 6062(22.82%) | 3706(20.87%) | 0.05 |
| Steroids | 39(0.11%) | 57(0.20%) | 0.02 | 24(0.09%) | 44(0.24%) | 0.04 |
| **Calculated biomarkers** |  |  |  |  |  |  |
| Abbreviated MDRD, mL/min/1.73m^2 | 85.7(39.0);n=28233 | 86.0(42.0);n=24103 | 0.01 | 84.5(26.7);n=19298 | 87.6(31.0);n=13932 | 0.11 |
| Neutrophil-to-lymphocyte ratio | 3.7(4.8);n=14200 | 3.5(4.8);n=11954 | 0.03 | 3.2(4.2);n=9749 | 2.8(3.1);n=7521 | 0.11 |
| Aspartate aminotransferase-to-platelet ratio | 0.2(0.4);n=7192 | 0.1(0.4);n=5746 | 0.05 | 0.13(0.17);n=5258 | 0.12(0.2);n=3480 | 0.07 |
| Fibrosis-4-index | 1.7(2.8);n=4392 | 1.8(6.9);n=3568 | 0.02 | 1.44(2.05);n=3370 | 1.36(1.58);n=2414 | 0.04 |
| **Liver function tests** |  |  |  |  |  |  |
| Aspartate transaminase, U/L | 28.8(45.4);n=8939 | 27.0(61.4);n=7108 | 0.03 | 27.2(23.6);n=6704 | 27.5(32.0);n=4473 | 0.01 |
| Alanine transaminase, U/L | 30.8(32.4);n=18496 | 26.3(36.1);n=15816 | 0.13 | 31.7(27.2);n=12182 | 29.2(27.2);n=9861 | 0.09 |
| Bilirubin, umol/L | 12.0(7.7);n=21831 | 10.2(6.0);n=18333 | 0.26* | 11.9(6.0);n=14918 | 10.2(5.6);n=11410 | 0.29* |
| **Lipid and glucose profiles** |  |  |  |  |  |  |
| Triglyceride, mmol/L | 1.71(1.62);n=26633 | 1.71(1.28);n=22412 | <0.01 | 1.78(1.81);n=18385 | 1.77(1.32);n=13522 | 0.01 |
| SD of triglyceride | 0.5(1.0);n=14150 | 0.4(0.8);n=11250 | 0.07 | 0.5(1.1);n=10717 | 0.4(0.8);n=7610 | 0.11 |
| Low-density lipoprotein, mmol/L | 2.3(0.8);n=26165 | 2.4(0.8);n=22071 | 0.09 | 2.3(0.8);n=18105 | 2.4(0.8);n=13347 | 0.11 |
| SD of low-density lipoprotein | 0.35(0.33);n=13710 | 0.38(0.36);n=10998 | 0.08 | 0.3(0.3);n=10343 | 0.4(0.3);n=7513 | 0.07 |
| High-density lipoprotein, mmol/L | 1.1(0.3);n=26596 | 1.3(0.3);n=22382 | 0.5* | 1.1(0.3);n=18362 | 1.3(0.4);n=13498 | 0.49* |
| SD of high-density lipoprotein | 0.1(0.08);n=13595 | 0.11(0.09);n=10850 | 0.17 | 0.09(0.08);n=10247 | 0.11(0.09);n=7388 | 0.21* |
| Total cholesterol, mmol/L | 4.2(1.0);n=26658 | 4.5(1.0);n=22437 | 0.25* | 4.2(1.0);n=18398 | 4.5(1.0);n=13532 | 0.27* |
| SD of total cholesterol | 0.4(0.4);n=14135 | 0.5(0.4);n=11286 | 0.08 | 0.43(0.42);n=10630 | 0.45(0.41);n=7631 | 0.03 |
| Hemoglobin A1C, % | 8.03(1.57);n=27641 | 8.04(1.5);n=23601 | <0.01 | 8.1(1.5);n=18905 | 8.2(1.5);n=14000 | 0.08 |
| SD of hemoglobin A1C | 0.6(0.8);n=19495 | 0.5(0.6);n=16638 | 0.06 | 0.58(0.91);n=14014 | 0.56(0.54);n=10720 | 0.03 |
| Fasting glucose, mmol/L | 8.9(3.98);n=25088 | 8.9(3.81);n=21345 | <0.01 | 9.0(3.5);n=16741 | 9.1(3.6);n=12336 | 0.01 |
| SD of fasting glucose | 1.98(2.2);n=16011 | 2.05(2.16);n=13161 | 0.03 | 1.8(1.8);n=11423 | 2.0(1.9);n=8746 | 0.09 |

**Supplementary Table 4B. Baseline and clinical characteristics of patient age groups before and after propensity score matching (1:1).**

* for SMD≥0.2; SD: standard deviation; SGLT2I: sodium glucose cotransporter-2 inhibitor; DPP4I: dipeptidyl peptidase-4 inhibitor; ACEis: Angiotensin-converting enzyme inhibitors; ARBs: angiotensin receptor blockers; MDRD: modification of diet in renal disease; NAFLD: Non-alcoholic fatty liver disease; # indicated the characteristics difference significance.

| **Characteristics** | **Before matching** |  | **SMD^#^** | **After matching** |  | **SMD^#^** |
| --- | --- | --- | --- | --- | --- | --- |
|  | **Age>=65 (N=27437) Mean(SD);N or Count(%)** | **Age<65 (N=35262) Mean(SD);N or Count(%)** |  | **Age>=65 (N=12453) Mean(SD);N or Count(%)** | **Age<65 (N=31855) Mean(SD);N or Count(%)** |  |
| Demographics |  |  |  |  |  |  |
| Male gender | 13400(48.83%) | 21201(60.12%) | 0.23* | 6703(53.82%) | 20373(63.95%) | 0.21* |
| Female gender | 14037(51.16%) | 14061(39.87%) | 0.23* | 5750(46.17%) | 11482(36.04%) | 0.21* |
| **Past comorbidities** |  |  |  |  |  |  |
| Charlson standard comorbidity index | 3.3(1.3);n=27437 | 1.2(1.0);n=35262 | 1.9* | 2.9(1.0);n=12453 | 1.1(0.9);n=31855 | 1.81* |
| Alcohol dependence | 42(0.15%) | 98(0.27%) | 0.03 | 10(0.08%) | 48(0.15%) | 0.02 |
| Hyperlipidaemia | 827(3.01%) | 899(2.54%) | 0.03 | 495(3.97%) | 959(3.01%) | 0.05 |
| Overweight, obesity and hyperalimentation | 49(0.17%) | 395(1.12%) | 0.12 | 145(1.16%) | 537(1.68%) | 0.04 |
| Hypertension | 9355(34.09%) | 5966(16.91%) | 0.40* | 4372(35.10%) | 5289(16.60%) | 0.43* |
| Chronic liver disease and cirrhosis | 460(1.67%) | 849(2.40%) | 0.05 | 281(2.25%) | 904(2.83%) | 0.04 |
| HBV infection | 713(2.59%) | 1730(4.90%) | 0.12 | 491(3.94%) | 1615(5.06%) | 0.05 |
| HCV infection | 49(0.17%) | 119(0.33%) | 0.03 | 18(0.14%) | 94(0.29%) | 0.03 |
| History of acute liver injury | 92(0.33%) | 107(0.30%) | 0.01 | 40(0.32%) | 48(0.15%) | 0.04 |
| Other liver disease | 372(1.35%) | 335(0.95%) | 0.04 | 152(1.22%) | 251(0.78%) | 0.04 |
| Gallstone | 38(0.13%) | 35(0.09%) | 0.01 | 37(0.29%) | 19(0.05%) | 0.06 |
| Bilirary disease | 607(2.21%) | 297(0.84%) | 0.11 | 237(1.90%) | 236(0.74%) | 0.1 |
| Disease of pancreas | 248(0.90%) | 179(0.50%) | 0.05 | 100(0.80%) | 185(0.58%) | 0.03 |
| Heart failure | 1532(5.58%) | 578(1.63%) | 0.21* | 557(4.47%) | 541(1.69%) | 0.16 |
| Ischemic heart disease | 4009(14.61%) | 2405(6.82%) | 0.25* | 2527(20.29%) | 2638(8.28%) | 0.35* |
| Acute myocardial infarction | 1054(3.84%) | 699(1.98%) | 0.11 | 666(5.34%) | 810(2.54%) | 0.14 |
| Peripheral vascular disease | 357(1.30%) | 142(0.40%) | 0.1 | 149(1.19%) | 100(0.31%) | 0.1 |
| Diabetic retinopathy | 2233(8.13%) | 2237(6.34%) | 0.07 | 870(6.98%) | 1976(6.20%) | 0.03 |
| Baseline non-alcoholic fatty liver disease | 427(1.55%) | 754(2.13%) | 0.04 | 261(2.09%) | 819(2.57%) | 0.03 |
| Cancer except hepatocellular carcinoma | 1224(4.46%) | 532(1.50%) | 0.17 | 466(3.74%) | 438(1.37%) | 0.15 |
| Duration from earliest diabetes mellitus diagnosis date to baseline date, day | 666.7(1373.9);n=27437 | 482.9(1191.1);n=35262 | 0.14 | 656.3(1436.9);n=12453 | 488.3(1224.6);n=31855 | 0.13 |
| **Medications** |  |  |  |  |  |  |
| SGLT2I v.s. DPP4I | 5739(20.91%) | 16415(46.55%) | 0.56* | 5739(46.08%) | 16415(51.53%) | 0.11 |
| SGLT2I frequency | 8.6(11.2);n=5739 | 6.8(9.3);n=16415 | 0.18 | 8.6(11.2);n=5739 | 6.8(9.3);n=16415 | 0.18 |
| DPP4I frequency | 5.2(8.49);n=21698 | 5.23(5.95);n=18847 | <0.01 | 4.4(6.4);n=6714 | 4.2(6.4);n=15440 | 0.04 |
| SGLT2I duration, days | 579.7(703.7);n=5739 | 511.3(659.1);n=16415 | 0.1 | 579.7(703.7);n=5739 | 511.3(659.1);n=16415 | 0.1 |
| DPP4I duration, days | 489.6(289.4);n=21698 | 522.5(280.5);n=18847 | 0.12 | 468.7(277.6);n=6714 | 487.5(290.1);n=15440 | 0.07 |
| Metformin | 23028(83.93%) | 32543(92.28%) | 0.26* | 11117(91.04%) | 30169(93.99%) | 0.11 |
| Sulphonylurea | 21617(78.78%) | 26438(74.97%) | 0.09 | 9464(77.51%) | 23242(72.40%) | 0.12 |
| Insulin | 15558(56.70%) | 16338(46.33%) | 0.21* | 6595(54.01%) | 16104(50.17%) | 0.08 |
| Acarbose | 4794(17.47%) | 3158(8.95%) | 0.25* | 625(5.11%) | 896(2.79%) | 0.12 |
| Glucagon-like peptide-1 receptor agonists | 1709(6.22%) | 2453(6.95%) | 0.03 | 235(1.92%) | 2274(7.08%) | 0.25* |
| Thiozolidinedone | 8155(29.72%) | 11021(31.25%) | 0.03 | 2636(21.58%) | 9009(28.06%) | 0.15 |
| ACEI | 2582(9.41%) | 2136(6.05%) | 0.13 | 752(6.15%) | 2096(6.53%) | 0.02 |
| ARB | 1222(4.45%) | 899(2.54%) | 0.1 | 405(3.31%) | 832(2.59%) | 0.04 |
| Antihypertensive drugs | 2441(8.89%) | 1093(3.09%) | 0.25* | 623(5.10%) | 618(1.92%) | 0.17 |
| HCV treatment drugs | 162(0.59%) | 485(1.37%) | 0.08 | 108(0.88%) | 537(1.67%) | 0.07 |
| HBV treatment drugs | 196(0.71%) | 591(1.67%) | 0.09 | 122(0.99%) | 659(2.05%) | 0.09 |
| Aspirin | 2029(7.39%) | 1229(3.48%) | 0.17 | 719(5.88%) | 1272(3.96%) | 0.09 |
| Anticoagulants | 5256(19.15%) | 5230(14.83%) | 0.12 | 2096(17.16%) | 6064(18.89%) | 0.04 |
| Antiplatelets | 2761(10.06%) | 1765(5.00%) | 0.19 | 1097(8.98%) | 1806(5.62%) | 0.13 |
| Lipid-lowering drugs | 4129(15.04%) | 4176(11.84%) | 0.09 | 2110(17.28%) | 4728(14.72%) | 0.07 |
| Statins and fibrates | 13292(48.44%) | 20580(58.36%) | 0.2 | 9231(75.60%) | 21876(68.15%) | 0.17 |
| Nitrates | 1367(4.98%) | 946(2.68%) | 0.12 | 683(5.59%) | 920(2.86%) | 0.14 |
| Diuretics | 3139(11.44%) | 2213(6.27%) | 0.18 | 1132(9.27%) | 2105(6.55%) | 0.1 |
| Beta-blockers | 2038(7.42%) | 1580(4.48%) | 0.12 | 811(6.64%) | 1585(4.93%) | 0.07 |
| Calcium channel blockers | 5908(21.53%) | 7054(20.00%) | 0.04 | 2927(23.97%) | 6841(21.31%) | 0.06 |
| Steroids | 44(0.16%) | 52(0.14%) | <0.01 | 4(0.03%) | 64(0.19%) | 0.05 |
| **Calculated biomarkers** |  |  |  |  |  |  |
| Abbreviated MDRD, mL/min/1.73m^2 | 70.8(35.3);n=24056 | 98.6(40.1);n=28280 | 0.74* | 74.5(24.1);n=10390 | 90.9(29.0);n=22840 | 0.62* |
| Neutrophil-to-lymphocyte ratio | 4.1(5.5);n=12924 | 3.1(4.1);n=13230 | 0.2* | 3.3(4.1);n=5769 | 2.9(3.5);n=11501 | 0.11 |
| Aspartate aminotransferase-to-platelet ratio | 0.15(0.57);n=6042 | 0.14(0.23);n=6896 | 0.03 | 0.13(0.24);n=2826 | 0.12(0.15);n=5912 | 0.03 |
| Fibrosis-4-index | 2.3(7.0);n=3892 | 1.3(1.7);n=4068 | 0.2* | 1.9(2.8);n=2021 | 1.1(1.0);n=3763 | 0.35* |
| **Liver function tests** |  |  |  |  |  |  |
| Aspartate transaminase, U/L | 27.2(73.8);n=7120 | 28.6(26.8);n=8927 | 0.03 | 25.9(37.9);n=3506 | 28.0(20.6);n=7671 | 0.07 |
| Alanine transaminase, U/L | 24.1(38.6);n=15949 | 32.8(29.3);n=18363 | 0.25* | 25.1(26.4);n=7150 | 33.3(27.2);n=14893 | 0.3* |
| Bilirubin, umol/L | 10.9(6.9);n=18289 | 11.4(7.1);n=21875 | 0.06 | 11.1(6.5);n=8224 | 11.2(5.6);n=18104 | 0.02 |
| **Lipid and glucose profiles** |  |  |  |  |  |  |
| Triglyceride, mmol/L | 1.5(1.0);n=21924 | 1.8(1.8);n=27121 | 0.22* | 1.6(1.0);n=9888 | 1.9(1.8);n=22019 | 0.2* |
| SD of triglyceride | 0.4(0.5);n=10287 | 0.5(1.1);n=15113 | 0.21* | 0.4(0.5);n=5423 | 0.6(1.2);n=12904 | 0.22* |
| Low-density lipoprotein, mmol/L | 2.2(0.8);n=21706 | 2.5(0.8);n=26530 | 0.3* | 2.2(0.7);n=9791 | 2.4(0.8);n=21661 | 0.3* |
| SD of low-density lipoprotein | 0.3(0.3);n=10162 | 0.4(0.4);n=14546 | 0.11 | 0.3(0.3);n=5342 | 0.4(0.3);n=12514 | 0.1 |
| High-density lipoprotein, mmol/L | 1.22(0.34);n=21898 | 1.17(0.32);n=27080 | 0.15 | 1.22(0.36);n=9878 | 1.18(0.32);n=21982 | 0.12 |
| SD of high-density lipoprotein | 0.11(0.09);n=9952 | 0.1(0.08);n=14493 | 0.1 | 0.1(0.08);n=5213 | 0.1(0.08);n=12422 | 0.08 |
| Total cholesterol, mmol/L | 4.2(0.9);n=21959 | 4.5(1.0);n=27136 | 0.31* | 4.1(0.9);n=9901 | 4.4(1.0);n=22029 | 0.32* |
| SD of total cholesterol | 0.4(0.4);n=10308 | 0.5(0.4);n=15113 | 0.13 | 0.4(0.4);n=5416 | 0.5(0.4);n=12845 | 0.15 |
| Hemoglobin A1C, % | 7.8(1.4);n=23298 | 8.2(1.6);n=27944 | 0.26* | 7.9(1.3);n=10287 | 8.3(1.6);n=22618 | 0.23* |
| SD of hemoglobin A1C | 0.55(0.81);n=15458 | 0.58(0.64);n=20675 | 0.04 | 0.5(1.0);n=7683 | 0.6(0.6);n=17051 | 0.04 |
| Fasting glucose, mmol/L | 8.6(3.7);n=20872 | 9.2(4.0);n=25561 | 0.15 | 8.6(3.2);n=8709 | 9.3(3.7);n=20368 | 0.2* |
| SD of fasting glucose | 2.1(2.2);n=12617 | 1.9(2.1);n=16555 | 0.08 | 1.8(1.8);n=6132 | 1.9(1.9);n=14037 | 0.06 |

**Supplementary Table 5A. Univariate Cox regression analysis models for New onset hepatocellular carcinoma and new onset NAFLD before and after propensity score matching (1:1).**

* for p≤ 0.05, ** for p ≤ 0.01, *** for p ≤ 0.001; HR: hazard ratio; CI: confidence interval; SD: standard deviation; SGLT2I: sodium glucose cotransporter-2 inhibitor; DPP4I: dipeptidyl peptidase-4 inhibitor; ACEis: Angiotensin-converting enzyme inhibitors; ARBs: angiotensin receptor blockers; MDRD: modification of diet in renal disease; NAFLD: Non-alcoholic fatty liver disease; # indicated the difference between SGLT2I users and DPP4I users.

|  | **Before matching** |  | **After matching** |  |
| --- | --- | --- | --- | --- |
| **Characteristics** | **New onset hepatocellular carcinoma**  **HR [95% CI];P value** | **New onset NAFLD**  **HR [95% CI];P value** | **New onset hepatocellular carcinoma**  **HR [95% CI];P value** | **New onset NAFLD**  **HR [95% CI];P value** |
| Demographics |  |  |  |  |
| Male gender | 2.48[1.87-3.30];<0.0001*** | 1.0[Reference] | 3.12[2.13-4.57];<0.0001*** | 1.0[Reference] |
| Female gender | 1.0[Reference] | 1.24[1.12-1.38];0.0001*** | 1.0[Reference] | 1.66[1.47-1.87];<0.0001*** |
| Baseline age, years | 1.03[1.02-1.04];<0.0001*** | 0.99[0.98-0.99];<0.0001*** | 1.06[1.05-1.07];<0.0001*** | 0.98[0.97-0.99];<0.0001*** |
| **Past comorbidities** |  |  |  |  |
| Charlson standard comorbidity index | 1.26[1.18-1.35];<0.0001*** | 0.99[0.95-1.03];0.5435 | 1.40[1.30-1.52];<0.0001*** | 0.91[0.87-0.96];0.0006*** |
| Alcohol dependence | 25.80[14.45-46.06];<0.0001*** | 9.96[6.65-14.91];<0.0001*** | 18.28[6.79-49.25];<0.0001*** | 3.82[1.59-9.21];0.0028** |
| Hyperlipidaemia | 0.85[0.38-1.91];0.6919 | 1.19[0.88-1.61];0.2585 | 0.16[0.02-1.11];0.0632 | 0.97[0.69-1.36];0.8668 |
| Overweight, obesity and hyperalimentation | 1.62[0.52-5.05];0.4082 | 2.87[1.96-4.19];<0.0001*** | 15.56[10.82-22.39];<0.0001*** | 5.46[4.35-6.86];<0.0001*** |
| Hypertension | 1.51[1.16-1.96];0.0021** | 1.22[1.09-1.38];0.0009*** | 3.19[2.40-4.26];<0.0001*** | 1.78[1.57-2.02];<0.0001*** |
| Chronic liver disease and cirrhosis | 4.62[3.01-7.09];<0.0001*** | 12.78[11.18-14.62];<0.0001*** | 2.00[1.06-3.79];0.0326* | 8.25[7.05-9.65];<0.0001*** |
| HBV infection | 9.82[7.48-12.88];<0.0001*** | 3.48[2.96-4.10];<0.0001*** | 7.27[5.25-10.08];<0.0001*** | 2.40[1.98-2.91];<0.0001*** |
| HCV infection | 15.73[8.36-29.60];<0.0001*** | 7.24[4.84-10.84];<0.0001*** | 0.00[0.00-Inf];0.9894 | 1.88[0.78-4.52];0.1594 |
| History of acute liver injury | 33.61[21.71-52.04];<0.0001*** | 27.10[21.56-34.07];<0.0001*** | 25.58[13.09-49.97];<0.0001*** | 15.64[10.81-22.62];<0.0001*** |
| Other liver disease | 7.31[4.58-11.66];<0.0001*** | 5.25[4.14-6.67];<0.0001*** | 4.23[1.99-9.00];0.0002*** | 4.69[3.46-6.37];<0.0001*** |
| Gallstone | 43.39[24.30-77.46];<0.0001*** | 11.90[7.38-19.20];<0.0001*** | 30.78[14.47-65.50];<0.0001*** | 8.37[4.62-15.15];<0.0001*** |
| Bilirary disease | 1.99[0.94-4.21];0.0731 | 1.89[1.35-2.64];0.0002*** | 1.01[0.25-4.06];0.9925 | 3.09[2.19-4.35];<0.0001*** |
| Disease of pancreas | 1.76[0.57-5.51];0.3282 | 2.03[1.28-3.24];0.0028** | 0.00[0.00-Inf];0.9887 | 1.87[1.08-3.24];0.0243* |
| Heart failure | 1.04[0.52-2.11];0.9088 | 1.36[1.04-1.78];0.0256* | 0.22[0.03-1.59];0.1351 | 1.73[1.28-2.34];0.0004*** |
| Ischemic heart disease | 1.19[0.82-1.75];0.3608 | 0.84[0.69-1.02];0.0712 | 0.61[0.36-1.06];0.0791 | 0.96[0.80-1.16];0.7076 |
| Acute myocardial infarction | 1.79[1.00-3.20];0.0484* | 0.68[0.46-1.01];0.0562 | 0.65[0.24-1.74];0.3904 | 0.38[0.22-0.64];0.0003*** |
| Peripheral vascular disease | 0.55[0.08-3.91];0.5489 | 1.03[0.55-1.92];0.9251 | 0.00[0.00-Inf];0.9899 | 0.67[0.25-1.80];0.4285 |
| Diabetic retinopathy | 1.01[0.62-1.63];0.9773 | 0.80[0.64-1.01];0.0562 | 0.40[0.16-0.97];0.0426* | 0.63[0.46-0.84];0.0020** |
| Baseline non-alcoholic fatty liver disease | 4.64[2.97-7.25];<0.0001*** | 14.49[12.67-16.57];<0.0001*** | 1.97[1.01-3.84];0.0480* | 9.43[8.07-11.02];<0.0001*** |
| Cancer except hepatocellular carcinoma | 1.19[0.59-2.40];0.6347 | 0.94[0.67-1.32];0.7159 | 4.95[3.01-8.15];<0.0001*** | 0.81[0.51-1.29];0.3693 |
| Duration from earliest diabetes mellitus diagnosis date to baseline date, day | 1.000[1.000-1.000];0.0812 | 1.000[1.000-1.000];0.6687 | 1.000[0.999-1.000];0.0012** | 1.000[1.000-1.000];0.6167 |
| **Medications** |  |  |  |  |
| SGLT2I v.s. DPP4I | 0.28[0.20-0.40];<0.0001*** | 0.75[0.67-0.84];<0.0001*** | 0.23[0.16-0.33];<0.0001*** | 0.58[0.51-0.66];<0.0001*** |
| SGLT2I frequency | 1.02[1.01-1.03];<0.0001*** | 1.02[1.01-1.02];<0.0001*** | 1.02[1.01-1.03];<0.0001*** | 1.02[1.01-1.02];<0.0001*** |
| DPP4I frequency | 1.011[1.006-1.015];<0.0001*** | 1.01[1.00-1.01];0.0009*** | 0.99[0.96-1.02];0.3559 | 1.05[1.04-1.05];<0.0001*** |
| SGLT2I duration, days | 1.000[1.000-1.001];0.0555 | 1.000[1.000-1.000];0.0265* | 1.000[1.000-1.001];0.0555 | 1.000[1.000-1.000];0.0265* |
| DPP4I duration, days | 1.000[0.999-1.000];0.0792 | 1.000[1.000-1.000];0.3018 | 1.001[1.000-1.001];0.0038** | 1.000[0.999-1.000];0.0211* |
| Metformin | 0.61[0.44-0.86];0.0041** | 0.95[0.80-1.13];0.5898 | 4.41[1.41-13.80];0.0108* | 1.91[1.39-2.62];0.0001*** |
| Sulphonylurea | 0.92[0.69-1.21];0.5389 | 1.11[0.97-1.26];0.1164 | 1.79[1.21-2.63];0.0032** | 1.14[0.99-1.31];0.0640 |
| Insulin | 4.55[3.32-6.25];<0.0001*** | 1.43[1.29-1.60];<0.0001*** | 0.92[0.69-1.23];0.5767 | 0.98[0.87-1.10];0.6986 |
| Acarbose | 1.32[0.95-1.86];0.1026 | 1.22[1.05-1.42];0.0097** | 1.12[0.53-2.39];0.7640 | 0.78[0.54-1.13];0.1929 |
| Glucagon-like peptide-1 receptor agonists | 0.45[0.22-0.91];0.0263* | 1.39[1.15-1.67];0.0006*** | 0.09[0.01-0.62];0.0149* | 2.06[1.70-2.49];<0.0001*** |
| Thiozolidinedone | 0.71[0.53-0.94];0.0183* | 1.23[1.10-1.37];0.0003*** | 0.57[0.39-0.84];0.0043** | 0.93[0.82-1.07];0.3355 |
| ACEI | 1.91[1.33-2.75];0.0005*** | 1.46[1.22-1.74];<0.0001*** | 0.73[0.38-1.43];0.3636 | 0.94[0.73-1.20];0.6107 |
| ARB | 1.58[0.90-2.76];0.1077 | 1.32[1.02-1.72];0.0381* | 0.75[0.28-2.02];0.5712 | 1.18[0.85-1.65];0.3232 |
| Antihypertensive drugs | 1.83[1.21-2.79];0.0046** | 1.23[0.99-1.53];0.0614 | 1.26[0.67-2.39];0.4749 | 0.87[0.64-1.19];0.3917 |
| HCV treatment drugs | 22.02[16.04-30.21];<0.0001*** | 5.79[4.58-7.32];<0.0001*** | 19.09[13.46-27.07];<0.0001*** | 3.55[2.68-4.68];<0.0001*** |
| HBV treatment drugs | 21.84[16.21-29.43];<0.0001*** | 5.10[4.07-6.39];<0.0001*** | 17.64[12.58-24.73];<0.0001*** | 3.06[2.33-4.02];<0.0001*** |
| Aspirin | 1.21[0.72-2.04];0.4671 | 1.47[1.19-1.81];0.0003*** | 0.46[0.17-1.25];0.1291 | 0.98[0.74-1.31];0.9040 |
| Anticoagulants | 1.92[1.46-2.53];<0.0001*** | 1.38[1.21-1.57];<0.0001*** | 0.62[0.40-0.96];0.0321* | 0.73[0.62-0.87];0.0003*** |
| Antiplatelets | 1.88[1.29-2.74];0.0010** | 1.34[1.11-1.61];0.0023** | 0.64[0.31-1.29];0.2103 | 0.86[0.66-1.11];0.2458 |
| Lipid-lowering drugs | 1.24[0.88-1.74];0.2136 | 1.16[1.00-1.34];0.0563 | 0.54[0.33-0.89];0.0153* | 0.72[0.60-0.87];0.0005*** |
| Statins and fibrates | 0.75[0.59-0.96];0.0234* | 1.18[1.06-1.32];0.0021** | 0.58[0.44-0.78];0.0003*** | 0.80[0.71-0.91];0.0007*** |
| Nitrates | 1.49[0.85-2.61];0.1588 | 1.38[1.07-1.77];0.0118* | 0.29[0.07-1.17];0.0827 | 0.68[0.46-1.00];0.0486* |
| Diuretics | 2.74[2.01-3.75];<0.0001*** | 1.91[1.64-2.23];<0.0001*** | 0.50[0.23-1.06];0.0698 | 0.82[0.64-1.05];0.1201 |
| Beta-blockers | 1.47[0.93-2.32];0.0984 | 1.29[1.05-1.59];0.0173* | 0.29[0.09-0.90];0.0316* | 0.84[0.63-1.12];0.2390 |
| Calcium channel blockers | 1.58[1.20-2.07];0.0010** | 1.14[1.00-1.29];0.0459* | 1.09[0.78-1.53];0.5990 | 1.07[0.93-1.23];0.3333 |
| Steroids | 0.00[0.00-Inf];0.9903 | 1.49[0.48-4.62];0.4911 | - | 1.18[0.30-4.74];0.8108 |
| **Calculated biomarkers** |  |  |  |  |
| Abbreviated MDRD, mL/min/1.73m^2 | 1.001[0.997-1.004];0.7361 | 1.003[1.002-1.004];<0.0001*** | 0.98[0.98-0.99];<0.0001*** | 1.009[1.007-1.011];<0.0001*** |
| Neutrophil-to-lymphocyte ratio | 1.03[1.00-1.05];0.0223* | 1.01[0.99-1.02];0.4181 | 1.06[1.04-1.08];<0.0001*** | 0.99[0.96-1.02];0.4489 |
| Aspartate aminotransferase-to-platelet ratio | 1.22[1.13-1.31];<0.0001*** | 1.18[1.13-1.24];<0.0001*** | 1.76[1.05-2.92];0.0303* | 2.11[1.84-2.43];<0.0001*** |
| Fibrosis-4-index | 1.01[1.00-1.02];0.0032** | 1.01[1.01-1.02];0.0001*** | 1.05[1.00-1.10];0.0549 | 1.04[1.02-1.06];0.0002*** |
| **Liver function tests** |  |  |  |  |
| Alkaline phosphatase, U/L | 1.01[1.00-1.01];<0.0001*** | 1.00[1.00-1.01];<0.0001*** | 1.01[1.00-1.01];0.0009*** | 1.006[1.005-1.007];<0.0001*** |
| Aspartate transaminase, U/L | 1.001[1.000-1.002];0.0033** | 1.001[1.001-1.001];<0.0001*** | 1.00[1.00-1.01];0.1251 | 1.01[1.00-1.01];<0.0001*** |
| Alanine transaminase, U/L | 1.002[1.001-1.003];<0.0001*** | 1.002[1.002-1.002];<0.0001*** | 1.01[1.00-1.01];<0.0001*** | 1.006[1.005-1.007];<0.0001*** |
| **Lipid and glucose profiles** |  |  |  |  |
| Triglyceride, mmol/L | 0.66[0.54-0.81];<0.0001*** | 1.04[1.02-1.06];0.0001*** | 0.62[0.48-0.79];0.0001*** | 1.05[1.04-1.07];<0.0001*** |
| SD of triglyceride | 0.62[0.35-1.08];0.0937 | 1.05[0.99-1.11];0.1173 | 0.90[0.60-1.33];0.5848 | 1.08[1.04-1.12];<0.0001*** |
| Low-density lipoprotein, mmol/L | 0.79[0.65-0.96];0.0178* | 1.01[0.94-1.09];0.7456 | 0.98[0.79-1.21];0.8306 | 1.17[1.09-1.26];<0.0001*** |
| SD of low-density lipoprotein | 0.38[0.16-0.88];0.0245* | 0.78[0.61-1.00];0.0529 | 0.10[0.02-0.41];0.0015** | 1.51[1.22-1.85];0.0001*** |
| High-density lipoprotein, mmol/L | 1.31[0.88-1.97];0.1855 | 0.80[0.66-0.96];0.0143* | 1.06[0.63-1.76];0.8351 | 0.57[0.46-0.71];<0.0001*** |
| SD of high-density lipoprotein | 16.72[2.97-94.02];0.0014** | 4.18[1.93-9.02];0.0003*** | 6.02[0.36-101.46];0.2126 | 5.36[2.30-12.48];0.0001*** |
| Total cholesterol, mmol/L | 0.77[0.65-0.90];0.0015** | 1.05[1.00-1.11];0.0709 | 0.82[0.68-1.00];0.0450* | 1.17[1.11-1.23];<0.0001*** |
| SD of total cholesterol | 0.38[0.18-0.80];0.0112* | 0.86[0.71-1.05];0.1381 | 0.22[0.07-0.65];0.0064** | 1.15[0.97-1.37];0.1118 |
| Hemoglobin A1C, % | 0.97[0.88-1.07];0.5393 | 0.98[0.94-1.02];0.2945 | 0.78[0.68-0.90];0.0006*** | 1.04[1.00-1.08];0.0544 |
| SD of hemoglobin A1C | 1.04[0.84-1.27];0.7308 | 1.08[1.02-1.15];0.0088** | 0.57[0.35-0.93];0.0244* | 1.00[0.90-1.10];0.9886 |
| Fasting glucose, mmol/L | 0.98[0.94-1.02];0.3755 | 1.01[0.99-1.02];0.3015 | 0.88[0.82-0.94];0.0004*** | 1.01[0.99-1.03];0.3231 |
| SD of fasting glucose | 1.03[0.96-1.11];0.4073 | 1.03[1.00-1.06];0.0663 | 1.06[0.95-1.19];0.2793 | 1.06[1.03-1.10];0.0004*** |

**Supplementary Table 5B. Univariate Cox regression analysis models for new onset hepatocellular carcinoma and new onset NAFLD before and after propensity score matching (1:1).**

* for p≤ 0.05, ** for p ≤ 0.01, *** for p ≤ 0.001; HR: hazard ratio; CI: confidence interval; SD: standard deviation; SGLT2I: sodium glucose cotransporter-2 inhibitor; DPP4I: dipeptidyl peptidase-4 inhibitor; ACEis: Angiotensin-converting enzyme inhibitors; ARBs: angiotensin receptor blockers; MDRD: modification of diet in renal disease; NAFLD: Non-alcoholic fatty liver disease; # indicated the difference between SGLT2I users and DPP4I users.

| **Characteristics** | **Before matching** |  | **After matching** |  |
| --- | --- | --- | --- | --- |
|  | **All-cause mortality**  **HR [95% CI];P value** | **Cancer-related mortality**  **HR [95% CI];P value** | **All-cause mortality**  **HR [95% CI];P value** | **Cancer-related mortality**  **HR [95% CI];P value** |
| Demographics |  |  |  |  |
| Male gender | 0.94[0.90-0.99];0.0145* | 1.08[0.95-1.21];0.2373 | 1.46[1.36-1.57];<0.0001*** | 1.46[1.26-1.68];<0.0001*** |
| Female gender | 1.0[Reference] | 1.0[Reference] | 1.0[Reference] | 1.0[Reference] |
| Baseline age, years | 1.09[1.09-1.10];<0.0001*** | 1.07[1.06-1.07];<0.0001*** | 1.070[1.066-1.073];<0.0001*** | 1.06[1.05-1.07];<0.0001*** |
| **Past comorbidities** |  |  |  |  |
| Charlson standard comorbidity index | 1.54[1.53-1.56];<0.0001*** | 1.46[1.42-1.49];<0.0001*** | 1.47[1.44-1.49];<0.0001*** | 1.37[1.32-1.42];<0.0001*** |
| Alcohol dependence | 4.05[3.09-5.31];<0.0001*** | 3.91[1.95-7.85];0.0001*** | 5.83[3.87-8.79];<0.0001*** | 1.87[0.47-7.48];0.3778 |
| Hyperlipidaemia | 0.97[0.84-1.12];0.6978 | 0.94[0.64-1.36];0.7294 | 0.57[0.45-0.73];<0.0001*** | 0.46[0.27-0.77];0.0036** |
| Overweight, obesity and hyperalimentation | 0.44[0.29-0.66];0.0001*** | 0.37[0.12-1.16];0.0887 | 0.45[0.31-0.67];0.0001*** | 0.07[0.01-0.50];0.0079** |
| Hypertension | 2.12[2.02-2.22];<0.0001*** | 1.30[1.14-1.49];0.0001*** | 1.24[1.15-1.34];<0.0001*** | 0.52[0.43-0.64];<0.0001*** |
| Chronic liver disease and cirrhosis | 0.95[0.80-1.12];0.5187 | 0.74[0.46-1.20];0.2185 | 0.42[0.31-0.57];<0.0001*** | 0.48[0.27-0.85];0.0113* |
| HBV infection | 0.81[0.71-0.92];0.0016** | 1.22[0.92-1.61];0.1610 | 1.60[1.40-1.81];<0.0001*** | 1.69[1.32-2.16];<0.0001*** |
| HCV infection | 2.04[1.48-2.82];<0.0001*** | 2.86[1.43-5.73];0.0031** | 3.65[2.55-5.23];<0.0001*** | - |
| History of acute liver injury | 3.36[2.63-4.28];<0.0001*** | 3.28[1.76-6.11];0.0002*** | 1.30[0.68-2.50];0.4321 | 0.56[0.08-3.97];0.5604 |
| Other liver disease | 1.92[1.63-2.26];<0.0001*** | 1.63[1.03-2.56];0.0353* | 1.36[1.01-1.84];0.0456* | 0.36[0.12-1.13];0.0799 |
| Gallstone | 1.56[0.90-2.68];0.1112 | 4.68[2.10-10.45];0.0002*** | 1.33[0.60-2.95];0.4895 | - |
| Bilirary disease | 2.01[1.74-2.32];<0.0001*** | 2.03[1.41-2.91];0.0001*** | 1.22[0.91-1.64];0.1844 | 0.73[0.35-1.54];0.4081 |
| Disease of pancreas | 1.68[1.34-2.10];<0.0001*** | 1.95[1.15-3.31];0.0128* | 0.92[0.60-1.42];0.7131 | 0.51[0.16-1.58];0.2414 |
| Heart failure | 4.09[3.80-4.41];<0.0001*** | 1.37[1.01-1.87];0.0408* | 3.01[2.63-3.44];<0.0001*** | 0.52[0.29-0.94];0.0303* |
| Ischemic heart disease | 1.70[1.60-1.82];<0.0001*** | 1.06[0.88-1.29];0.5403 | 1.24[1.13-1.36];<0.0001*** | 0.71[0.56-0.90];0.0042** |
| Acute myocardial infarction | 2.19[1.98-2.43];<0.0001*** | 1.45[1.06-1.98];0.0201* | 1.89[1.64-2.18];<0.0001*** | 0.92[0.63-1.35];0.6738 |
| Peripheral vascular disease | 3.77[3.25-4.38];<0.0001*** | 1.48[0.81-2.67];0.1996 | 3.40[2.64-4.38];<0.0001*** | 1.90[0.99-3.67];0.0548 |
| Diabetic retinopathy | 1.65[1.53-1.78];<0.0001*** | 0.91[0.71-1.16];0.4451 | 1.16[1.03-1.32];0.0187* | 0.61[0.43-0.85];0.0032** |
| Baseline non-alcoholic fatty liver disease | 0.93[0.78-1.11];0.4061 | 0.82[0.51-1.32];0.4177 | 0.35[0.24-0.50];<0.0001*** | 0.48[0.27-0.87];0.0158* |
| Cancer except hepatocellular carcinoma | 2.36[2.13-2.60];<0.0001*** | 4.85[4.03-5.85];<0.0001*** | 1.68[1.39-2.02];<0.0001*** | 2.88[2.16-3.83];<0.0001*** |
| Duration from earliest diabetes mellitus diagnosis date to baseline date, day | 1.000[1.000-1.000];<0.0001*** | 1.000[1.000-1.000];0.3401 | 1.000[1.000-1.000];<0.0001*** | 1.000[1.000-1.000];<0.0001*** |
| **Medications** |  |  |  |  |
| SGLT2I v.s. DPP4I | 0.16[0.15-0.17];<0.0001*** | 0.23[0.19-0.28];<0.0001*** | 0.20[0.18-0.22];<0.0001*** | 0.16[0.13-0.19];<0.0001*** |
| SGLT2I frequency | 1.02[1.01-1.02];<0.0001*** | 1.02[1.02-1.03];<0.0001*** | 1.02[1.01-1.02];<0.0001*** | 1.02[1.02-1.03];<0.0001*** |
| DPP4I frequency | 0.99[0.99-1.00];0.0001*** | 1.01[1.00-1.01];<0.0001*** | 1.01[1.00-1.01];0.0010** | 0.99[0.98-1.00];0.1325 |
| SGLT2I duration, days | 1.000[1.000-1.000];0.0020** | 1.000[1.000-1.000];0.3708 | 1.000[1.000-1.000];0.0020** | 1.000[1.000-1.000];0.3708 |
| DPP4I duration, days | 0.999[0.999-0.999];<0.0001*** | 0.999[0.999-0.999];<0.0001*** | 1.000[1.000-1.000];0.0011** | 1.000[0.999-1.000];0.0001*** |
| Metformin | 0.27[0.26-0.29];<0.0001*** | 0.60[0.51-0.70];<0.0001*** | 0.65[0.58-0.73];<0.0001*** | 0.82[0.65-1.05];0.1153 |
| Sulphonylurea | 1.03[0.97-1.09];0.2942 | 1.04[0.90-1.20];0.5692 | 1.10[1.02-1.19];0.0139* | 0.76[0.66-0.88];0.0001*** |
| Insulin | 5.20[4.88-5.53];<0.0001*** | 3.42[2.97-3.94];<0.0001*** | 2.14[1.99-2.30];<0.0001*** | 2.65[2.29-3.07];<0.0001*** |
| Acarbose | 2.02[1.91-2.13];<0.0001*** | 1.52[1.30-1.78];<0.0001*** | 1.51[1.29-1.76];<0.0001*** | 0.52[0.32-0.86];0.0102* |
| Glucagon-like peptide-1 receptor agonists | 0.86[0.78-0.95];0.0034** | 0.83[0.64-1.07];0.1545 | 0.26[0.20-0.34];<0.0001*** | 0.18[0.10-0.34];<0.0001*** |
| Thiozolidinedone | 0.98[0.93-1.03];0.3783 | 0.89[0.78-1.02];0.0865 | 0.65[0.59-0.70];<0.0001*** | 0.60[0.51-0.71];<0.0001*** |
| ACEI | 2.25[2.10-2.40];<0.0001*** | 1.61[1.33-1.94];<0.0001*** | 0.90[0.78-1.04];0.1543 | 0.60[0.43-0.84];0.0030** |
| ARB | 1.78[1.61-1.97];<0.0001*** | 1.62[1.24-2.12];0.0004*** | 0.59[0.46-0.76];0.0001*** | 0.50[0.29-0.87];0.0140* |
| Antihypertensive drugs | 2.48[2.31-2.66];<0.0001*** | 1.45[1.16-1.82];0.0013** | 1.02[0.87-1.20];0.7677 | 0.53[0.35-0.82];0.0043** |
| HCV treatment drugs | 0.92[0.72-1.16];0.4670 | 2.04[1.35-3.08];0.0007*** | 1.35[1.06-1.72];0.0158* | 2.94[2.11-4.08];<0.0001*** |
| HBV treatment drugs | 0.79[0.63-1.00];0.0467* | 1.66[1.10-2.52];0.0156* | 1.13[0.89-1.44];0.3052 | 2.40[1.73-3.33];<0.0001*** |
| Aspirin | 2.69[2.51-2.89];<0.0001*** | 1.77[1.43-2.20];<0.0001*** | 1.04[0.89-1.21];0.6461 | 0.73[0.51-1.05];0.0938 |
| Anticoagulants | 2.11[2.00-2.22];<0.0001*** | 1.87[1.63-2.14];<0.0001*** | 0.96[0.88-1.05];0.3897 | 1.04[0.88-1.23];0.6131 |
| Antiplatelets | 2.55[2.40-2.73];<0.0001*** | 1.77[1.47-2.14];<0.0001*** | 1.06[0.93-1.21];0.3889 | 0.71[0.53-0.97];0.0310* |
| Lipid-lowering drugs | 1.58[1.49-1.68];<0.0001*** | 1.42[1.21-1.66];<0.0001*** | 0.70[0.63-0.78];<0.0001*** | 0.52[0.41-0.65];<0.0001*** |
| Statins and fibrates | 0.51[0.49-0.54];<0.0001*** | 0.71[0.63-0.80];<0.0001*** | 0.50[0.47-0.53];<0.0001*** | 0.35[0.31-0.40];<0.0001*** |
| Nitrates | 2.60[2.39-2.83];<0.0001*** | 1.63[1.25-2.12];0.0003*** | 1.36[1.17-1.59];0.0001*** | 0.96[0.67-1.38];0.8340 |
| Diuretics | 3.06[2.89-3.25];<0.0001*** | 2.46[2.10-2.88];<0.0001*** | 1.32[1.18-1.48];<0.0001*** | 0.80[0.61-1.06];0.1163 |
| Beta-blockers | 2.55[2.37-2.73];<0.0001*** | 1.79[1.45-2.20];<0.0001*** | 1.25[1.09-1.43];0.0010** | 0.82[0.59-1.12];0.2108 |
| Calcium channel blockers | 1.29[1.22-1.36];<0.0001*** | 1.23[1.07-1.41];0.0036** | 0.75[0.69-0.82];<0.0001*** | 0.68[0.57-0.81];<0.0001*** |
| Steroids | 1.63[1.01-2.62];0.0454* | 0.62[0.09-4.39];0.6308 | - | - |
| **Calculated biomarkers** |  |  |  |  |
| Abbreviated MDRD, mL/min/1.73m^2 | 0.977[0.976-0.978];<0.0001*** | 0.99[0.99-1.00];<0.0001*** | 0.981[0.979-0.983];<0.0001*** | 1.00[0.99-1.00];0.1292 |
| Neutrophil-to-lymphocyte ratio | 1.04[1.03-1.04];<0.0001*** | 1.03[1.02-1.04];<0.0001*** | 1.04[1.04-1.05];<0.0001*** | 1.04[1.02-1.06];<0.0001*** |
| Aspartate aminotransferase-to-platelet ratio | 1.09[1.04-1.14];0.0007*** | 1.14[1.04-1.25];0.0042** | 1.49[1.22-1.81];0.0001*** | 1.50[0.86-2.62];0.1550 |
| Fibrosis-4-index | 1.010[1.007-1.013];<0.0001*** | 1.01[1.00-1.02];0.0265* | 1.05[1.03-1.06];<0.0001*** | 1.04[1.01-1.08];0.0118* |
| Aspartate aminotransferase-to-alanine transaminase ratio | 1.01[1.00-1.02];0.0042** | 1.01[0.98-1.03];0.6280 | 1.10[1.07-1.13];<0.0001*** | 1.08[0.98-1.18];0.1041 |
| **Liver function tests** |  |  |  |  |
| Alkaline phosphatase, U/L | 1.00[1.00-1.01];<0.0001*** | 1.00[1.00-1.01];<0.0001*** | 1.006[1.005-1.007];<0.0001*** | 1.000[0.995-1.004];0.8807 |
| Aspartate transaminase, U/L | 1.000[0.999-1.001];0.8749 | 1.001[1.000-1.002];0.1729 | 1.002[1.000-1.004];0.0398* | 0.99[0.97-1.00];0.0424* |
| Alanine transaminase, U/L | 0.99[0.98-0.99];<0.0001*** | 0.999[0.996-1.002];0.6203 | 0.989[0.986-0.992];<0.0001*** | 0.99[0.98-1.00];0.0153* |
| Bilirubin, umol/L | 0.99[0.98-0.99];<0.0001*** | 1.01[1.00-1.01];0.0011** | 1.00[0.99-1.01];0.7536 | 1.01[1.00-1.02];0.0573 |
| **Lipid and glucose profiles** |  |  |  |  |
| Triglyceride, mmol/L | 0.93[0.91-0.96];<0.0001*** | 0.76[0.70-0.83];<0.0001*** | 0.88[0.85-0.92];<0.0001*** | 0.70[0.62-0.80];<0.0001*** |
| SD of triglyceride | 0.92[0.87-0.97];0.0032** | 0.50[0.36-0.70];<0.0001*** | 0.77[0.68-0.87];<0.0001*** | 0.30[0.16-0.56];0.0001*** |
| Low-density lipoprotein, mmol/L | 0.93[0.90-0.96];<0.0001*** | 0.89[0.81-0.97];0.0101* | 0.96[0.90-1.01];0.1378 | 0.92[0.81-1.04];0.1679 |
| SD of low-density lipoprotein | 1.20[1.08-1.34];0.0008*** | 0.77[0.55-1.09];0.1430 | 1.16[0.97-1.38];0.1060 | 0.77[0.45-1.31];0.3346 |
| High-density lipoprotein, mmol/L | 1.09[1.00-1.18];0.0492* | 1.08[0.87-1.33];0.4884 | 1.05[0.92-1.20];0.4897 | 0.67[0.48-0.92];0.0136* |
| SD of high-density lipoprotein | 15.46[11.29-21.18];<0.0001*** | 5.91[2.09-16.76];0.0008*** | 10.29[5.67-18.70];<0.0001*** | 1.42[0.19-10.83];0.7331 |
| Total cholesterol, mmol/L | 0.92[0.90-0.95];<0.0001*** | 0.82[0.76-0.89];<0.0001*** | 0.90[0.85-0.94];<0.0001*** | 0.76[0.68-0.85];<0.0001*** |
| SD of total cholesterol | 1.22[1.13-1.32];<0.0001*** | 0.75[0.56-1.01];0.0602 | 1.04[0.90-1.20];0.6036 | 0.62[0.38-1.00];0.0488* |
| Hemoglobin A1C, % | 0.95[0.94-0.97];<0.0001*** | 0.90[0.85-0.94];<0.0001*** | 0.98[0.95-1.01];0.2584 | 0.84[0.78-0.90];<0.0001*** |
| SD of hemoglobin A1C | 1.14[1.11-1.16];<0.0001*** | 1.07[0.98-1.17];0.1289 | 1.09[1.04-1.13];0.0001*** | 1.01[0.86-1.18];0.9236 |
| Fasting glucose, mmol/L | 1.02[1.01-1.02];<0.0001*** | 0.99[0.97-1.01];0.4862 | 1.02[1.01-1.03];0.0010** | 0.93[0.90-0.96];0.0001*** |
| SD of fasting glucose | 1.11[1.10-1.12];<0.0001*** | 1.08[1.05-1.11];<0.0001*** | 1.14[1.12-1.16];<0.0001*** | 1.05[0.98-1.12];0.1501 |

**Supplementary Table 6. Subgroup analysis: Multivariate Cox regression models with adjustments to predict primary and secondary outcomes in the HBV infection**

**subgroup.**

* for p≤ 0.05, ** for p ≤ 0.01, *** for p ≤ 0.001; HR: hazard ratio; CI: confidence interval; SGLT2I: sodium glucose cotransporter-2 inhibitor; DPP4I: dipeptidyl peptidase-4 inhibitor; HBV: hepatitis B virus

Model 1 adjusted for significant demographics.

Model 2 adjusted for significant demographics, and past comorbidities.

Model 3 adjusted for significant demographics, past comorbidities, and non-SGLT2I/DPP4I medications.

Model 4 adjusted for significant demographics, past comorbidities, non-SGLT2I/DPP4I medications, abbreviated MDRD, fasting glucose, HbA1c, and duration from earliest diabetes mellitus date to initial drug exposure date.

^ Patients with prior NAFLD were excluded in the multivariate Cox regression models to predict new onset NAFLD.

|  | |  | **Model 1** | **Model 2** | **Model 3** | **Model 4** |
| --- | --- | --- | --- | --- | --- | --- |
| **SGLT2I v.s. DPP4I** | **HBV infection positive (n=** **2,106)** | **New onset NAFLD**  **HR [95% CI];P value^** | 0.68[0.45-1.04]; 0.0738 | 0.74[0.47-1.16]; 0.1897 | 0.81[0.51- 1.29] 0.3772 | 1.21[0.69- 2.13]; 0.5038 |
|  |  | **New onset HCC**  **HR [95% CI];P value** | 0.29[0.15-0.54]; <0.0001*** | 0.25[0.12-0.49]; <0.0001*** | 0.24[0.11-0.50]; 0.0001*** | 3.28[1.21-8.90]; 0.0200 * |
|  |  | **New onset cancer related mortality**  **HR [95% CI];P value** | 0.12[0.06-0.23]; <0.0001*** | 0.09[0.04-0.20];<0.0001*** | 0.10[0.04- 0.21];<0.0001*** | 0.08[0.05-0.19]; <0.0001*** |
|  |  | **New onset all-cause mortality**  **HR [95% CI];P value** | 0.10[0.07-0.15]; <0.0001*** | 0.10[0.07-0.15]; <0.0001*** | 0.11[0.07-0.16]; <0.0001*** | 0.09[0.04-0.12]; <0.0001*** |
|  | **HBV infection negative (n=** **42,202)** | **New onset NAFLD**  **HR [95% CI];P value^** | 0.45[0.39-0.52]; <0.0001*** | 0.44[0.38-0.51]; <0.0001*** | 0.43[0.37-0.50]; <0.0001*** | 0.35[0.30-0.42]; <0.0001*** |
|  |  | **New onset HCC**  **HR [95% CI];P value** | 0.21[0.16-0.39]; <0.0001*** | 0.25[0.16-0.39]; <0.0001*** | 0.27[0.18-0.43]; <0.0001*** | 0.30[0.18-0.50]; <0.0001*** |
|  |  | **New onset cancer related mortality**  **HR [95% CI];P value** | 0.17[0.14-21]; <0.0001*** | 0.18[0.15-21]; <0.0001*** | 0.19[0.16-0.23]; <0.0001*** | 0.32[0.25-0.41]; <0.0001*** |
|  |  | **New onset all-cause mortality**  **HR [95% CI];P value** | 0.22[0.20-0.24]; <0.0001*** | 0.22[0.20-0.24]; <0.0001*** | 0.24[0.22-0.27]; <0.0001*** | 0.30[0.27-0.34]; <0.0001*** |
